# Population-scale Long-read Sequencing in the *All of Us* Research Program

**DOI:** 10.1101/2025.10.02.25336942

**Authors:** Kiran V Garimella, Qiuhui Li, Julie Wertz, Samuel K Lee, Fabio Cunial, Yongqing Huang, Yulia Mostovoy, Ryan Lorig-Roach, Adam English, Hang Su, Shawn Levy, Donna M Muzny, Chelsea Berngruber, Matt C Danzi, William T Harvey, Emily L LaPlante, Karynne Patterson, Allison N Rozanski, Sophie Schwartz, Beri Shifaw, Yuanyuan Wang, Isaac Wong, Isaac R. L. Xu, Shadi Zaheri, Stephan Zuchner, Xinchang Zheng, Shannon Dugan-Perez, Michal Izydorczyk, Heer Mehta, Richard A Gibbs, Lee Lichtenstein, Namrata Gupta, Niall Lennon, Stacey Gabriel, All of Us Research Program Long Read Working Group, Winston Timp, Kimberly F Doheny, Tara Dutka, Anjene Musick, Chia-Lin Wei, Fritz J Sedlazeck, Michael C Schatz, Michael E Talkowski, Evan E Eichler

**Affiliations:** All of Us Research Program, National Institutes of Health, Bethesda, MD, USA; Broad, Color, and Mass General Brigham Laboratory for Molecular Medicine, Boston, MA, USA; Center for Genomic Medicine, Massachusetts General Hospital, Boston, MA, USA; Program in Medical and Population Genetics, Broad Institute of MIT and Harvard, Cambridge, MA, USA; Department of Neurology, Massachusetts General Hospital and Harvard Medical School, Boston, MA, USA; Data Sciences Platform, Broad Institute of MIT and Harvard, Cambridge, MA, USA; Department of Computer Science, Johns Hopkins University, Baltimore MD, USA; Department of Genetic Medicine, Johns Hopkins University School of Medicine, Baltimore, MD, USA; Department of Genome Sciences, University of Washington School of Medicine, Seattle, WA, USA; Department of Human Genetics and John P. Hussman Institute for Human Genomics, University of Miami Miller School of Medicine, Miami, FL, USA; HudsonAlpha Institute for Biotechnology; Human Genome Sequencing Center, Baylor College of Medicine, Houston, TX, USA; Vanderbilt Institute for Clinical and Translational Research, Vanderbilt University Medical Center, Nashville, TN, USA

## Abstract

The *All of Us* Research Program (AoU) is a national biobank seeking to enroll one million individuals in the United States to link genomic and biomedical data, including short- and long-read whole-genome sequencing (srWGS/LRS), with rich electronic health record (EHR) information. Here, we present the first large-scale analyses of long-read sequencing (LRS) in AoU and offer a new framework for deriving genomic insights into complex structural variation (SV) of relevance to human health and disease. We performed joint analyses of 1,027 individuals self-identifying as Black or African American, sequenced to ∼8x coverage with Pacific Biosciences HiFi technology and processed using cloud-native pipelines. From these LRS data we constructed a comprehensive variant callset encompassing known (*FMR1* and *HTT*) and novel repeat expansions, clinically relevant haplotypes at loci inaccessible to srWGS, and haplotypes relevant to disease risk (HLA) and pharmacogenomics (*CYP2D6)*, including SNVs, indels, and SVs. We developed methods for cohort-level variant calling and a scalable workflow to impute >750,000 of these SVs into existing srWGS datasets for trait association and human disease studies. Expanding to 10,000 self-identified Black or African American AoU participants with srWGS and matched EHRs, we identified 291 SV-disease associations (p < 1×10⁻⁵) spanning 226 conditions with 50.9% of associations involving SVs absent from the matched srWGS callset. Across the 226 traits, after fine-mapping using SVs and SNVs we identified 191 SV-disease pairs spanning 160 traits (70.8%) where the SV had the strongest association within the locus. Associations specific to those with computed ancestry similar to the African reference population exhibited larger effect sizes and lower allele frequencies, consistent with high-risk, ancestry-specific variants. These results demonstrate that the integration of LRS into AoU and future biobank initiatives can provide transformative new insights into genomic variation with potentially profound impact on precision medicine.

## INTRODUCTION

The NIH-funded *All of Us* Research Program (AoU) is aggregating longitudinal electronic health records (EHRs), physical measurements, survey responses, and genome sequences for one million participants in the United States^1,2^. Current enrollment has surpassed 864,000 people, with recruitment emphasis on individuals from groups whose health outcomes remain understudied. A comprehensive catalog of EHRs, phenotypes, and genetic variation in individuals representative of the U.S. population can facilitate the identification of clinically relevant and potentially actionable variants, identify putative disease biomarkers, elucidate the genetic architecture of complex and quantitative traits, and further advance the goal of widespread access to genomic medicine. To this end, the AoU Curated Data Repository (CDR; version 8) now hosts genomic data from short-read whole-genome sequencing (srWGS) on 414,830 individuals, with more to come in future releases. These AoU data are freely accessible to qualified researchers at authorized institutions for diverse biomedical research applications via the cloud-based Researcher Workbench^3^.

Despite the release of ∼1.1 billion genomic variants from AoU participants, including single-nucleotide variants (SNVs), small insertion/deletion variants (indels), and structural variants (SVs), srWGS continues to leave significant gaps in genomic variant discovery for human disease and trait associations. At present, srWGS is the dominant technology in biomedical research, forms the basis of the most widely used variant databases^4,5^, and is the primary technology used in clinical genomics to diagnose patients with genetic disease^6^. While highly cost-effective, its reliance on 300 bases of paired-end reads and 500-800 bp fragment sizes limits access to many genomic regions and classes of genetic variation^7–12^. These blind spots, particularly in repetitive regions and discovery of the complex rearrangements that are mediated by such sequences, are well documented^13^. Such regions are frequently associated with disease, often stratified between different populations, and are most likely to be misrepresented or absent in a singular reference sequence^14^. As a result, more genetically diverse populations are underrepresented in both variant catalogs and downstream analyses.

Long-read sequencing (LRS) technologies can yield reads that span much longer sequences in the human genome (often exceeding 15,000 bases) that offers a powerful solution to interrogate many of the blind spots in the human genome that were refractory to srWGS. These long reads enable near-complete ascertainment of genomic variation, either through alignment to a reference genome or via diploid *de novo* assembly without reliance on a reference^12,15–18^. The read lengths span the majority of pathogenic repeat expansions and contractions, allow for single-base-pair breakpoint resolution of SVs, and capture haplotype backgrounds through the physical phasing of dozens of variants per read^19,20^. However, despite these advantages, two major gaps remain: most LRS studies have been limited to a small number of well-characterized genomes, and very few population-level efforts have integrated LRS with EHRs or other rich phenotypic datasets at scale.

Recent advances in LRS technologies have dramatically increased throughput and reduced costs, making population-scale trait association and clinical studies more feasible. In response, All of Us launched one of the first and largest surveys of human genetic variation using Pacific Biosciences (PacBio) HiFi sequencing in individuals with matched longitudinal EHRs and srWGS data. In this Phase 1 study, we developed open-source computational tools for scalable LRS analysis and comprehensively characterized genomic variation among genetically diverse individuals self-identified as Black or African American. These analyses uncovered a broad spectrum of genomic variants, quantitative trait loci (QTLs), trait associations, and population-level diversity at previously intractable loci, offering an early glimpse into the potential of LRS to advance precision medicine. Here we present the results of Phase 1, while noting that Phase 2, currently underway, will extend this effort more comprehensively across populations.

## RESULTS

### Participant selection and LRS

We selected 1,027 self-identified Black or African American participants over 20 years of age who met all requirements for sequencing library construction (Methods) from the AoU CDR version 7 release of 245,388 participants recruited from sites across the U.S. (Fig. 1a-d). This focus was motivated by three considerations: (1) Black and African American self-identity reflects a socio-political population construct that captures the lived experiences, social context, and structural factors that shape health outcomes. However, lower representation of individuals who self-identify as Black and African American in genomics research has made it challenging to characterize the full impact of how these factors intersect with genetic variation to influence health outcomes and disease risk in this population; (2) many, but not all, individuals who self-identify as Black or African American have substantial African genetic ancestry, which may enrich certain variants; and (3) populations of African genetic ancestry harbor the highest levels of genetic diversity across geographic populations, which increases the power of variant discovery, particularly for SVs. Approximately 96% of sequenced participants had associated phenotypic data as part of their EHR (general phenotype distribution in Fig. 1e). Importantly, this information was not used to inform selection criteria.

**Fig. 1:**
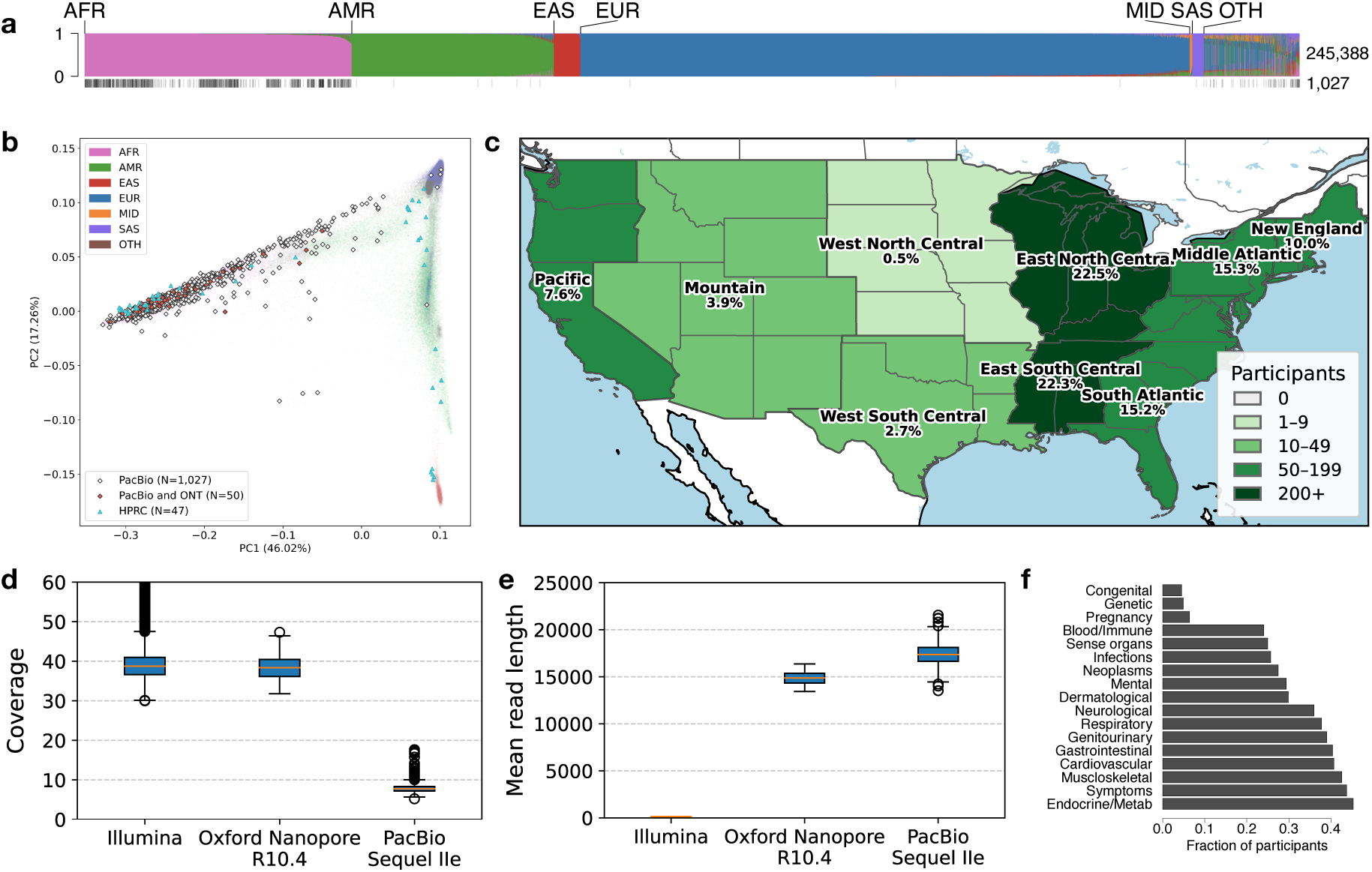
LRS of self-identified Black or African American participants from *All of Us*. **a,** 1,027 participants selected for PacBio HiFi sequencing (black vertical lines) from 245,388 AoU participants (top multicolored bar) with existing short-read data in CDRv7 releases and beyond. Computed genetic ancestry from srWGS data shown for: AFR–African, AMR–Admixed American, EAS– East Asian, EUR–European, MID–Middle Eastern, SAS–South Asian, OTH–other (typically admixed). **b,** PCA of SNV genetic data from participants selected for PacBio LRS (gray diamonds), overlapping participants with Oxford Nanopore Technologies (ONT) data (red, filled diamonds), and participants from the Human Pangenome Reference Consortium (HPRC) for comparison and evaluation (blue triangles), projected onto the AoU srWGS data. **c,** Geographic sampling locations for 1,027 long-read participants collected across the continental U.S., shown at the US Census Bureau Census Division level (no LRS participants sampled from Alaska, Hawaii, or other territories). **d,** Coverage distributions of selected LRS participants, stratified by sequencing technology. **e,** Mean read lengths of selected LRS participants, stratified by sequencing technology. **f,** Clinical phenotype distribution in AoU-LR participants based on EHR data. Diagnoses were mapped using the Observational Medical Outcomes Partnership (OMOP) concept IDs and interpreted via standard Observational Health Data Sciences and Informatics (OHDSI) concept mappings. For each disease category, all descendant concept IDs were identified using the OHDSI CONCEPT_ANCESTOR table, and counts reflect the number of unique participants with a diagnosis matching any related concept.

Participant selection for LRS occurred in 2020 before either genotyping array or srWGS data generation for *All of Us* participants began, precluding the use of either for selection or ancestry confirmation prior to sequencing. Nevertheless, after srWGS data became available, a principal component analysis (PCA) with ancestry labels computationally inferred using a random forest classifier trained on 1000 Genomes Project (1KGP) and the Human Genome Diversity Project (HGDP) autosomal variants from gnomAD v3.1^3^ confirmed that the selected participants were of majority genetically similar to African reference populations or admixed across reference (Fig. 1b).

For LRS data generation, we used the PacBio (Sequel IIe) platform, which provides highly accurate reads through circular consensus sequencing (CCS) and outperforms other LRS technologies in base-level accuracy^21^. To enable scalable population-level variant discovery, we adopted a mid-pass sequencing strategy targeting ∼8x coverage. Our groups have previously shown that this strategy increases SV discovery sensitivity at substantially lower cost than deep LRS using multiple technologies to completely assemble diploid genomes^20,22^. Compared to Illumina srWGS performed on the same individuals, the average length of LRS reads was 115-fold longer (mean=17.5 kbp, Table 1). As a control for variant discovery, a subset (n=50) of participants were also sequenced at 35x sequence coverage using Oxford Nanopore Technologies (ONT) (Table 1, Fig. 1). The combined dataset (termed the “CDRv7 AoU Long-Read dataset”), along with variant calls and local assemblies, is already available to approved researchers via the AoU Researcher Workbench (RW).

**Table 1.** Sequencing Summary of *All of Us* Participants in CDRv7.

|  | Illumina | PacBio Sequel IIe | ONT R10.4 |
| --- | --- | --- | --- |
| **Participants** | 245,388 | 1,027 / 245,388 | 50 / 1,027 |
| **Sex at birth** |  |  |  |
| *female* | 145,563 | 724 | 34 |
| *male* | 94,756 | 295 | 16 |
| *PMI: Skip* | 2,532 | 0 | 0 |
| *Unknown* | 2,229 | 0 | 0 |
| **Participant Provided Information Survey, question 2: “What category best describes you?”** |  |  |  |
| *Black or African American* | 50,969 | 952 | 46 |
| *More than one population* | 4,273 | 75 | 4 |
| *PMI: Skip* | 5,387 | 0* | 0 |
| *Other* | 188,759 | 0 | 0 |
| **EHRs** | 206,173 (84.0%) | 985 (95.9%) | 48 (96.0%) |
| **Physical measurements** | 245,149 (99.9%) | 1,027 (100.0%) | 1,027 (100.0%) |
| **Survey responses** | 245,388 (100.0%) | 1,027 (100.0%) | 1,027 (100.0%) |
| **Sequencing metrics** |  |  |  |
| *coverage* | 39.0 ± 4.3 | 8.0 ± 1.3 | 35.9 ± 4.8 |
| *read length (bp)* | 151 | 17,415 ± 1,106 | 14,867 ± 795 |

\* Note: In the All of Us curated data repository version 7 (CDRv7), 13 participants erroneously reported as having self-identified status of “PMI: Skip” (indicating survey responses where participants explicitly chose not to answer a given question). This is scheduled to be corrected in a future release.
Demographic and sequencing metrics of AoU short-read, PacBio long-read, and ONT long-read cohorts showing the distribution of participants by sex, self-identified category, availability of electronic health records (EHRs), physical measurements, and survey responses. 1,027 participants self-identifying as Black or African American were selected from 245,388 AoU participants with short-read data. PacBio HiFi data was generated for all selected participants, with additional ONT R10.4 data generated to serve as validation data for putative variants.

### Variant discovery and harmonization

We identified small genomic variants (SNVs and indels <50 bp) using the DeepVariant deep learning-based variant caller^23^. All SVs (≥50 bp) were detected using an ensemble approach that combined *de novo* assembly-based discovery via Phased Assembly Variant Caller (PAV)^12^ with two read-based algorithms: Sniffles2 and PBSV^21,24^.

The ensemble algorithm approach improved sensitivity to diverse variant types, including those detectable by only a single method. However, integrating results from multiple tools, particularly at moderate (∼8x) coverage, poses major challenges for variant filtering and merging while attempting to distinguish true variants from false positives from LRS^25^. To address this and construct a unified SV callset across the cohort, we developed a custom harmonization pipeline, including filtering, normalization, and merging (Supplementary Fig. 1). Briefly, we clustered and normalized raw calls within each participant using Truvari^26^, selected for its high recall and minimal over- and under-merging when benchmarked against high-quality diploid assemblies from the Human Pangenome Reference Consortium (HPRC)^14^. We then trained a per-participant XGBoost^27^ classifier to filter likely false positives using genotyping features extracted with Kanpig^28^ (i.e., allele depth, allele balance, variant length, and multi-caller support). Training labels were derived from Truvari matches to dipcall^29^ variants on HPRC assemblies, enabling receiver operating characteristic (ROC) curve-based thresholding to approximate target true positive rates (TPRs). We applied thresholds to generate both lenient (TPR ≈ 0.9) and stringent (TPR ≈ 0.7) filtered callsets. Finally, filtered per-participant callsets were merged across the cohort using Truvari, and the resulting cohort-level VCF was regenotyped with Kanpig to maximize sensitivity and recover shared variants across participants (Supplementary Methods).

In our final variant discovery callsets, the number of SNVs was comparable between the PacBio LRS and Illumina srWGS platforms analyzed using 990 overlapping participants (Table 2). As expected, the number of indels was substantially reduced in the PacBio LRS as a consequence of the challenges of sequencing short tandem repeats on different sequencing platforms^17,21^. By contrast, there was a dramatic increase in SV discovery ranging from 60% to 290% depending on the stringency level applied to the putative SV callset before comparisons. This result is consistent with prior studies demonstrating the superior sensitivity of long-read technologies for SV detection^12,15,30^. We have therefore released two SV callsets—a lenient callset of 1,213,876 SVs maximized for sensitivity and a strict callset of 665,869 SVs optimized for high specificity. Both SV datasets include the HPRC controls and were subjected to similarly rigorous QC steps with all metrics provided (Fig. 2).

**Fig. 2:**
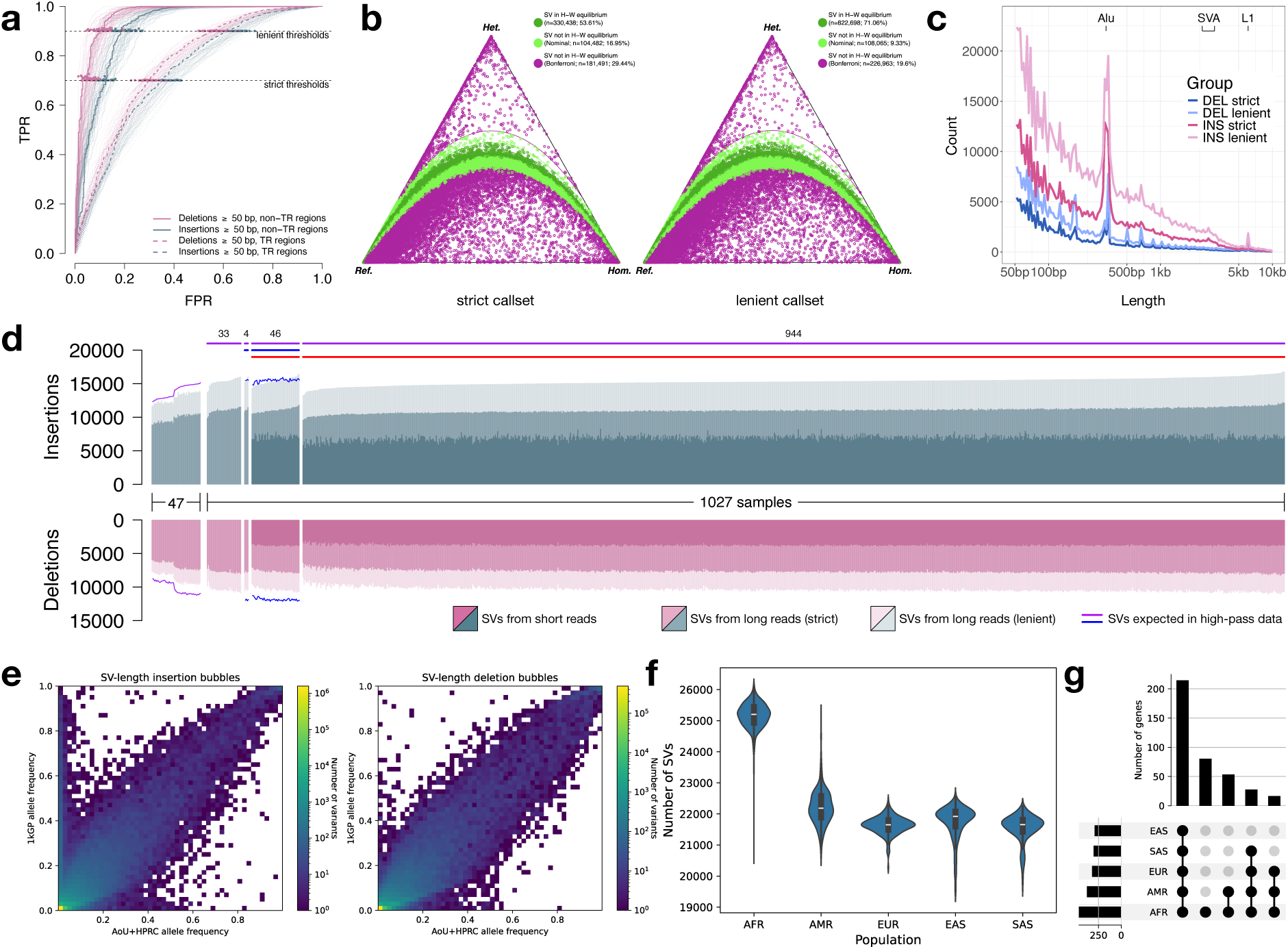
Structural variant calling and quality control. **a,** Receiver operating characteristic (ROC) curves for per-participant SV filtering performance, evaluated against 47 high-quality diploid assemblies from the HPRC. True positive rate (TPR) and false positive rate (FPR) are shown under strict and lenient filtering thresholds, stratified by variant type and genomic context (tandem repeat [TR] vs. non-TR regions). One participant is highlighted for clarity. **b,** De Finetti diagram depicting ratios of homozygous reference, heterozygous, and homozygous alternate genotypes at each SV site in strict (left, n=616,411 SVs) and lenient (right, n=1,157,726 SVs) callsets, as well as the fraction of SV sites in Hardy-Weinberg equilibrium. Every site is assumed to be biallelic. **c,** Distribution of SV lengths for insertions and deletions, stratified by callset. Peaks corresponding to known mobile element families (*Alu*, SVA, and L1) are labeled. **d,** Per-participant SV counts across different groups: 47 HPRC samples used for comparison, 1,027 AoU participants with long reads (subsets with other data types indicated by horizontal bars; PacBio: purple; ONT: blue; Illumina: red). Purple and blue curves represent expected variant counts based on raw Sniffles2 calls on ∼30x PacBio data from HPRC samples and ∼35x ONT data from AoU, respectively, illustrating expected SV yield at higher coverage. **e,** Histograms of unfiltered allele frequencies from the AoU+HPRC panel (x-axis) and 2,504 unrelated 1KGP short-read samples imputed against it (y-axis), for SV-length insertion/deletion (left/right, Pearson correlation coefficient 0.84/0.94) bubbles. **f,** Number of imputed AoU LRS SVs per participant across five continental population groups in 2,504 unrelated 1KGP short-read samples. **g,** UpSet plots show the number of protein-coding genes (including intronic loci) intersected by an SV across the five continental groups in 1KGP.

**Table 2.** Discovered variation in CDRv7 AoU Long-Read dataset.

|  | PacBio |  | PacBio (srWGS subset) |  | Illumina (with SVs) |
| --- | --- | --- | --- | --- | --- |
| Sequencing platform | Sequel IIe |  | Sequel IIe |  | NovaSeq 6000 |
| Participants | 1,027 / 245,394 |  | 990 / 1,027 |  | 990 / 1,027 |
| SNVs | 66,467,539 |  | 65,541,649 |  | 62,895,482 |
| Ti/Tv ratio | 1.908 ± 0.015 |  | 1.907 ± 0.012 |  | 2.010 ± 0.002 |
| Het/hom ratio | 1.924 ± 0.141 |  | 1.923 ± 0.138 |  | 2.184 ± 0.102 |
| indels (<50 bp) |  |  |  |  |  |
| insertions | 3,212,616 |  | 3,045,864 |  | 6,073,915 |
| deletions | 4,714,659 |  | 4,072,836 |  | 3,161,161 |
| SVs (≥50 bp) | strict callset | lenient callset | strict callset | lenient callset |  |
| insertions | 499,272 | 934,491 | 469,592 | 837,975 | 117,312 |
| deletions | 166,597 | 279,385 | 157,945 | 254,765 | 71,338 |

Variant discovery (SNVs, small indels, and SVs) in the full 1,027 AoU long-read cohort and a subset of 990 participants overlapping available short-read SV calls for AoU participants. For SVs, counts from strict and lenient callsets are shown.

### Evaluation and quality control

We applied a series of quality controls to assess the accuracy of both the lenient and strict SV callset. First, to evaluate Mendelian consistency as a function of sequence coverage, we selected three trios with high-coverage HiFi sequencing data^11^. Each participant’s LRS data were downsampled to 8x, 16x, and 25x coverage using GATK DownsampleSam^31^. For each coverage level for each participant, we applied our full pipeline, merging and regenotyping separately for each coverage level. Using the GRCh38 reference, we found a mean Mendelian discordance rate of 6.4% for the TPR=0.9 leniently filtered callset at 8x coverage, with the rate dropping to 3% at 25x coverage (Supplementary Fig. 2). Notably, this discordance rate from our analysis of 8x coverage data was consistent to that reported cohort-wide by the Human Genome Structural Variation Consortium (HGSVC) using high-coverage LRS (6%), further validating the utility of the mid-pass approach^12^. The strictly filtered TPR=0.7 callset had a similar discordance rate of 6.8% at 8x coverage, dropping to 2.9% at 25x coverage. Discordance rates were similar but slightly elevated on the T2T-CHM13 reference (Supplementary Fig. 3). Both references exhibited substantially improved concordance rates for variants outside of tandem repeats (Supplementary Fig. 4).

Both datasets show the expected SV size distribution with notable peaks at 300 bp and 6 kbp, consistent with SINE and LINE mobile element insertions (MEIs)^12^ as well as an increasing discovery curve for insertions and deletions (Fig. 2c,d) consistent with expectations. After collapsing MEIs at the same locus, we detected a total of 28,101 (24,663) *Alu*, 3,760 (2,873) LINE-1, and 1,750 (1,253) SVA insertions in the lenient (and strict) callsets. Per participant, we detected a median of 1,620 (1,500) *Alu*, 166 (140) LINE-1, and 83 (68) SVA insertions in the lenient (and strict) callsets (Supplementary Fig. 5a), with expected length distributions for each class (Supplementary Fig. 5b). For comparison, using high-coverage LRS, the HGSVC^11^ detected 1,585 *Alu*, 212 LINE-1, and 98 SVA insertions per participant of African genetic ancestry. We observed extensive allelic heterogeneity in SVA length (Supplementary Fig. 5c,d), with the top-most variable SVAs having length differences exceeding 2 kbp across participants. These length differences were predominantly driven by copy number changes in the central CG-rich variable number tandem repeat (VNTR) region (Supplementary Fig. 5d).

As a final validation approach to assess precision, we selected 50 AoU participants that overlapped with the PacBio data analysis and sequenced them with ONT (Fig. 1, Table 1). We utilized Kanpig^28^ to genotype the HiFi-based SV calls on GRCh38 and T2T-CHM13. We observe high support for the lenient SV callset with 95.5% precision for GRCh38 and 90.5% precision for T2T-CHM13. Subsequent genotyping (see below) against the 1KGP samples confirmed that the majority of common SVs are observed among all population groups, but that SVs specific to African genetic ancestry are the second most abundant (Fig. 2e-g).

### Creation of an LRS reference panel for srWGS genotyping, phasing, and imputation

To build a phased, haplotype-resolved reference panel optimized for SV imputation into srWGS data, we integrated the AoU and HPRC LRS callsets (spanning SNVs, small indels, and SVs) using the stringent (TPR=0.7) SV callset to minimize false positives. We first performed read-based physical phasing of both callsets jointly using HiPhase^32^, followed by postprocessing to remove (1) singleton variants, (2) duplicate variants between SV and SNV callsets, (3) short variants with AF < 0.5% and >25 kbp from any SV, and (4) positionally conflicting variant calls yielding inconsistent haplotypes. We then applied SHAPEIT4 for statistical phasing and imputation and postprocessed the phased haplotypes by concatenating adjacent alleles to define non-overlapping “bubble” variant sites, generating a pangenome graph representation (see Supplementary Methods for further details)^33^. Finally, using the KAGE^34^ genotyping and GLIMPSE^35^ phasing and imputation methods, we called variants in this LRS-derived reference panel from srWGS case participants.

The resulting phased, imputed, and haplotype-resolved SNV/indel/SV reference panel of 1,074 AoU+HPRC participants covers the autosomal chromosomes and consists of 19,942,647 variant sites in bubble representation, which contain 30,918,204 variant bubble alleles; 189,382 of these bubbles are SV-length and 74,550 are further multiallelic. For comparison, the 44-sample HPRC panel contains 21,304,582 (15,684,910) bubble sites in the autosomes, of which 71,873 are SV-length and 55,156 are further multiallelic. After decomposing bubble alleles back to their original representations, the panel contains 27,143,444 distinct variant alleles, including 770,473 SVs (653,740 insertions and 117,698 deletions).

Both LRS-panel and srWGS-case pipelines were validated using HPRC assembly-based dipcall truth with the vcfdist alignment-based benchmarking method^36^. Performing a leave-one-out experiment with 40 HPRC samples on a single evaluation chromosome, the LRS panel (unfiltered srWGS cases) achieved 94.0 ± 1.1% (95.5 ± 1.0%) precision and 91.4 ± 1.3% (87.9 ± 2.0%) recall for SVs with length <5 kbp in dipcall-confident, non-repeat/homopolymer regions. This accuracy is comparable across both pipelines, which demonstrates the additional power that prior knowledge derived from LRS resources can unlock in readily available srWGS data. Further details of the panel creation and case pipelines, as well as our validation procedures, are provided in the Supplementary Methods.

### Characterization of SVs enriched in African genetic ancestry

Using our AoU+HPRC reference panel, we genotyped and imputed variants in srWGS data using all 3,202 individuals from the 1KGP^37^, generating a comprehensive SV callset across five continental populations (Fig. 2f). 1KGP samples with African genetic ancestry showed the highest number of SVs per individual compared to other groups, consistent with previous findings of greater genetic diversity among individuals of African genetic ancestry^38^. A PCA of the SV callset achieved clear population stratification, closely matching patterns observed in the published 1KGP SNV dataset^39^ and supporting the reliability of our SV genotypes for population genetic analyses (Supplementary Fig. 6a,b).

We next intersected SVs with coding regions of genes and regulatory elements (Figs. 2g, Supplementary Fig. 6c, Supplementary Table 1). The majority of SV-overlapping elements (40.3% of coding; 43.7% of regulatory) were shared across all populations, indicating that our panel captures globally common variation. The second-largest category included elements specific to African genetic ancestry (15.1% coding; 17.1% regulatory), reflecting both the elevated diversity within genomes of African genetic ancestry and their strong representation in the AoU cohort.

The majority of the imputed SVs (315,307 of 541,049; 58.3%) had higher allele frequencies in African genetic ancestry than in non-African genetic ancestry. To further assess population differentiation, we calculated the fixation index (F_st_) between African and non-African groups and identified 8,249 SVs with F_st_ ≥ 0.15, indicating strong enrichment in African genetic ancestry (Supplementary Fig. 6d; Supplementary Table 1). These variants were distributed across the genome and intersected with 776 high-priority disease-associated genes (Supplementary Fig. 6e). One notable example is a 61 bp deletion in *BARD1* (African ancestry AF = 0.12; non-African ancestry AF = 0.002; F_st_ = 0.19; Supplementary Fig. 6f), a gene linked to elevated breast cancer risk^40^. We also observed SVs specific to African genetic ancestry in genes associated with conditions disproportionately affecting individuals of African ancestry, such as *APOB* and *MYH7*, both of which are implicated in cardiovascular disease^41^ (Supplementary Table 1).

### Characterization of clinically relevant and population-enriched SVs

Because this dataset represents the largest LRS survey to date of participants self-identifying as Black or African American, we conducted an in-depth analysis of SVs, which have greater potential for novel allelic variant discovery and outsized impact on health^42^. We first investigated known disease-associated loci (such as those involving triplet repeat instability and complex pharmacogenomic regions like *CYP2D6*) to assess the added value of LRS in identifying novel variants of potential clinical significance. Second, we focused on SVs that are enriched in groups self-identifying as Black or African American and absent from existing variant catalogs. These variants, more likely to have larger effect sizes, were tested for association with gene expression and disease. Together, these analyses provide a valuable resource for understanding genetic diversity and improving health outcomes in populations that continue to face structural barriers to receiving high-quality tailored healthcare.

### Tandem repeat loci and disease

Pathogenic expansions of short tandem repeats (STRs) have been implicated in over 50 inherited diseases^43^, where both tract length and purity of the repeat influence instability. However, many of these disease-associated loci have been difficult to fully characterize by srWGS or PCR due to their size or GC-rich content. To address this, we performed detailed characterization of two well-known triplet repeat loci: the CGG repeat in the 5’ untranslated region (UTR) of *FMR1*, associated with Fragile X syndrome (FXS) (Fig. 3a,b), and the CAG repeat in the coding region of *HTT*, associated with Huntington’s disease (Fig. 3d,e). Repeat sequences were derived with TRGT^44^ for a total of 1,753 (sex-filtered) haplotypes at *FMR1* and 1,944 at *HTT*. In both cases, we observed the most common haplotypes reported in the literature, including characteristic AGG interruptions occurring every 9-10 CGG units within the *FMR1* repeat (Fig. 3a,d)^45,46^.

**Fig. 3:**
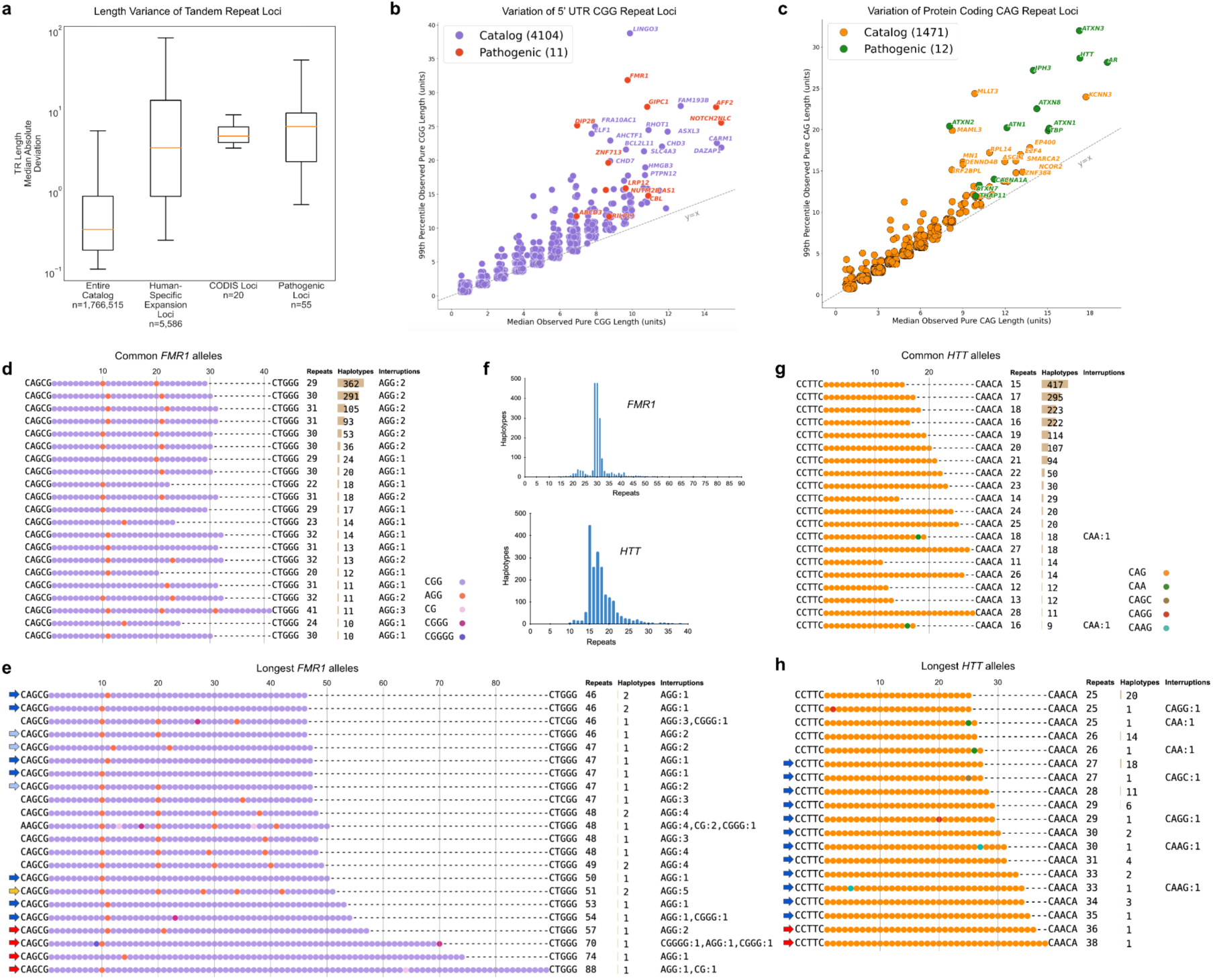
Expanded triplet repeat loci and structure of disease-associated loci. **a,** Most common and **b,** longest *FMR1* repeat alleles. Sequences were extracted from 1,753 sex-filtered long-read haplotypes with TRGT and plotted according to repeat and interruption patterns. AGG interruptions are included in repeat counts. Arrows indicate unstable transmission risk alleles with 25 to 33 (light blue) or at least 34 (dark blue) uninterrupted CGG repeats, premutation alleles with at least 55 repeats (red), and an allele with five interruptions (yellow). **c,** Length distributions indicating the number of repeat units in extracted TRGT sequences for *FMR1* and *HTT*. **d,** Most common and **e,** longest *HTT* repeat alleles. Sequences were extracted from 1,944 long-read haplotypes with TRGT and plotted. Blue arrows indicate intermediate-length unstable transmission risk alleles with 27 to 35 CAG repeats, and red arrows indicate potential low-penetrance Huntington’s disease alleles with at least 36 repeats. **f,** Boxplot of the distribution of tandem repeat length variation values (measured by median absolute deviation) for the entire catalog (first box), a set of loci specifically expanded in humans relative to nonhuman primates (second box), the CODIS loci (third box), and the known pathogenic loci (fourth box). **g,** 5’ UTR CGG repeats and **h,** protein-coding CAG repeats with median (50^th^ percentile) repeat length for each locus plotted against the 99^th^ percentile repeat length, both measured in trinucleotide units. At each locus, the longest pure (uninterrupted) repeat was segmented. A y=x trend line is included for reference, indicating where the median and 99^th^ percentile repeat lengths are equal. Out of 4,115 CGG loci and 1,483 CAG loci, 11 and 12 disease-associated loci are highlighted in red/green with annotated genes, while non-disease-associated loci are shown in purple/orange.

Because expanded alleles are associated with disease/disorder risk, we specifically focused on the tail-end of each length distribution (Fig. 3b,e). In *FMR1*, we predicted ten of the longest alleles to be potentially unstable due to the loss of the 3’ AGG interruption, extending the length of the longest pure CGG repeat tract^47,48^. We also identified four premutation-sized alleles (≥55 repeat units), of which the longest occurred in a participant for whom we estimate an ∼83-93% probability of expansion to a full mutation in offspring on maternal transmission. This estimate was phenotypically supported by the participant’s EHR, which describes a child with symptoms commonly associated with FXS.^49,50^. In *HTT*, we observed 12 potentially unstable intermediate-length alleles and two reduced-penetrance alleles (≥36 CAG repeat units), corresponding to a carrier allele frequency of ∼0.1% (2/1,944 haplotypes). We identified several noncanonical and rare interruptions in the *FMR1* and *HTT* repeats (Supplementary Note 1). In addition, we identified a participant with an *ATXN3* repeat expansion exceeding the pathogenic, high-penetrance threshold of >55 CAGs for spinocerebellar ataxia type 3 (Supplementary Fig. 7) with a diagnosis of cerebellar ataxia recorded in their EHR, supporting this genotype^51^.

To identify potentially novel loci associated with triplet repeat instability, we applied a catalog-driven approach^52^ to GRCh38-aligned participants and identified several classes of tandem repeats exhibiting significantly greater length variability than the catalog as a whole (Fig. 3f). These included loci known to be expanded in humans relative to nonhuman primates, autosomal CODIS loci used in forensic analyses, and most recently described pathogenic tandem repeats^53–56^. We further characterized this set by plotting the 99^th^ percentile length of the longest pure repeat segment against the median (50^th^ percentile) longest pure repeat segment length for every 5’-UTR CGG repeat (Fig. 3g) and every coding-region CAG repeat (Fig. 3h) in the catalog. Consistent with the initial observation, known pathogenic repeats for both groups clearly segregate from the majority of loci, occupying the right tail of the length variability distribution. In addition to these known loci, we identified 13 CAG repeat loci and 15 CGG repeat loci that exhibit similar distributions of variation to the known pathogenic loci. We hypothesize that these loci represent strong candidates for further study as potentially pathogenic STRs. A companion manuscript further explores these observations and candidates^57^.

### CYP2D6 variation

Given the crucial role of CYP2D6 in the metabolism of commonly prescribed medications and the known difficulty of resolving this locus using srWGS due to complex structural variation^58,59^, we evaluated whether LRS assemblies could provide higher-resolution genotyping. We first developed a k-mer-based method to determine the identity and copy number of *CYP2D6* and its paralog *CYP2D7* (Methods) and applied it to 1,266 AoU haplotypes (62%) contiguously assembled over the full *CYP2D6-7* region, following HPRC validation. This analysis revealed 13 distinct structural configurations (Fig. 4a), including 38 *CYP2D6* duplications, 87 full-gene deletions, and 18 hybrid alleles corresponding to four structures: *CYP2D6-CYP2D7::6* (n=6), *CYP2D6-CYP2D7::6-CYP2D7* (n=5), *CYP2D6::7* (n=4), and *CYP2D7::6-CYP2D7* (n=3). We next identified star alleles, defined as haplotypes associated with enzymatic activity levels. Consensus calls from four validated short-read genotypers supported duplications for 28/33 participants and deletions for 56/75 participants, and classified seven hybrid alleles as *68+*4. Among the five duplications with discordant calls, three had indeterminate short-read consensus, and the Pangu genotyper from PacBio-supported duplications in three cases^60^. Among 19 deletion discrepancies, one was indeterminate by srWGS, and Pangu called the *5 full-gene deletion allele in one participant and reported only a single haplotype in 14 others. srWGS calls identified four of ten hybrid discrepancies as potential *4 hybrids, while Pangu called three as the *4.013 exon 9 conversion.

**Fig. 4:**
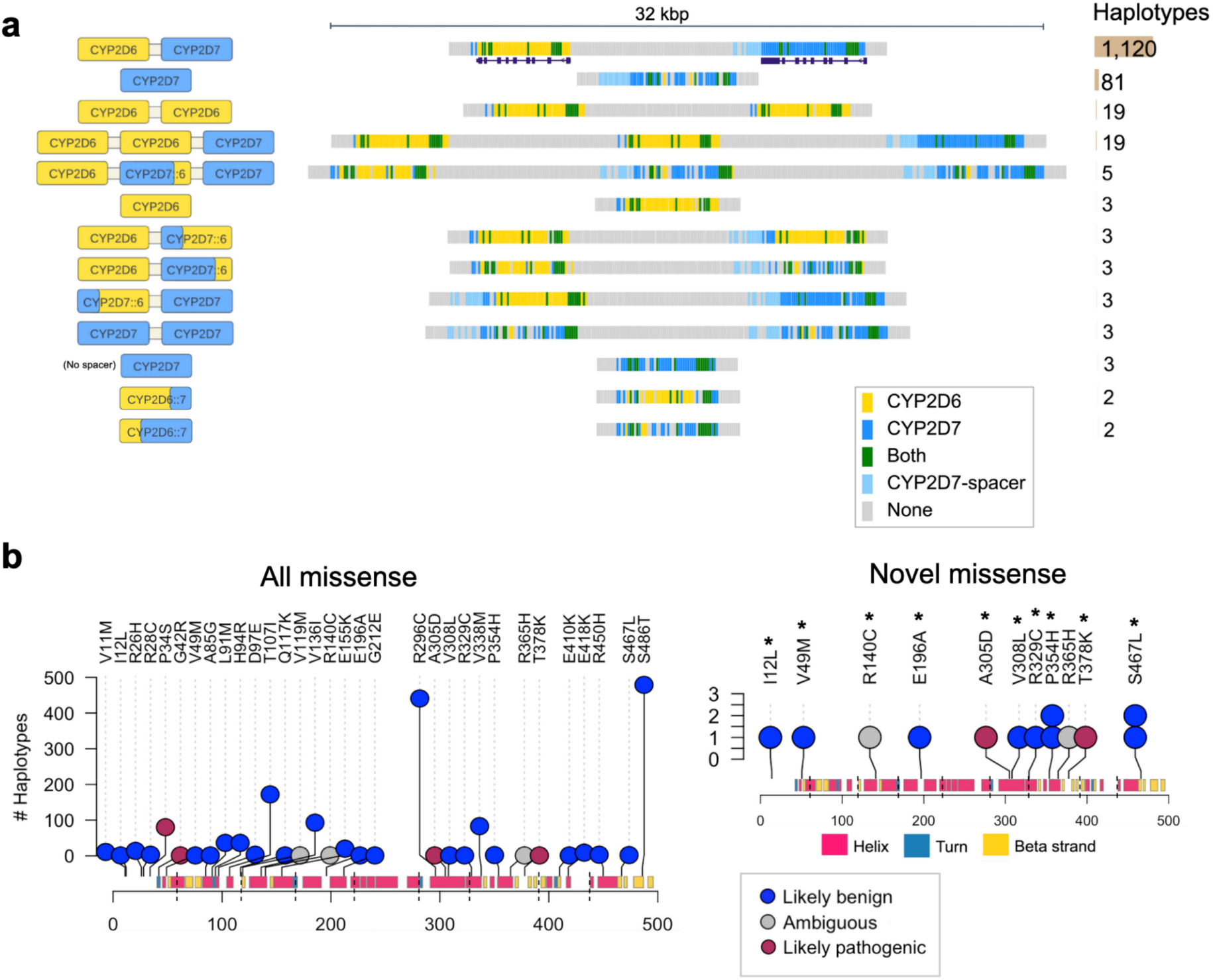
*CYP2D6-7* haplotype structure and content. **a,** Copy number variation and hybrid structures for 1,266 AoU-LR Phase 1 haplotypes. Hifiasm assemblies with a single contig over the full *CYP2D6*-*7* locus were divided into 100 bp k-mers and colored by the best-mapping reference gene. An example plot for each structure is shown along with the number of haplotypes the structure was seen in. **b,** All (left) and novel (right) missense variants. Predicted haplotype coding sequences were compared with the GRCh38 coding sequence, which was substituted with changes for the assigned (sub)star allele to find novel variants. Variants are colored by AlphaMissense classification, and asterisks indicate presence in the srWGS and LRS AoU Hail matrix tables.

We developed an LRS star-allele inference pipeline that selects the closest-matching reference allele by maximizing variant matches and alignment score. Applying this to the AoU LRS assemblies, we observed 91% concordance with srWGS star allele calls. The most frequent unambiguous disagreements involved differences in copy number or hybrid structure (Supplementary Table 2). We restricted our analysis to 255 participants with a canonical *CYP2D6-7* structure, for which diplotypes (and thus metabolizer status) could be fully inferred. Among these, 172 were categorized as CYP2D6 normal metabolizers, 77 as CYP2D6 intermediate metabolizers, and 6 as CYP2D6 poor metabolizers. To examine novel mutations beyond the SNVs and indels found in the closest-matching star allele, we predicted the coding and protein sequence for each haplotype and compared them to the reference translation. Among 861 haplotypes with unambiguous assignments and a single predicted transcript, 13 carried missense variants not present in the corresponding suballele reference. Of these, two were classified by AlphaMissense as likely pathogenic and two as having ambiguous pathogenicity (Fig. 4b)^61^. Only one of these four variants was also supported by the srWGS data.

### Functional and clinical interpretation of AoU SVs

Using the stringent SV callset, we evaluated the ability of the LRS dataset to detect SVs in other biologically and clinically significant regions. We identified 251,723 SVs mapped to protein-coding loci, including intronic regions. Among these, 68.2% of deletions and 63.4% of insertions were not detected by srWGS, yet intersected 8,219 OMIM genes^62^, 2,801 genes prioritized for inherited diseases^63^, and 49 from the American College of Medical Genetics and Genomics (ACMG) dataset^41^, suggesting the considerable value of LRS data for improving the discovery of medically relevant variants (Supplementary Table 3). LRS also enabled the resolution of SVs in complex, highly polymorphic regions. For example, 723 LRS-unique SVs were identified within the major histocompatibility complex, including an 816 bp deletion within *HLA-A* (Supplementary Fig. 8a).

We then prioritized the stringent SVs using CADD-SV^64^, which classified over 99% (574,531 SVs) as benign (Fig. 5a). However, 473 SVs received PHRED-scaled scores ≥20, placing them among the top 1% of predicted deleterious variants relative to a healthy population reference (Supplementary Table 3). As expected, most putatively deleterious SVs were rare within the AoU cohort (Fig. 5a, Supplementary Fig. 8b), particularly those with scores ≥30, consistent with purifying selection acting against highly damaging variants^5,65,66^. We further compared the AoU long-read SVs with recently published LRS datasets, including the 1KGP ONT callset, HSGVC, and HPRC^12,14,67^. SVs absent from these external resources generally had a lower allele frequencies in the AoU cohort than shared SVs (Fig. 5a, Supplementary Fig. 8b), suggesting they are both rare and potentially population-specific. Among 273 high-priority, previously unreported SVs, 172 overlapped 170 medically relevant genes^62^, 73 intersected 69 high-priority disease-related genes^63^, and 15 affected 14 genes associated with cancer risk^68^ (Supplementary Table 3). These SVs represent compelling candidates for studying disease risk, especially in individuals of African genetic ancestry.

**Fig. 5:**
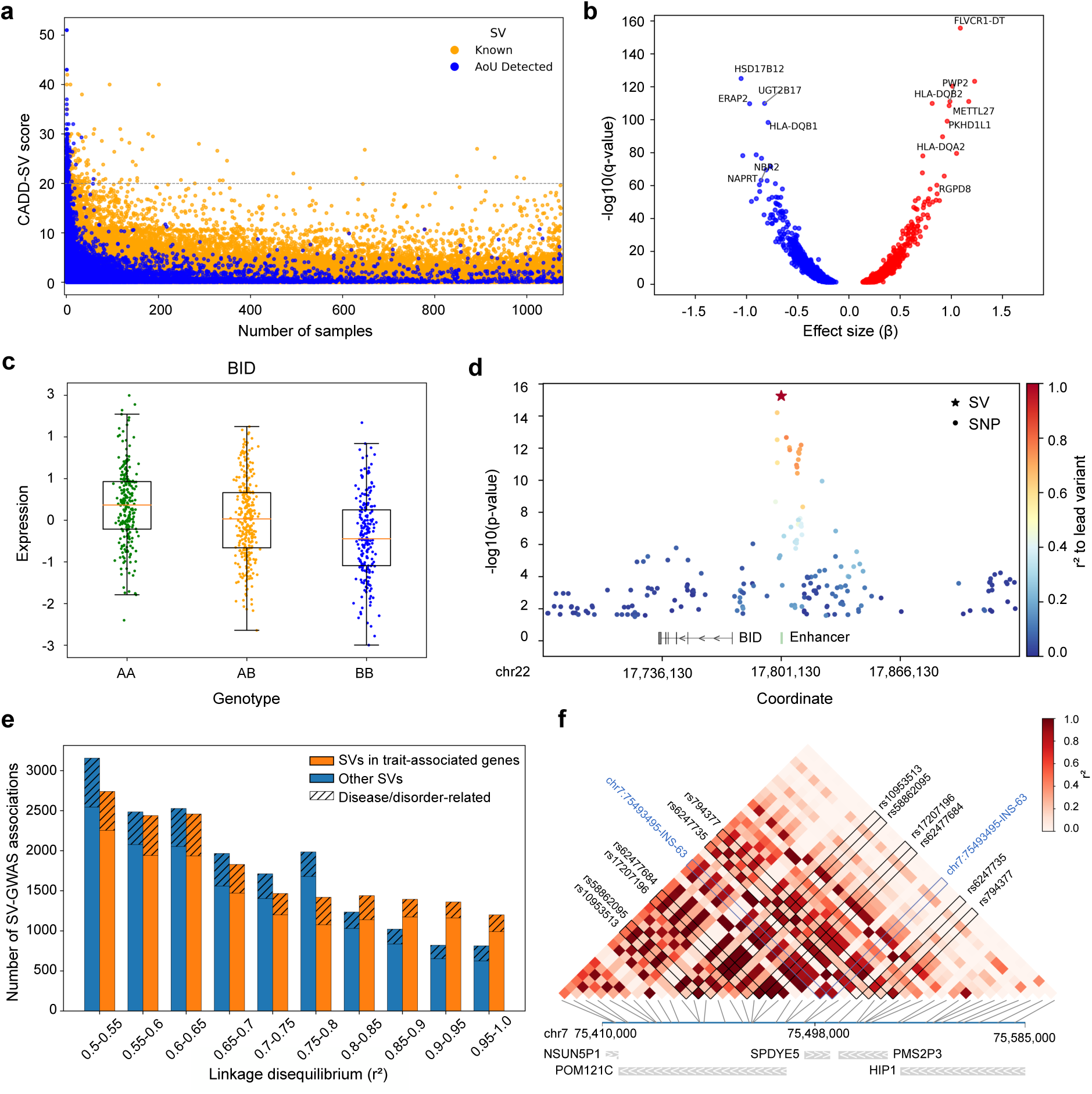
Functional impact of AoU SVs. **a,** Distribution of CADD-SV score for shared SVs in the AoU strict cohort. Each data point represents an SV. The x-axis indicates the number of participant containing the SV, and the y-axis shows the PHRED-scaled CADD score. “Known” SVs are those identified in at least one of the 1KGP-ONT, HGSVC, or HPRC datasets^12,14,67^, while “AoU detected” SVs are those absent in those datasets. The dashed gray horizontal line denotes the score threshold above which SVs are considered likely pathogenic. **b,** Relationship between the best-fit slope (*β*) derived from OLS regression and gene-level q-values. The eGenes shown are medically significant and listed in OMIM^62^, with high-confidence q-values (<1 × 10^-60^). **c,** Distribution of genotypes and gene expression values in 731 participants for the *BID*-associated deletion, with a q-value of 5.92 × 10^-13^. **d,** Manhattan plot of the 322 bp deletion and nearby SNVs, with log_10_ P values. The deletion is the top variant, and points are colored by their LD (r^2^) with this SV. The enhancer overlapping the SV is shown in green. **e**, Histogram showing the number of SVs in LD (r^2^ ≥ 0.5) with SNVs from the GWAS catalog. SVs located within trait-associated genes are shown in orange, and those outside these genes are shown in blue. Bars with diagonal hatching (///) indicate SVs linked to disease- or disorder-related traits. **f,** LD heatmap of an insertion (marked in blue) along with nearby GWAS variants and SVs located within ±100 kbp of the insertion. GWAS variants in LD are labeled with their IDs. GWAS-associated genes are shown below the heatmap.

We then leveraged matched EHR data to explore the clinical outcomes of participants carrying rare, high-impact SVs not found in previous LRS datasets. For instance, this highlighted a 52 bp insertion in *SLC2A12* (CADD-SV score: 51) found in a participant diagnosed with Type 2 diabetes and hyperglycemia (Supplementary Fig. 8c, Supplementary Tables 3 and 4). As *SLC2A12* encodes GLUT12, a glucose transporter implicated in metabolic regulation^69,70^, this suggests a potential link to disease. In addition, a 536 bp insertion in *TOX* (CADD-SV score: 43), a gene involved in asthma pathogenesis^71^, was found in a participant with asthma (Supplementary Fig. 8d, Supplementary Tables 3 and 4). Several high-impact SVs were predicted to disrupt protein structure. One example is a 226 bp insertion in *PSIP1* (CADD-SV score: 31), associated with hearing loss^72^, which introduced a premature stop codon and was predicted by AlphaFold to truncate the protein (Supplementary Fig. 8e,f, Supplementary Tables 3 and 4)^73^. Notably, the participant carrying this variant presented with sensorineural hearing loss. In another case, a 4,025 bp deletion (CADD-SV score: 21.3) removed three exons of *TARS2* in a participant with chronic kidney disease, consistent with prior studies linking *TARS2* mutations to this condition^74^ (Supplementary Fig. 8g,h, Supplementary Tables 3 and 4).

### Measuring effects of SVs on gene expression and complex traits

To evaluate the regulatory impact of SVs, we performed expression quantitative trait loci (eQTL) mapping using the imputed SV callset from 1KGP and paired gene expression data from the MAGE dataset^75^. This analysis identified 3,835 significant SV-eQTLs (q-value < 0.05), involving 2,511 medically relevant OMIM genes, 747 high-priority disease genes, and 117 cancer-associated genes, highlighting potential mechanisms through which SVs contribute to disease risk (Fig. 5b, Supplementary Table 1). Notably, 889 of the eGenes were absent from two earlier long-read SV-eQTL analyses involving 31 diverse long-read genomes and 65 samples from the 1KGP-ONT dataset^67,76^. The majority (84.6%) of SVs in SV-eQTL pairs were common across all populations (AF ≥ 0.05), allowing robust cross-population analysis of regulatory effects (Supplementary Fig. 9a). Consistent with the AoU LRS reference panel was enriched for participants genetically similar to African reference populations, we further identified 34 statistically significant SV-eQTLs in which the SVs were common in individuals of African genetic ancestry (AF ≥ 0.05) but rare in others (AF < 0.05), indicating newly identified regulatory factors associated with African genetic ancestry (Supplementary Table 1). For example, a 512 bp insertion (AFR AF=0.56; Non-AFR AF=0.02, F_st_=0.65) was associated with expression changes in *SHPK*, a gene involved in sedoheptulokinase deficiency and broader metabolic regulation (Supplementary Fig. 9b-c, Supplementary Tables 1 and 5).^77,78^

To identify SVs with likely causal effects on gene expression, we applied the fine-mapping tool CAVIAR^79^ to quantify the posterior probability of causality while accounting for linkage disequilibrium (LD) with SNVs within a 1 Mbp window. This analysis identified 144 SV-eQTLs in which the SV had the highest CAVIAR score compared to the flanking SNVs for the eGene. One notable example is a 322 bp deletion within an enhancer upstream of *BID*, a pro-apoptotic BCL-2 family member involved in cancer progression and immune regulation^80,81^. SV carriers showed decreased *BID* expression, consistent with the expectation that disruption of enhancer activity would alter gene expression (Fig. 5c, Supplementary Fig. 9d, Supplementary Tables 1 and 5).

This deletion is the lead variant, with a 386-fold stronger statistical significance than the strongest nearby SNP with a concordant direction of effect, indicating the importance of SVs in gene regulation (Fig. 5d, Supplementary Fig. 9e).

### SVs in LD with GWAS-significant SNVs

To explore the relationship between SVs and complex traits, we intersected imputed SVs with the NHGRI-EBI GWAS Catalog^82^, a comprehensive repository of genome-wide association study (GWAS) summary statistics. We identified SNV-SV pairs in moderate LD (r^2^ ≥ 0.5) where the SNV had genome-wide significance. This analysis revealed 6,019 traits associated with 5,549 SVs, including 1,103 traits related to diseases or disorders (Fig. 5e; Supplementary Fig. 10a, Supplementary Table 1). For instance, a 321 bp insertion in *TSPAN8*, a negative regulator of airway mucin secretion^83^, showed high LD with the asthma-associated rs11178649 (*β* = −0.04, p = 1 × 10^-10^, r^2^ = 0.69; Supplementary Fig. 10b). Using matched EHRs from our AoU cohort, carriers of this SV had reduced asthma risk (odds ratio [OR] = 0.46, 95% confidence interval [CI]: 0.25−0.87, p = 0.01; Supplementary Tables 1 and 5), supporting a potential protective role.

We next evaluated 677 SVs in high LD (r^2^ ≥ 0.8) with SNVs implicated in 537 medically relevant traits. One example is a 1,375 bp deletion in LD with rs16856925, an SNV linked to breast cancer risk (OR = 1.19, p = 7 × 10^-10^, r^2^ = 0.93; Supplementary Fig. 10c). This SV was also significantly associated with increased breast cancer risk in the AoU cohort (OR = 2.91, 95% CI: 1.21−7.01, p = 0.02; Supplementary Tables 1 and 5). Furthermore, some SVs exhibited potential pleiotropic effects, a phenomenon previously observed in a multi-trait SNP-based PheWAS^84^. For instance, a 63 bp insertion at position 75,493,495 on chromosome 7 was in strong LD (maximum r^2^ = 0.88) with six GWAS SNVs (rs10953513, rs17207196, rs58862095, rs62477684, rs62477735, rs794377) associated with cardiovascular and metabolic traits, including atrial fibrillation, hypertension, obesity, and metabolic syndrome (Fig. 5f, Supplementary Fig. 10d). Interestingly, the EHR data from SV carriers revealed elevated risks of hypercholesterolemia (OR = 4.73, 95% CI: 1.48−15.07, p = 0.009) and ventricular tachycardia (OR = 3.35, 95% CI: 1.07−10.52, p = 0.039; Supplementary Tables 1 and 5), which are closely related to the GWAS-identified traits, suggesting this SV may contribute broadly to cardiometabolic disease.

### Phenome-wide SV analysis identifies disease risk factors

To further explore the clinical impact of SVs, we expanded our analysis to the srWGS and EHRs of 10,000 self-identified Black or African American participants in the *All of Us* cohort (CDRV7) (Supplementary Fig. 11a). For this, we performed genotyping and imputation based on the long-read-derived AoU reference panel, along with additional quality controls to exclude low confidence variants (Methods, Supplementary Fig. 11b-e). Using these results, we performed a phenome-wide association study (PheWAS) and identified 291 SV-disease associations (p < 1 × 10^-5^) across the genome spanning 226 distinct conditions. (Fig. 6a-c; Supplementary Fig. 12; Supplementary Table 6). Of these associations, 148 (50.9%) involved SVs absent from the matched short-read callset derived from 990/1,027 of the same participants, indicating the importance of LRS in capturing clinically relevant variants.

**Fig. 6:**
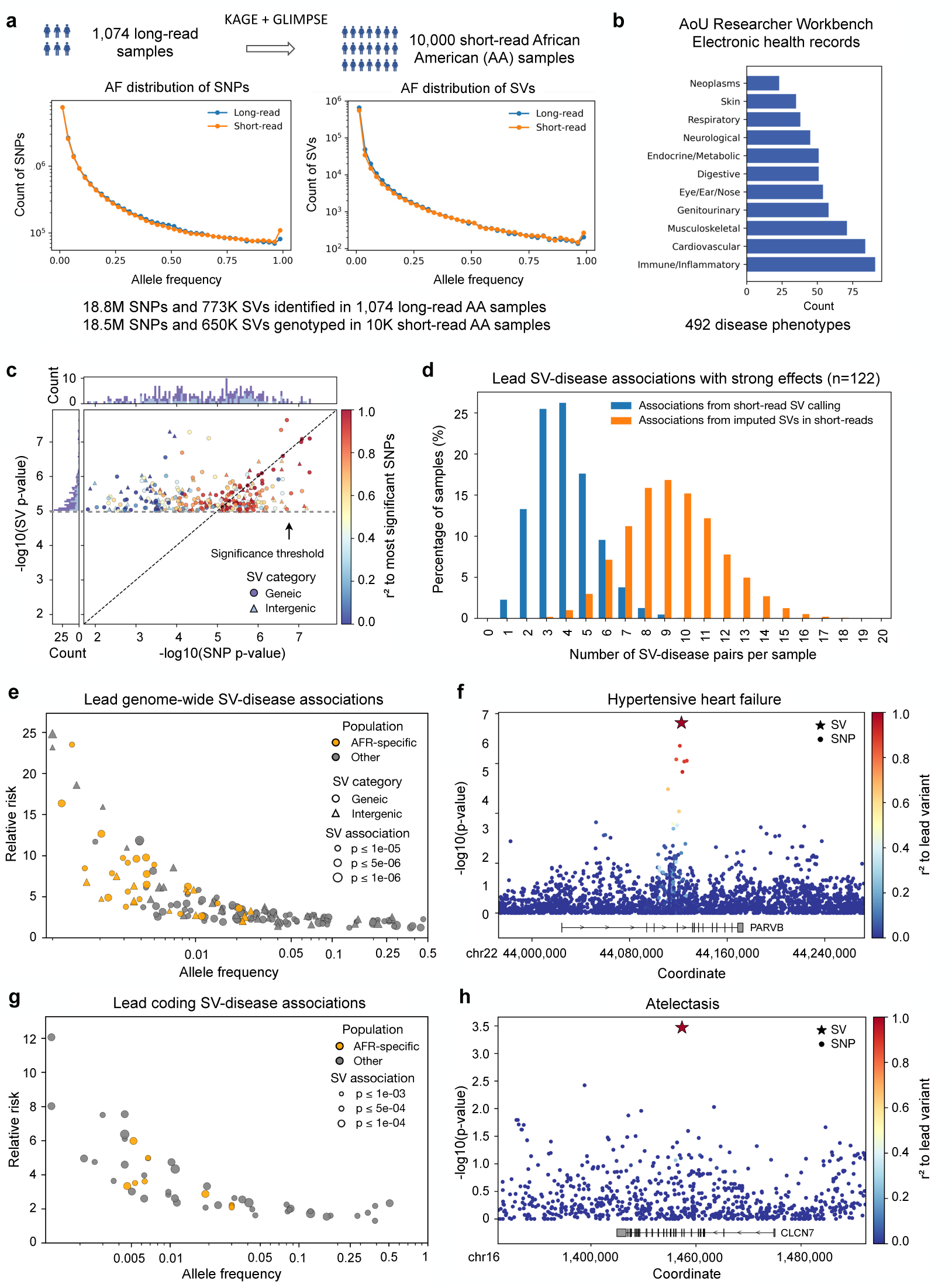
Genome-wide linkage and trait associations of SVs. **a**, Genotyping and imputation with the long-read panel. Small variants and SVs are identified from 1,074 long-read participants (AoU + HPRC) and then genotyped and imputed in 10,000 short-read self-identified Black or African American participants from *All of Us* biobank, with the total number and allele frequency distributions of SNVs and SVs shown. **b**, Disease phenotypes are extracted from EHRs of the same participants and grouped into 11 categories. Conditions belonging to multiple categories are assigned to all relevant groups. **c**, Comparison of association significance between each SV and the strongest nearby SNV (within ±100 kbp) for the same phenotype (central panel). Points are colored by LD (r^2^) between the SV and SNP. Circles indicate genic SVs (overlapping gene bodies), and triangles indicate intergenic SVs. The top and right panels display stacked histograms of −log_10_(p-values) for SNP- and SV-disease associations, respectively. Genic SVs are shown in purple; intergenic SVs are shown in light blue. **d,** Distribution of genome-wide SV-disease associations with large effect sizes (odds ratio [OR] ≥ 2.5 or ≤ 0.4) in which the SV is the lead variant. Orange indicates associations with SVs discovered in 1,027 AoU and 47 HPRC long-read samples and subsequently imputed in 10,000 AoU short-read participants, while blue represents associations with SVs identified in the short-read callset derived from the same 1,027 AoU participants. **e**, Relationship between allele frequency (log scale) and relative risk (OR > 1) for genome-wide disease-associated SVs with stronger signals than nearby SNPs. Circles indicate SVs within gene bodies, and triangles indicate intergenic SVs. Associations specific to African genetic ancestry are shown in yellow and all others in gray. Point size reflects association significance. **f,** Manhattan plots showing a 50bp deletion linked to hypertensive heart failure. The insertion is the lead variant, and surrounding points are colored based on their LD (r^2^) with the SV. **g**, Relationship between allele frequency (log scale) and relative risk (OR > 1) for coding-region disease-associated SVs with stronger signals than nearby SNPs. Associations specific to African genetic ancestry are shown in yellow and all others in gray. Point size reflects association significance. **h,** Manhattan plots showing a 200 bp insertion associated with atelectasis. The insertion is the lead variant, and surrounding points are colored based on their LD (r^2^) with the SV.

Our initial set of associations used a screening threshold of significance so that we could compare the relative significance of SNVs and SV across a broad range of traits. To prioritize likely causal variants, we conducted fine-mapping to estimate the posterior probability that each SV contributed to the disease risk, accounting for LD with nearby SNVs within ±100 kbp. This analysis identified 191 (65.6%) SV-disease pairs across 160 traits where the SV was the most likely causal variant within the locus (Fig. 6c, Supplementary Table 6). On average, each participant carried 9 strong associations (OR ≥ 2.5 or ≤ 0.4) in which the SV was the lead variant, compared with 4 in matched short-read SV callset, indicating the important contribution of SVs to participant disease risk, particularly observable with long-read sequencing (Fig. 6d). We further observed that common, low-penetrance SVs were generally associated with more modest relative risks than rare SVs, consistent with evolutionary constraints on high-impact variants. Of note, associations specific to the African genetic ancestry group tended to exhibit larger effect sizes (median AFR-specific = 5.85, median cross-ancestry = 2.72; p = 1.02 × 10^-8^) and lower allele frequencies (median AFR-specific = 0.0004, median cross-ancestry= 0.028; p = 8.68 × 10^-11^), which may represent high-risk variants enriched in individuals of African genetic ancestry (Fig. 6e). In addition, 98 (51.3%) of these lead SV associations contained variants not detected in the paired short-read callset. For example, a 50bp deletion in *PARVB*, which encodes *β*-parvin, a protein that regulates cardiomyocyte shape and sarcomere assembly, was associated with hypertensive heart failure (OR = 1.87; 95% CI: 1.52–2.31; p = 2.32 × 10^-8^)(Fig. 6f, Supplementary Fig. 13, Supplementary Tables 6 and 7)^85^.

Of particular interest are SV-disease associations where SVs intersect and potentially disrupt the coding sequence of genes. Given the potentially severe functional consequences of such disruptions, we applied a relatively permissive threshold (p < 1 × 10^-3^) and identified 114 associations. Among them, 67 were linked to SVs absent from the short-read callset, further demonstrating the potential of LRS in PheWAS. In 62 of the 114 associations (54.3%), SVs showed stronger association signals than nearby SNVs within ±100 kbp, including 40 involving SVs not detected in short-read data (Fig. 6g, Supplementary Fig. 14, Supplementary Table 6). For example, a 200 bp insertion in *CLCN7* was associated with atelectasis (OR = 2.60; 95% CI: 1.62−4.18; p = 3.4 × 10^-4^) (Fig. 6h, Supplementary Fig. 15a-d, Supplementary Tables 6 and 7). This variant introduces a premature stop codon, resulting in a truncated protein lacking key functional domains, as predicted by AlphaFold (Supplementary Fig. 15e). *CLCN7* mutations have been reported to cause severe alveolar atelectasis in mice, providing mechanistic support for this finding in humans^86^. Additionally, we found a 190 bp insertion in *COL14A1* associated with periodontal disease (OR = 4.97; 95% CI: 2.27−10.88; p = 8.9 × 10^-4^) (Supplementary Fig. 16a-e, Supplementary Tables 6 and 7). Prior SNP-based GWAS have also identified an association between *COL14A1* and periodontal disease^87^, although chiefly through intronic variants potentially altering splicing rather than a direct protein coding change. Here the insertion disrupted the coding sequence and caused a truncated protein lacking a segment of the collagenous triple-helical domain and the entire C-terminal tail, likely disrupting protein function (Supplementary Fig. 16f).

## Discussion

The advent of LRS technologies has ushered in a new era in genomics, making large, more complex forms of variation available for the first time. Foremost among the transformative capabilities afforded by LRS are access to repeat-mediated variation that was largely refractory to conventional analysis, and the enhanced capacity for SV discovery. Both types of variation involve intricate complexities and have an outsized influence on human genetic diversity and disease etiology. Herein, we delineate population-scale implementations of LRS for comprehensive analysis encompassing 1,027 participants within AoU who self-identify as Black or African American, a group whose genetic ancestry reflects contributions from populations in Africa shown to harbor high levels of genetic diversity while also experiencing poorer health outcomes and persistent challenges in access to high-quality, tailored healthcare. This Phase 1 effort illustrates the value of integrating population-scale LRS with linked EHR data and provides a foundation for future phases of the program that will extend to the full breadth of diversity represented in All of Us.

The goal of the AoU long-read working group was to capture the broadest swath of genomic variation accessible from participants across the U.S. and to discover novel, informative variation of potential relevance to biomedical research that was cryptic to conventional approaches. From this goal, we substantially expand the landscape of sequence-resolved SVs previously achieved with smaller LRS datasets, benefiting from the progressive refinement of analytical methods over the last five years. To contextualize this undertaking, the largest preceding population study using LRS spanned 3,622 Icelandic individuals and reported the genotyping of 133,886 SVs^88^, whereas the present AoU study jointly genotyped an expansive catalog of 1.2 million SVs from a lenient initial callset, which we further refined to a high-confidence set comprising 666,000 SVs. Key to this was our “mid-pass” strategy (∼8x average coverage), which enables rapid and affordable population-based analyses of SVs within a substantial population-based cohort. These data lay a critical foundation of methods and analytic pipelines for large-scale LRS analyses to facilitate a catalog of genomic diversity that has implications for the diagnosis of rare genetic diseases, trait associations, and the advancement of precision medicine initiatives at scale.

The capacity to scale LRS in AoU necessitated the development of several new approaches to sequencing and analytic strategies that will be relevant for future population-scale datasets in AoU and other resources. By way of comparison, two foundational initiatives in the field, the HGSVC and HPRC, have published studies of 65 and 47 genomes, respectively^11,14^. These flagship programs produced near complete ascertainment of variation in each individual by employing an impressive suite of complementary technologies at extremely high cost per individual, including deep LRS, and sophisticated analytic methods to generate phased diploid genome assemblies. Our approach balanced the value of increasing sequencing depth for variant discovery with the overall goal to advance population-scale LRS analyses, recognizing the necessity to forego complete variant ascertainment to maximize the discovery power of variants potentially relevant to human health across the largest swath of participants.

To enable broader integration of LRS discoveries into existing srWGS-based datasets, we constructed a haplotype-resolved reference panel incorporating AoU and HPRC assemblies. Benchmarking against diploid assemblies revealed high imputation accuracy and precision, particularly for common and intermediate-frequency SVs, and demonstrated that the inclusion of ancestrally diverse haplotypes substantially improves genotyping across all populations. This imputation framework bridges the current cost and throughput gap between srWGS and LRS. It also facilitates the retrospective application of LRS-derived insights to legacy datasets for population genetic discovery and trait association.

Our analyses are distinct from most existing LRS resources in that nearly all participants are linked to extensive EHR data. The integration of LRS-derived variants with EHR data provides a rich resource of genomic variants with potential functional consequences that may be associated with phenotypes not surveyed in prior studies. In particular, we find newly-indentified SVs that intersect with annotated regulatory elements, epigenetically active regions, and even protein-coding genes, with hundreds predicted to be deleterious using CADD-SV^64^. Importantly, SVs uniquely identified by LRS were linked to relevant phenotypes in the EHR, including insertions and deletions predicted to disrupt genes such as *SLC2A12*, *TOX*, and *GALNT5* in participants with corresponding clinical diagnoses^89,90^. Among the thousands of SV-eQTLs detected, fine-mapping analyses further implicated 144 associations in which SVs were the lead causal variants, providing compelling evidence of their critical and underappreciated role in modulating gene expression and disease risk. Notably, our analysis revealed that the absence of an *Alu* element reduced *BID* expression and was associated with increased osteoarthritis risk in the AoU Phase 1 dataset (OR = 2.34, 95% CI = 1.00–5.50, P = 0.049), highlighting the potential regulatory impacts of *Alu* insertions on gene function and their relevance to trait association and disease susceptibility

Beyond aggregate variant discovery, the resolution afforded by LRS permitted targeted dissection of medically consequential loci, including tandem repeat expansions at *FMR1* and *HTT*^48,91,92^, and pharmacogenetically relevant rearrangements. The *FMR1* locus, for example, revealed multiple pre-mutation range alleles, AGG interruption patterns associated with intergenerational stability, and rare haplotypes with pathogenic potential. These findings not only illustrate the diagnostic and pharmacogenomic potential of LRS but also demonstrate the feasibility of applying mid-pass LRS for robust interrogation of structurally complex loci in population cohorts. Pharmacogenetic loci such as *CYP2D6* additionally highlight the importance of LRS for resolving regions of substantial clinical relevance that remain challenging for srWGS. The ability to reconstruct complete haplotypes and identify novel structural and sequence variants in these loci underscores the value of LRS for refining phenotype prediction and enabling equitable pharmacogenomic discovery across diverse populations.

Finally, our PheWAS results revealed new candidate SVs across hundreds of traits for future investigation to resolve their mechanisms of action. Our two-stage approach, first employing a screening threshold to enable balanced comparison of SNVs and SVs, followed by stringent SV-specific filtering and locus-level fine-mapping, was designed to capture a broad spectrum of candidate signals while ensuring that reported associations represent robust and biologically meaningful findings. Using this approach, across the majority (>70%) of the traits considered, we find that SVs have more significant associations than to any of the nearby SNVs. This suggests that when a significant SV association is identified, it is likely to be the strongest association present in the locus. This includes hundreds of newly-identified non-coding SV associations as well as dozens of SVs where the associated variants fall within annotated genes implicated in disease, such as for atelectasis and periodontal disease, where these previously-unknown SVs have direct protein-altering effects.

This initial analysis of LRS demonstrates the profound benefits of incorporating LRS into national biobank efforts. The data presented here provide the most comprehensive view to date of genomic variation in a large self-identified Black or African American cohort, highlighting both population-specific variants and globally relevant loci that shape human health. Our findings establish a critical foundation for more equitable precision medicine, revealing genetic variation that is both biologically consequential and potentially actionable in the future. As LRS becomes increasingly scalable and accessible, its integration into population-scale resources will be essential to build comprehensive variant catalogs. AoU continues the processing and analysis of LRS datasets and anticipates expanding these analyses to over 10,000 participants in the coming year. As these datasets, methods, analyses, and interpretation frameworks expand, they will enable the capture of near-complete diversity of genetic variation in each genome and enhance the potential of genomic medicine.

## Methods

### Population and participants

#### Participant selection

We selected 1,027 participants for PacBio HiFi sequencing based on the following criteria: (1) self-identified as Black or African American, (2) were slated for srWGS data generation from the main project, (3) had sufficient high molecular weight (HMW) DNA available at the biobank (>5 μg), (4) had EHRs available for the majority of participants, (5) were unrelated, and (6) were not known to have a rare disease.

Neither genotyping array nor short-read sequencing data were available at the time of participant selection. Thus, participants for this study were selected because they self-identified as African American or Black, among the other aforementioned considerations. We recognize that self-identified race or ethnicity is not synonymous with genetic ancestry. In the United States, many groups, including African Americans, represent admixed genetic ancestries. Within these groups, individuals may have varying levels of genetic similarity to populations originating from the African continent. As a result, broad continental categories such as ‘African’ can mask considerable within-group diversity.

### Library preparation

#### PacBio HiFi sequencing at HudsonAlpha Discovery

HMW stock DNA concentration was measured using the Picogreen assay (Invitrogen), and the DNA size was estimated using the Fragment Analyzer (Agilent). 5µg of DNA was sheared to a target size of 20-30 kbp on a Megaruptor 3 (Diagenode) and then purified using 0.45x Ampure XP PB beads with a final elution of 40 µL Elution Buffer (EB, PacBio). Sheared DNA was size selected using the Pippin HT instrument (Sage Science) with a target range of 15-22 kbp followed by purification using 0.45x Ampure XP PB beads with a final elution of 50 µl EB. CCS library preparation was performed using the SMRTBell Express Template Prep Kit 2.0 and Enzyme Cleanup Kit 1.0 (PacBio). Each library was barcoded using PacBio Barcoded Overhang Adapters 8A and 8B. Post enzyme cleanup, the libraries were purified 2 times using 1x and 0.6x Ampure XP beads (PacBio) with a final elution in 22 µL EB. Using PacBio SMRTlink, final libraries were annealed with Sequencing Primer v5 and polymerase bound with Sequel II Binding Kit 2.2. Libraries were loaded at 85pM and 30-hour movie time, with Adaptive loading and no pre-extension. Sequel II DNA Internal Control Complex 1.0 was added to each sample as per manufacturer’s recommendation. Sequencing was done on a PacBio Sequel IIe running SMRT Version 10.1.0.119588.

#### ONT sequencing at Baylor College of Medicine

Genomic DNA was quantified using the Lunatic UV/Vis spectrophotometer (Unchained Labs). DNA size was determined using the Agilent Femtopulse. DNA was diluted to 60 ng/µL using nuclease free water to a volume of 50 µL. Covaris g-tubes (Covaris 520079) were used for shearing starting with 3 µg of the diluted DNA. Each sample was passed four times through the orifice of the g-tube using centrifugation at 3,800 rpm. Following bead purification, using AMPure XP beads (Beckman Coulter), the sheared DNA was size-selected on the PippinHT instrument (Sage Science) using the 6-10 kbp High-Pass definition with a minimum size selection threshold of 10 kbp. Loading and elution of samples from the PippinHT cassette were performed following manufacturer instructions. The eluted sample was then bead purified using AMPure XP beads (Beckman Coulter) and resuspended in 50 µL nuclease free water. The average insert size was determined by analyzing the eluate (1 µL) on the Agilent 2100 Bioanalyzer using the DNA 12000 chip for an expected average size of 15-20 kbp. An aliquot (48 µL) of the purified fragments was then used as input for the ONT SQK-LSK114 library preparation kit. End repair/damage repair and adapter ligation were performed as per manufacturer’s instructions. Post ligation, libraries were purified using 1X AMPure XP beads and washed using the Long Fragment Buffer. Final libraries were eluted in 27 µL of the ONT elution buffer and quantified using the Qubit dsDNA quantification broad range assay (Thermo Fisher Scientific). ONT sequencing libraries were processed using R10.4.1 flow cells (FLO-PRO114M, ONT) on a PromethION device (PromethION 24, ONT) with default settings. Briefly, 150-200 ng of sequencing libraries with an average insert size of 15-20 kbp, were loaded onto the flow cell. Sequencing data were collected for 80 hours using MinKnow v22.10.7. On-board basecalling was performed using the super-accurate (SUP) model. If nanopore blockage occurred in the flow cell, samples were nuclease washed and reloaded with 150-200 ng of the library.

### Read mapping and assembly

#### Alignment and metrics calculation

We aligned reads to the <u>GRCh38_no_alt</u> and <u>CHM13v2.0</u> reference sequences. For PacBio HiFi reads, we applied the following pbmm2 v1.12.0 command:

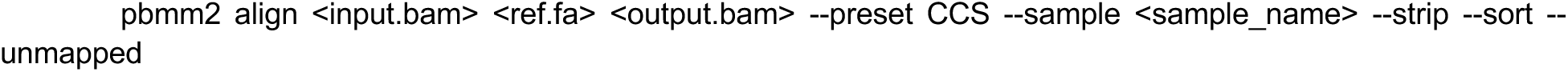

For ONT data, we applied the following minimap2^93,94^ v2.24 command:

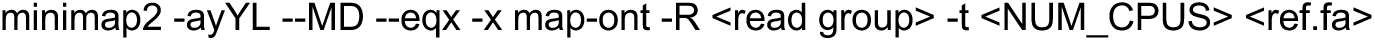

The MD tag encoding mismatched and deleted reference bases were computed post-alignment using the samtools calmd command. We computed genome-wide and per-chromosome coverage metrics using MosDepth, and read length/sequence identity metrics using NanoPlot and custom scripts.

#### De novo assembly

Diploid genome assemblies were generated using hifiasm v0.16.1, which constructs haplotype-resolved contig graphs from PacBio HiFi reads. Assemblies were run per sample using 32 threads and default parameters optimized for CCS reads. The command used was:

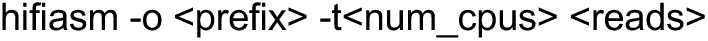

This produced assembly graphs in GFA format: a primary contig graph (<prefix>.bp.p_ctg.gfa) and per-haplotype contig graphs (<prefix>.bp.hap*.p_ctg.gfa). GFA files were converted to FASTA format using the following awk command:

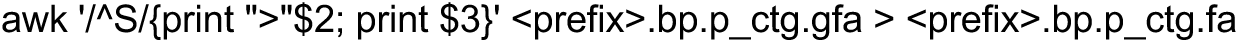

For each haplotype-specific contig set, individual FASTA files were generated:

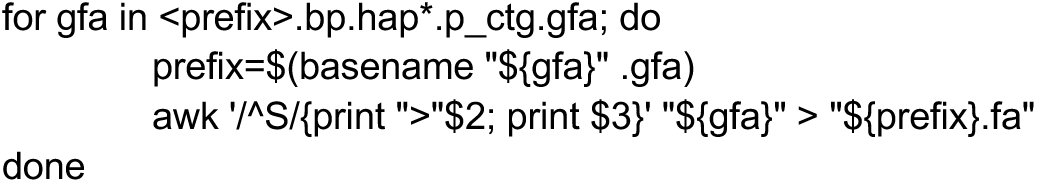

### Variant detection

#### Small variant detection

Small variants, defined as single-nucleotide variants (SNVs) and insertions or deletions (indels) shorter than 50 bp, were detected using DeepVariant v1.4.0. BAM files containing aligned PacBio HiFi reads were used as input, along with the GRCh38 reference genome. DeepVariant was run with the PacBio model type and the following parameters:

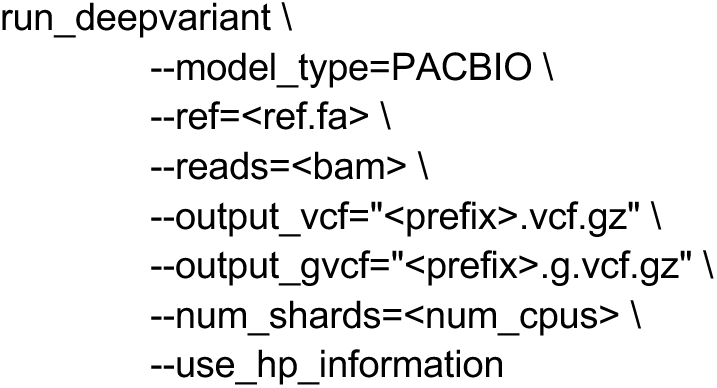

The output VCF and gVCF files were retained for downstream genotyping and joint variant calling. Genomic positions with insufficient coverage or low confidence were annotated accordingly. Quality metrics were computed using standard tools (bcftools, vcftools) to assess depth, transition/transversion ratio, heterozygosity rate, and call concordance in samples with matched short-read data.

#### Joint small variant calling with GLnexus

Joint genotyping of small variants across samples was performed using GLnexus v1.4.1. DeepVariant gVCF files for each sample were first compressed and indexed with bgzip and tabix to prepare them for cohort-level processing. We used the `--config DeepVariantWGS` preset, which is optimized for whole-genome sequencing data generated by DeepVariant, and modified parameters for compatibility with PacBio HiFi reads.

The following command was used for joint genotyping:

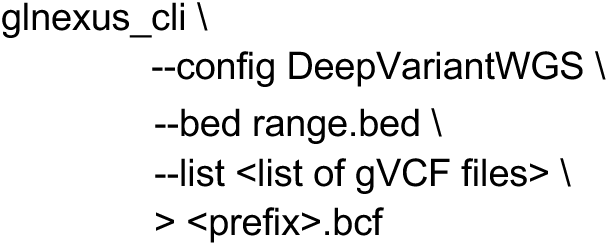

#### Structural variants (SVs)

SVs ≥ 50 bp were identified using an ensemble of two reference-based algorithms, PBSV and Sniffles2, and one assembly-based algorithm, PAV. Each caller was run independently per chromosome to parallelize processing and manage computational resources efficiently. Per-chromosome outputs were later merged to generate per-sample VCFs for downstream integration and harmonization.

PBSV (v2.6.0) was run in discovery and calling mode using the `--tandem-repeats` flag to improve sensitivity for repeat expansions and contractions. Tandem repeat loci were derived from Tandem Repeat Finder annotations available from <u>PacBio</u>. The following commands were applied:

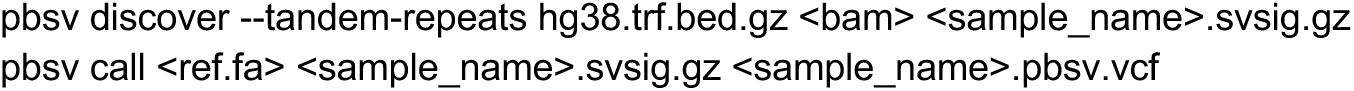

Sniffles2 (v2.0.6) was used to detect SVs with low coverage support using the following command:

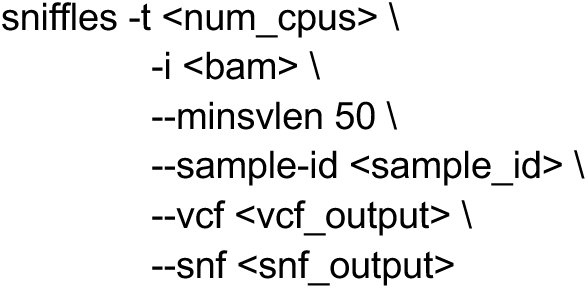

The PAV v1.2.1 pipeline was used for assembly-based SV detection. Primary and haplotype-resolved contigs from hifiasm were aligned to the GRCh38_noalt or CHM13v2.0 reference using minimap2 with -x asm20 parameters. SVs were then called by comparing the assembled contigs to the reference. PAV identifies complex rearrangements, insertions, and deletions not easily captured by alignment-based methods.

The resulting VCFs from PBSV, Sniffles2, and PAV were retained separately and labeled with their source caller for traceability. These per-caller, per-chromosome VCFs were later normalized and harmonized as part of the SV integration and filtering pipeline (see Supplemental Materials: Building a cohort-level SV callset).

### Cohort-level Callset Integration

Per-sample SV callsets from PBSV, Sniffles2, and PAV were normalized to a single callset, clustered and merged within each sample, filtered into stringent and lenient tiers, and then merged across all samples to produce cohort-level callsets. Tool selection and parameterization were guided by benchmarking against assembly-based truth sets. Finally, we re-genotyped the merged callset across the cohort to recover missed variants. Full implementation details, including software versions, parameters, and benchmarking comparisons, are provided in the Supplementary Methods.

### Reference panel construction

We combined SVs from our strict callset on 1,027 AoU participants with 47 HPRC samples into a phased reference panel. Variants were first physically phased within samples using HiPhase^32^, filtered by allele frequency and proximity to SVs, statistically phased across the cohort using SHAPEIT4^95^, and postprocessed to create a pangenome bubble graph Full implementation details are provided in the Supplementary Methods.

### Imputation

Using the reference panel described above, we imputed SVs into (1) all 3,202 samples from the 1000 Genomes Project, and (2) 10,000 self-identified Black or African American AoU srWGS datasets. Genotyping was performed with KAGE to estimate genotype likelihoods from k-mer counts, and phasing and imputation was performed with GLIMPSE^34,35^. Accuracy was evaluated through allele-frequency correlations, Hardy-Weinberg equilibrium, leave-out validation experiments, comparison against assembly-based truth, and Mendelian error rate in 1kGP trios. Full implementation and benchmarking details are described in the Supplementary Methods.

### Repeat loci analysis

#### Catalog-based tandem repeat analysis with TRGT

All samples were aligned to the GRCh38 build of the human genome. TRGT v0.3.3 software^44^ was then run on each sample using default parameters with the Adotto v1.0 catalog^52,96^ padded by 25 bp on each side to resolve the flanking and repeat sequences of the two alleles in each person at each of the 1.7 million specified loci. The alleles called by TRGT were then analyzed for overall length as well as the length of the longest purely repeating segment. Then, variation among the measured alleles for each locus was computed.

#### FMR1 CGG/ HTT CAG repeat analyses

Read consensus sequences were obtained by running Tandem Repeat Genotyping Tool (TRGT) v0.3.3 with the parameters `--genome <ref.fa> --reads <bam> --repeats <trgtcatalog> --output-prefix <sample_name>_TRGT` and `--karyotype XY` for males or `--karyotype XX` for females, using the Adotto v1.0 catalog^96^. Repeat and interruption sequences were resolved and plotted (Code Availability). LongTR (v1.2) was run on the 50 ONT samples for noncanonical interruption validation with the parameters `--bams <bam> --regions <locus.bed> --fasta <ref.fa> --min-reads 0 --tr-vcf <sample_name>.vcf.gz`^97^.

### *CYP2D6* variation

Hifiasm^98,99^ haplotype-resolved assemblies from the 1,027 AoU participants were aligned to the T2T-CHM13v2.0 reference genome^17^. Haplotigs spanning the full *CYP2D6-7* region were extracted, including up to 20 kbp of flanking sequence. Haplotypes with no or multiple haplotigs over the full locus were excluded.

Extracted haplotig sequences were divided into 100 bp k-mers and aligned to *CYP2D6*, *CYP2D7*, and *CYP2D7*-spacer T2T-CHM13v2.0 reference sequences. K-mers were colored according to best match by MAPQ score, and k-merized sequences were plotted (Code Availability). This pipeline was tested on 47 high-quality phased HPRC assemblies, for which the copy number of *CYP2D6* and *CYP2D7* showed 100% concordance to reported HPRC haplotypes, disregarding hybrid alleles^14^.

Short-read haplotyping from CRAM files for the 1,027 long read samples was completed using validated haplotype callers Aldy (v4.4)^100^, Cyrius (v1.1.1)^101^, PyPGx (v0.20.0)^102^, and StellarPGx (v1.2.7)^103^. A consensus call was calculated as a composite value of the haplotype call from all callers based on majority agreement between callers (minimum 2). An independent short-read analysis in which PyPGx was run equivalently except using control gene VDR produced the same PyPGx calls.

Star alleles were assigned by aligning PharmVar reference sequences to each non-CNV haplotype and selecting a best-matching star allele by CIGAR scoring, requiring the presence of all defining variants^104^. Diplotypes and phenotypes were derived according to CPIC guidelines for samples for which both haplotypes were fully resolved^105^.

To assess the accuracy of our star allele calling pipeline, star allele calls on 40 HPRC assemblies in the high-coverage 1KGP set were validated against the short-read consensus calls, achieving 95% concordance for unambiguously assigned non-SV haplotypes^14,39^. For all disagreements our call was supported by at least one caller.

Missense variants were identified by predicting haplotype coding sequences using Augustus (v3.5.0)^106^ with gene structure hints, aligning these to the GRCh38 coding sequence, and comparing the resulting amino acid sequences to those for reference star alleles. Lollipop plots were generated with ProteinPaint^107,108^. AlphaMissense hg38 pathogenicity classifications^61^ were obtained for each base change from the *CYP2D6* (P10635) AlphaFold heatmap.

#### Genotype assessment with high-coverage ONT

Both stringent and lenient callsets were genotyped on ONT samples aligned to GRCh38 and CHM13-T2T references independently as indicated above. Kanpig^28^ v0.3.1 was employed for variant genotyping with the following parameters: `--hapsim 0.97 --chunksize 500 --maxpaths 1000 –gpenalty 0.04`. The genotyped VCF files were sorted using bcftools v1.21 sort with the -Oz parameter. Tabix was subsequently utilized to generate indexes for the sorted genotyped VCF files with the -p vcf parameter. Truvari^26^ v4.3.1 with default parameters, was used to assess the concordance between the genotyped VCF files and the original VCF files.

### Impact prediction of SVs

From the *All of Us* SV callset, we filtered out SVs with genotypes labeled as "./." or "0/0" across all samples and those ≤50 bp in length. We assessed the functional impact of the remaining SVs using CADD-SV^64^ and analyzed the relationship between the number of samples sharing each SV and its predicted pathogenicity score. For SVs predicted to be likely pathogenic or absent from the matched short-read SV dataset, we examined their overlap with medically relevant genes, including those listed in OMIM^62^, high-priority genes associated with inherited diseases^63^, oncology/tumor suppressor genes from OncoKB^68^, and genes in the American College of Medical Genetics and Genomics (ACMG) list^41^.

To investigate the potential impact of these SVs on human health, we then accessed EHRs for each individual from the *All of Us* Researcher Workbench and mapped the clinical conditions to Systematized Nomenclature of Medicine - Clinical Terms (SNOMED CT). For high-scoring SVs intersecting coding regions of medically significant genes, we used AlphaFold3 to predict the resulting protein structures and compared them to the reference structures to assess potential functional disruption at the protein level.

### Cross-population SV interpretation

We analyzed SV counts per sample and their allele frequencies across 2,504 unrelated 1KGP samples from five continental groups. To assess population stratification, we performed a PCA on genotyped SVs using PLINK^109^, excluding variants within tandem repeat regions^110^. For comparison, we conducted a PCA using chromosome 1 SNVs from the 1KGP, applying the same filtering criteria. To investigate the functional impact of SVs, we mapped them to gene regions using GENCODE annotations, regulatory elements from Ensembl and analyzed their distribution across populations. F_st_ values were calculated for each SV between African and non-African populations with BCFtools^111^.

### SV-eQTL analysis

We performed eQTL calling following the method described by Kirsche et al^76^ to investigate the impact of SVs on gene expression in 731 samples from the 1KGP with the matched RNA-seq data. Briefly, SV genotypes were categorized as 0 (homozygous reference), 1 (heterozygous alternative), and 2 (homozygous alternative), and only SVs with at least 5% of samples in each genotype class were selected to ensure statistical robustness. Using an ordinary least squares (OLS) linear regression method, we evaluated associations between SV genotypes and normalized gene expression levels, including sex as a covariate. Then we computed gene-level eQTL p-values by applying Bonferroni correction to the minimum eQTL p-value for each gene, adjusting by the number of eQTLs associated with that gene. These p-values were further corrected for multiple testing using the Benjamini-Hochberg method at a false discovery rate of ≤5%. All reported SVs had a mean genotype posterior (GP) ≥ 0.7 across the cohort. To validate genotyping accuracy, we selected 40 samples with both long-read and short-read data from the 1KGP cohort and computed per-genotype concordance for each sample (Supplementary Fig. 9f).

To compare the effects of SVs and SNVs on genes associated with both variant types, we used the previously generated SNV data from (https://ftp.1000genomes.ebi.ac.uk/vol1/ftp/data_collections/1000G_2504_high_coverage/working/20201028_3202_phased/), restricted to the same 731 samples. For each gene, we evaluated SNV-expression associations within a 1 Mbp window of the gene using the same OLS regression model applied to SVs. We then selected the top SV-eQTL and the 1,000 most strongest SNV-eQTLs per gene to calculate the z-scores and LD between variants. These values were used as input for the CAVIAR^79^ fine-mapping analysis to estimate the posterior probability of each variant being causal.

### Overlap between SVs and GWAS-significant traits

To investigate potential associations between SVs and phenotypes, we downloaded the GWAS Catalog v1.0 database and identified genome-wide significant variants (p < 5 × 10^-8^) located within ±100,000 bp of our SVs. Genotype data for these GWAS variants was obtained from the 1KGP Phase 3 dataset^37^, focusing on the same set of 2,504 unrelated samples used for SV genotyping. For each SV-GWAS variant pair, we calculated LD and retained those with r^2^ ≥ 0.5. To characterize disease- and disorder-related traits, we used the ontologies from the Ontology Lookup Service (http://www.ebi.ac.uk/ols) and condition-domain concepts from the SNOMED vocabulary to classify traits, enabling further exploration of the potential medical implications of associated variants.

To further evaluate the medical relevance of SVs linked to GWAS variants, we identified SV carriers among 1,027 AoU participants and retained SVs present in at least 10 individuals. We then extracted EHR data from AoU Researcher Workbench and included 848 participants with available condition records. Clinical conditions with at least 10 cases and 10 controls were selected for analysis. We applied Fisher’s exact test to assess associations between SV presence and clinical diagnoses. Analyses of sex-specific conditions were limited to individuals of the corresponding sex. All reported SVs had a mean GP ≥ 0.7 and a minor allele frequency (MAF) ≥ 0.05 among the 2,504 1KGP samples. Genotyping accuracy was validated using the same 40 long-read/short-read 1KGP samples (Supplementary Fig. 10e).

### Phenome-wide analysis of SVs in 10,000 self-identified Black or African American participants

To explore the clinical impact of identified SVs, we integrated genomic data with EHRs from an expanded AoU cohort. We firstly genotyped and imputed variants in 10,000 short-read samples from self-identified Black or African American participants, retaining SVs present in at least 10 individuals. Clinical condition data were obtained from AoU Researcher Workbench, and only conditions with at least 100 cases and 100 controls were included to ensure statistical power. To improve the accuracy of the phenome-wide association study, we excluded conditions not known to be influenced by genetic factors and those with overly broad or ambiguous names.

We applied Fisher’s exact test to identify conditions significantly associated with SV carriers. Analyses of sex-specific conditions were conducted only in individuals of the relevant sex. For SVs associated with disease, we extracted the GP values across all SV carriers and calculated the mean GP. SVs with a mean GP < 0.7 or deviating from Hardy-Weinberg equilibrium (HWE < 1×10^-5^) were excluded to reduce false-positive associations. We next compared SV AFs between the 1,027 AoU long-read samples and the 10,000 short-read samples, excluding SVs with AF differences ≥ 1.5-fold.

Genotype confidence was further assessed by re-genotyping SVs with Locityper^112^ and comparing concordance with imputed allele calls across carriers. This was performed in two steps: (1) building a variant database, for example with:

~~~
locityper add -@ ${nthreads} -d vcf_db -v input.vcf.gz -r reference.fa -j counts.jf -L regions.bed
~~~

and (2) re-genotyping each variant against individual samples with:

~~~
locityper genotype -a sample.cram -r reference.fa -d vcf_db -p locityper_preproc -S greedy:i=5k,a=1 -S anneal:i=20,a=20 --subset-loci "${locus_name}" -o out_dir
~~~

SVs were removed if fewer than 70% of carriers showed allele differences < 20 between the two methods (Supplementary Fig. 11).

To evaluate the robustness of identified associations, we excluded samples with kinship > 0.177 or with fewer than 10 SV carriers and applied Firth’s penalized logistic regression, while adjusting for age, sex, sequencing center, and the first ten principal components. Associations were retained only if the effect direction was consistent across both methods and the OR estimates differed by no more than 1.5-fold (Supplementary Fig. 12).

We used a moderate significance threshold (p < 1×10⁻⁵) for the initial association scan to facilitate direct comparison of association strength between SNVs and SVs across traits, rather than to declare genome-wide significance. This screening threshold was chosen to capture a broader set of candidate variants for downstream evaluation. To determine whether the SV was the most significantly associated variant for the condition from this screening set, we identified SNVs within ±100 kbp and assessed their association with the corresponding phenotype using the same statistical method. Then, we applied CAVIAR for fine-mapping, incorporating each significant SV and the 1,000 nearby SNVs with the most significant association signals to estimate their posterior probabilities of causality. Finally, for SVs intersecting coding regions of medically significant genes with known disease relevance, we used AlphaFold3 to predict altered protein structures and compared them to reference models to evaluate potential functional consequences at the protein level.

We then assessed whether the disease-associated SVs were unique to the long-read dataset by comparing them with the short-read SV callset derived from 990 of the 1,027 samples used to construct the long-read reference panel (for comparison methods, see Supplementary Methods: Variant annotation and comparison to external datasets). We quantified the number of associations in which the SV was absent from the matched short-read callset. In addition, we identified associations specific to individuals with African genetic ancestry by selecting SVs that were only imputed among the 2,504 unrelated 1KGP samples with African genetic ancestry.

Furthermore, we performed a PCA using PLINK on 2,504 unrelated 1KGP individuals and the 10,000 AoU samples to assess population structure. For the reported associations in the results, AoU samples were further stratified into: (i) cases vs. controls, (ii) SV carriers among controls vs. all other individuals, and (iii) SV carriers among cases vs. all other individuals. The diffuse distribution of these subgroups across the PCA plots suggests the observed associations reflect real biological signals rather than batch effects or other artifacts in phenotypes coded at a single hospital or collection site.

## Data availability

The *All of Us* Research Hub has a tiered data access data passport model with three data access tiers. The Public Tier dataset contains only aggregate data with identifiers removed. These data are available to the public through Data Snapshots (https://www.researchallofus.org/data-tools/data-snapshots/) and the public Data Browser (https://databrowser.researchallofus.org/). The Registered Tier curated dataset contains individual-level data, available only to approved researchers on the Researcher Workbench. At present, the Registered Tier includes data from EHRs, wearables and surveys, as well as physical measurements taken at the time of participant enrolment. The Controlled Tier dataset contains all data in the Registered Tier and additionally genomic data in the form of short- and long-read whole-genome sequencing and genotyping arrays, previously suppressed demographic data fields from EHRs and surveys, and unshifted dates of events. At present, Registered Tier and Controlled Tier data are available to researchers at academic institutions, nonprofit institutions, and both nonprofit and for-profit health care institutions. Work is underway to begin extending access to additional audiences, including industry-affiliated researchers. Researchers have the option to register for Registered Tier and/or Controlled Tier access by completing the *All of Us* Researcher Workbench access process, which includes identity verification and *All of Us*-specific training in research involving human participants (https://www.researchallofus.org/register/). Researchers may create a new workspace at any time to conduct any research study, provided that they comply with all Data Use Policies and self-declare their research purpose. This information is made accessible publicly on the *All of Us* Research Projects Directory at: https://allofus.nih.gov/protecting-data-and-privacy/research-projects-all-us-data.

Additional ONT data is available in a featured workspace available for those who have registered Controlled Tier access at:

https://workbench.researchallofus.org/workspaces/aou-rw-a138edac/longreadsontvalidationdata/data.

Reported results with group counts under 20 are disclosed under a granted exception to the Data and Statistics Dissemination Policy from the All of Us Resource Access Board.

## Code availability

All analysis workflows, code, and processed data supporting this study are made available to the research community through the *All of Us* Researcher Workbench and a public GitHub repository. The primary codebase, including links to workflows used for long-read alignment, variant calling, and quality control, is hosted at <u>GitHub - all-of-us/long-reads-public-codebase</u> and runnable in the cloud-native bioinformatics platform, Terra (https://app.terra.bio/). In addition, we created a dedicated workspace on the *<u>All of Us</u>* <u>Researcher Workbench</u> (“RW”), which includes interactive Jupyter notebooks, access to the phased long-read variant callsets, and links to reference files and metadata. Registered researchers with approved access to the *All of Us* controlled tier can explore, replicate, and extend the analyses directly within the RW platform, ensuring that insights from LRS can be broadly leveraged by the genomics community.

## Acknowledgments

We gratefully acknowledge *All of Us* participants for their contributions, without whom this research would not have been possible. We also thank the National Institutes of Health’s *All of Us* Research Program for making available the participant data examined in this study.

We thank Timofey Prodanov and Tobias Marschall for insightful discussions on the use of Locityper and for their helpful code improvements for the benefit of our study. We thank Peter Audano for his assistance in developing a cloud-capable workflow for PAV.

The 1000 Genomes Project high-coverage whole genome data used in this study were generated at the New York Genome Center with funds provided by NHGRI Grant 3UM1HG008901-03S1.

## Author information

### Authors and Affiliations

**Center for Genomic Medicine, Massachusetts General Hospital, Boston, MA, USA; Program in Medical and Population Genetics, Broad Institute of MIT and Harvard, Cambridge, MA, USA; Department of Neurology, Massachusetts General Hospital and Harvard Medical School, Boston, MA, USA**

Harrison Brand, Xuefang Zhao

**Data Sciences Platform, Broad Institute of MIT and Harvard, Cambridge, MA, USA**

Evie Wan, Lucas van Dijk

**Department of Genetics, Cell Biology, and Development, University of Minnesota, Minneapolis, MN, USA**

PingHsun Hsieh, Natthapon Soisangwan

**Department of Genome Sciences, University of Washington School of Medicine, Seattle, WA, USA**

Tristan Shaffer, Joshua D Smith, Aparna Radhakrishnan, Sean McGee

**Department of Laboratory Medicine and Pathology, Mayo Clinic, Rochester, MN, USA**

Stephen N Thibodeau, Mine S Cicek

**Department of Pharmacy and Therapeutics, University of Pittsburgh School of Pharmacy, Pittsburgh, Pennsylvania, USA**

Andrew Haddad, Philip E Empey

**Discovery Life Sciences, Huntsville, AL, USA**

Nripesh Prasad

**HudsonAlpha Institute for Biotechnology**

Jane Grimwood

**Human Genome Sequencing Center, Baylor College of Medicine, Houston, TX, USA**

Ginger A Metcalf, Huyen Dinh, Yi Han, Jianhong Hu, James Hwang, Ziad Khan, Medhat Madmoud, Hua Shen, Vanesa Vee, George Weissenberger, Yiming Zhu,

**Vanderbilt Institute for Clinical and Translational Research, Vanderbilt University Medical Center, Nashville, TN, USA**

Chris Lord

### Contributions

The *All of Us* Biobank (Mayo Clinic - S.N.T., M.S.C.) collected, stored, and plated participant biospecimens. HudsonAlpha and Discovery Life Sciences (N.P., C.B., S.L., J.G.) generated the PacBio whole-genome sequencing data, and Baylor College of Medicine (F.J.S., G.A.M., D.M.M.) generated the ONT whole-genome sequencing data. The Broad Institute (S.K.L., F.C., R.L.R., H.S., Y.H., E.L., S.S., B.S., S.Z., E.W., L.v.D., K.V.G.) performed sample quality control, alignment, variant calling, genome assembly, led the SV filtering and integration subgroup, conducted SV quality control analyses, and developed and executed the scalable imputation framework. Baylor College of Medicine (A.E., X.C., F.J.S.) and Johns Hopkins University (Q.L., M.S.) contributed to the SV filtering and integration subgroup. Johns Hopkins University (Q.L., M.S.) also led the SV interpretation and SV-phenotype association analyses. The University of Washington (J.W., W.T.H., K.P., A.N.R., I.W., T.S., J.D.S., E.E.E.) and University of Miami (M.D., I.X., S.Z.) contributed to SV filtering and integration, performed additional SNV, indel, and SV quality control, and led the analyses of repeat expansions and CYP2D6. The *All of Us* Research Program’s Long-Read Working Group contributed additional analyses. The Working Group co-chairs and NIH *All of Us* Research Program Staff provided crucial programmatic support. Members of the manuscript writing group (K.V.G., Q.L., J.W., S.K.L., F.C., M.D., Y.M., M.C.S., M.E.T., E.E.E.) wrote the manuscript, which was revised with contributions and feedback from all authors.

## Ethics declarations / Competing Interests

K.V.G. is a co-inventor on a pending international patent application related to long-read RNA isoform sequencing, licensed to Pacific Biosciences, but not used in this study. E.E.E. is a scientific advisory board (SAB) member of Variant Bio, Inc. F.J.S. receives research support from Illumina and Nanopore. The remaining authors declare no competing interests.

## Supplementary information

### Supplementary Information

This file contains Supplementary Methods, legends for Supplementary Figs. 1-24, and Supplementary Note.

**Supplementary Table 1: Summary of AoU Phase1 SVs imputed in 1KGP samples**

This table reports SV genomic coordinates, identifier, length, type, and mean genotyping posterior across carriers, allele frequencies across the five continental groups and F_st_ values between African and non-African populations. Gene-level annotations include overlaps with OMIM, disease-associated, OncoKB cancer, and ACMG genes; for each category, overlaps are reported separately for gene bodies and coding sequences, along with pLI and LOEUF scores of intersecting genes. The table also reports overlaps with regulatory elements, segmental duplications, and tandem repeats, and indicates whether each SV is present in other long-read callsets (HPRC, HGSVC, 1000G-ONT) or in the AoU GATK-SV callset (detected in 990 of 1,027 Phase I matched samples). In addition, the table summarizes SV–eQTLs and SVs in LD with variants from the GWAS Catalog. For SVs in LD with GWAS variants, associations with disease are computed using AoU Phase I data, including frequencies of carrier and non-carrier with the disease, OR, 95% CI, and P values. All eQTL- and LD-related SVs are genotyped with Locityper in 2,504 unrelated 1KGP samples, and the table reports the proportion of carriers with allele differences <20 between imputed and Locityper calls. Selected SVs are further reviewed using matched AoU short-read data and long-read assemblies.

**Supplementary Table 2: *CYP2D6* star allele disagreements in long vs. short reads**

This table reports 90 unambiguous disagreements were observed in 1,008 resolved haplotypes between our long read assembly-based star allele calling pipeline and consensus calls from genotypers Aldy, Cyrius, PyPGx, and StellarPGx run on short reads for the same samples. Most non-CNV discrepancies are due to potential false duplication of one haplotype in the low-coverage assemblies. Commas indicate multiple disagreeing star alleles.

**Supplementary Table 3: Characterization of AoU Phase 1 SVs**

This table lists SV genomic coordinates, identifier, length, type, and confidence assessment. Gene-level annotations include overlaps with OMIM, disease-associated, OncoKB cancer, and ACMG genes; for each category, overlaps are reported separately for gene bodies and coding sequences, together with pLI and LOEUF scores of intersecting genes. The table also indicates whether each SV is present in other long-read callsets (HPRC, HGSVC, 1000G-ONT) or in the AoU GATK-SV callset (detected in 990 of 1,027 Phase I matched samples). Additional annotations include overlaps with regulatory elements, segmental duplications, tandem repeats, and CADD-SV pathogenicity scores. Selected high-scoring SVs are further evaluated using matched AoU short-read data and long-read assemblies, and genotyped with Locityper in 1,027 Phase I samples; the table reports the proportion of carriers with allele differences <20 between reported and Locityper calls.

**Supplementary Table 4: Curated AoU Phase 1 SVs**

This table reports curated SVs with high CADD-SV scores and relevance to medically significant genes, using the same annotation categories as Supplementary Table 3.

**Supplementary Table 5: Curated AoU Phase 1 SVs imputed in 1KGP samples**

This table reports a curated set of SVs prioritized for functional relevance, including SV–eQTLs and SVs in LD with variants from the GWAS Catalog, annotated using the same categories as Supplementary Table 1.

**Supplementary Table 6: Annotation and validation of AoU Phase 1 SVs imputed in 10,000 short-read samples of African ancestry**

This table reports SV genomic coordinates, identifier, length, type, number of carriers, and mean genotyping posterior across carriers. Gene-level annotations include overlaps with OMIM, disease-associated, OncoKB cancer, and ACMG genes; for each category, overlaps are reported separately for gene bodies and coding sequences, together with pLI and LOEUF scores of intersecting genes. Additional annotations include overlaps with regulatory elements, segmental duplications, and tandem repeats. The table also indicates whether each SV is present in other long-read callsets (HPRC, HGSVC, 1000G-ONT) or in the AoU GATK-SV callset (detected in 990 of 1,027 Phase I matched samples). It further summarizes genome-wide and CDS-specific PheWAS results, reporting the percentage of carriers and non-carriers with each condition, OR, 95% CI, and P values for SV–disease associations, along with validation using Firth’s penalized logistic regression and the corresponding best-hit SNP ORs and P values. All PheWAS-identified SVs are genotyped with Locityper in 10,000 AoU short-read samples, and the table reports the proportion of carriers with allele differences <20 between imputed and Locityper calls. Selected SVs are further evaluated using matched AoU short-read data and long-read assemblies.

**Supplementary Table 7: Curated structural variants prioritized by PheWAS**

This table reports SVs curated from PheWAS analyses using the same annotation categories as Supplementary Table 6.

