## Supplemental information for "Population-scale Long-read Sequencing in the *All of Us* Research Program"

**Table of Contents**

**SUPPLEMENTARY METHODS	3**

Building a cohort-level SV callset	3

Intra-sample merging	3

Intra-sample filtering	5

Inter-sample merging	6

Genotyping and imputation of srWGS case samples with LRS panel resources: motivation	9

Creation of a LRS reference panel	9

Genotyping, phasing, and imputation of srWGS case samples	10

Evaluations of the LRS reference panel and imputed srWGS cases	11

Leave-out evaluation of PanGenie and KAGE+GLIMPSE	11

Leave-out evaluation of imputation quality metrics	12

Evaluation against assembly-based truth	12

Evaluation of Mendelian consistency using 602 1kGP imputed trios	14

Creation of the LRS reference panel: methods	14

Physical phasing with HiPhase	14

Filtering, concatentation, and deduplication	14

Removing colliding variants	14

Statistical phasing with SHAPEIT4	15

Converting to bubble representation	15

Genotyping, phasing, and imputation of srWGS cases: methods	16

Genotyping with KAGE	16

Phasing and imputation with GLIMPSE	17

Variant annotation and comparison to external datasets	17

Detection and annotation of mobile element insertions	18

CYP2D6 variation	18

Assembly sequence extraction	18

CNV/ SV k-mer plotting	19

Star allele and phenotype assignment	19

Star allele validation	19

Missense variant analysis	20

**SUPPLEMENTARY NOTES	21**

Supplementary Note 1: Noncanonical and rare FMR1 and HTT interruptions	21

**SUPPLEMENTARY FIGURES	22**

Supplementary Fig. 1: Overview of the AoU long-read variant discovery, reference panel construction, and imputation pipelines.	22

Supplementary Fig. 2: Median Mendelian discordance rates of SVs for three trios on reference GRCh38	23

Supplementary Fig. 3: Median Mendelian discordance rates of SVs for three trios on reference T2T-CHM13	24

Supplementary Fig. 4: Median Mendelian discordance rates of SVs for three trios, stratified by child coverage level, SV filtering stringency, and genome context.	25

Supplementary Fig. 5: Mobile element insertions and allelic heterogeneity in SVAs in the lenient SV callset.	26

Supplementary Fig. 6: Population-level analysis of genotyped structural variant callset	27

Supplementary Fig. 7: Repeat expansion at ATXN3	28

Supplementary Fig. 8: Functional annotation and gene associations of AoU SVs	29

Supplementary Fig. 9: Population patterns and functional examples of SV-eQTLs	31

Supplementary Fig. 10: SVs in linkage disequilibrium with GWAS variants associated with diseases and traits	32

Supplementary Fig. 11: Population structure and evaluation of SV genotypes	34

Supplementary Fig. 12: Evaluation of SV–disease associations	36

Supplementary Fig. 13: Variant evidence and population structure of disease-associated SVs	37

Supplementary Fig. 14: Clinical associations of structural variants in coding regions	38

Supplementary Fig. 15: Characterization of an atelectasis-associated SV	39

Supplementary Fig. 16: Characterization of an SV associated with periodontal disease	40

Supplementary Fig. 17: Heterozygous-variant counts for AoU+HPRC panel and 3,202 1kGP imputed samples	41

Supplementary Fig. 18: Allele-frequency correlation and Hardy-Weinberg equilibrium for AoU+HPRC panel and 3,202 1kGP imputed samples	42

Supplementary Fig. 19: Allele-frequency correlation and Hardy-Weinberg equilibrium for AoU+HPRC panel and 10,000 self-identified Black or African American AoU imputed samples	43

Supplementary Fig. 20: Comparison of KAGE+GLIMPSE and PanGenie accuracy in leave-out evaluations with HGSVC2 (assembly-based) and AoU+HPRC (8x LRS) panels	44

Supplementary Fig. 21: Imputation dosage r2 in a leave-many-out evaluation over 40 HPRC samples	45

Supplementary Fig. 22: Non-reference concordance rate in a leave-many-out evaluation over 40 HPRC samples	46

Supplementary Fig. 23: Accuracy of imputation panel and cases against assembly-based truth, outside of TR/homopolymer regions	47

Supplementary Fig. 24: Accuracy of imputation panel and cases against assembly-based truth, inside of TR/homopolymer regions	48

Supplementary Fig. 25: Mendelian consistency for 602 1kGP imputed trios	49

**REFERENCES	51**

### SUPPLEMENTARY METHODS

#### Building a cohort-level SV callset

Recall that, for each sample, we generated SV calls using PAV, pbsv and Sniffles. The next step consists in building a unified, cohort-level callset whose rows are distinct, de-duplicated SVs, and whose columns are the genotype of every sample that contains each variant. Although Sniffles and pbsv provide functions for building such a cohort VCF directly from their single-sample signature files, each tool works only on its own calls, and one would still need to integrate the two cohort-level VCFs with each other, as well as with all the single-sample PAV calls. Thus, we decided to build the cohort-level callset by first clustering all the calls in each sample, and then integrating all intra-sample-merged VCFs across the cohort.

Before proceeding, we enforced a common format over all single-sample, single-caller VCFs, as follows. We filtered out every PAV call shorter than 50bp. Then, we converted multiallelic records into biallelic records using BCFtools norm, and we filtered out every BND call, every symbolic CNV and INS call, and every call longer than 1Mbp. We converted every remaining symbolic call to an explicit pair of REF and ALT alleles, we set every missing genotype to zero, and we made sure that every record has the correct SVLEN value. We assigned a constant integer quality to every call from the same caller (PAV=4, pbsv=3, Sniffles=2), based on which variant representations we observed to be more accurate against the Genome in a Bottle (GIAB) draft SV benchmark for HG002 (based on the v0.9 release of the T2T HG002 Q100 assembly^1,2^, and we assigned quality 1 to every symbolic call that we converted to explicit alleles. Note that we did not apply any hard filter on the QUAL, FILTER or GT fields of a VCF: in particular, we kept calls that were not marked as PASS by their caller, as well as calls with 0/0 or missing GT.

##### Intra-sample merging

To decide how to cluster the PAV, pbsv and Sniffles calls of each sample, we assessed the rate of correct merging in several tools from a sampling of the state of the art at the time of the study: Truvari collapse v4.2.2^3^, Jasmine v1.1.5^4^, sv-merger^5^ (commit b774523), svimmer^6^ (commit f2d78b2), and SV-Pop v3.4.2^7^ (BCFtools merge^8^ collapses only exact matches and was not evaluated). We set the following Truvari parameters:

--sizemin 0 --sizemax 1000000 --keep maxqual --gt het --intra --pctseq 0.90 --pctsize 0.90 --refdist 500

which make the method use sequence similarity when collapsing calls, and select a call of highest quality as a representative of each cluster. We observed that some callers made redundant homozygous calls, and these parameters allowed Truvari to merge them. We kept the default parameters in every other tool: this means that Jasmine did not take the sequence similarity of insertions into account (sv-merger and svimmer do not use sequence similarity by design). We note that sv-merger is the only tool that uses a different merging strategy inside and outside a tandem repeat track provided in input. To evaluate the merging tools, we ran SV discovery on the 47 PacBio BAMs from the Human Pangenome Reference Consortium (HPRC)^9^, downsampled to 8x and mapped to both GRCh38 and CHM13. We matched the calls to the phased calls made by dipcall^10^ (with default parameters) on the corresponding high-quality diploid assemblies aligned to both references. Matching was restricted to variants with length ≥50 bp in dipcall’s confident regions. The output of dipcall was determined to represent a reliable truth set by comparison to the GIAB draft SV benchmark for HG002 (on GRCh38) in its confident regions, using Truvari bench with default parameters (F1=0.977), Truvari bench with Truvari refine (F1=0.983), and vcfdist^11^ (F1=0.952). Additionally, the fraction of GRCh38 spanned by high-confidence dipcall regions (2.75 Gbp, 89.3% of the genome) was comparable to the fraction spanned by the high-confidence regions from GIAB (2.78 Gbp, 90.3%).

For each HPRC sample, intra-sample merging of the three discovery tools’ results were compared to the dipcall assembly-derived baseline VCF using Truvari bench with default parameters and multi-matching (i.e., allowing multiple calls to be matched to the same entry in the truth set). The output of Truvari collapse achieved 0.649 average recall (the highest) and 0.689 average precision across the two references. Jasmine and sv-merger yielded comparable results, with slightly higher average precision (0.706 and 0.724, respectively) and slightly lower average recall (0.631 and 0.628, respectively). Svimmer showed low recall (0.426) and high precision (0.740), suggesting that it over-merges, and SV-Pop failed to complete. Both recall and precision tended to be higher in CHM13 than in GRCh38, with svimmer being the only exception. The low recall is likely due to the fact that state-of-the-art SV callers are not optimized for the 8x coverage regime. Compared to the output of BCFtools merge (which just collapses exact matches), Truvari discarded an average of 42 true-positive (TP) calls, whereas the next best tool (Jasmine) discarded an average of 600 true-positive calls. A merged callset should also have low redundancy, and this can be measured by the ratio between the number of calls in the truth set that are matched in the merged set, and the cardinality of the merged set. This ratio was closest to one for Truvari (average 1.008), followed by Jasmine (average 0.985) and sv-merger (0.977); svimmer consistently over-merged (average 1.087).

To evaluate over-merging and under-merging in more detail, we took all the raw calls, all the true calls (phased), and all the calls from a given tool and analyzed windows with a set of overlapping or adjacent calls that are at distance ≥1kb from the next closest call. If a window contains exactly two heterozygous calls in the truth set (from different haplotypes), and if there are corresponding true-positive heterozygous calls among the raw calls in the window (one for each haplotype), then we say that a tool over-merges if it outputs less than two heterozygous calls in the window. Of the 19,740 loci meeting the selection criteria across all samples, Truvari had the lowest fraction of windows with over-merging (average 40.3%), followed by sv-merger (56.7%) and Jasmine (64.5%), whereas svimmer over-merged in 91.9% of its windows on average. Symmetrically, if a window contains exactly one homozygous call in the truth set, and if there are more than one true-positive calls among the raw calls in the window, we say that a tool under-merges if it reports more than one call in the window. Truvari had the largest fraction of windows with no under-merging (91.9% on average), followed by sv-merger (85.4%) and Jasmine (84.9%).

Finally, given a cluster of merged calls, its representative should be the record that is most similar to the truth. Consider the set *T* of all true calls that have a match in the output of every merging tool. For every call in *T*, we can find a most similar call in the set of all raw calls. For every merging tool, we counted how many calls in *T* see their most similar call being reported in the output of the tool: Truvari achieved 90.5% for DELs and 87.2% for INS, followed by Jasmine with 88.5% and 85.8%, respectively. We computed similarity as the average of the sequence similarity score, length similarity score, and reciprocal overlap score emitted by Truvari bench.

As a result of this analysis, we decided to cluster the PAV, pbsv and Sniffles calls of each sample using Truvari. Note that the records in the resulting VCF have heterogeneous annotations, since they may come from different callers, and that Truvari annotates which callers contributed to each output record.

##### Intra-sample filtering

An SV merging tool can only reduce the number of false-positive (FP) calls by collapsing similar false calls together, but it cannot remove isolated false calls or clusters of false calls. A small fraction of FPs in each sample, possibly with different sequence, location or length in different samples, may accumulate over a large cohort to the point that FPs may become a consistent fraction of the final inter-sample-merged VCF. Previous studies have filtered out FPs by keeping e.g., only records that are supported by at least two callers or by a specific trusted caller^7,12,13^. This heuristic assumes that different callers use significantly different discovery algorithms and input features, which might not be the case in our setting since we used two alignment-based callers that work on the same BAM input, and it does not take advantage of the unique capabilities of each caller, if any. For example, by comparing our HG002 callset to the Q100 assembly, we observed that PAV (our only assembly-based caller) emits several FPs due e.g., to Hifiasm^14^ misassemblies at 8x coverage, but it is also capable of detecting true SVs that are missed by pbsv and Sniffles. Keeping only calls that are supported by at least two callers yields also a fixed tradeoff between false-positive rate (FPR, the fraction of FPs that are kept after filtering) and true-positive rate (TPR, the fraction of TPs that are kept after filtering), which might not be the best achievable tradeoff nor the optimal one for a specific application. Instead, we train a classifier that provides a continuous set of TPR vs FPR tradeoffs based on a uniform set of features, as follows.

Given the merged VCF of a sample and its long-read BAM, we used the kanpig genotyper v0.3.1^15^ to both refine the genotypes and annotate every record with features that include genotype quality (GQ), the coverage over a window of nearby variants (DP), the coverage of reference and alternate allele (AD), the likelihood that the alternate allele is present in the sample (SQ), and a score that captures the similarity of the reads to the variant graph (KS). Specifically, we used the following kanpig parameters, that were observed to perform best on single-sample callsets^15^:

--sizemin 20 --sizemax 10000 --chunksize 1000 --gpenalty 0.02 --hapsim 0.9999 --sizesim 0.90 --seqsim 0.85 --maxpaths 10000

We verified that such features are informative by marking the TPs in each HPRC sample using Truvari bench with default parameters and --sizefilt 50 --sizemax 50k --sizemin 0 against the dipcall truth, and by drawing the ROC curve of TPR vs FPR in dipcall’s confident regions as each feature in isolation gets thresholded: the area under such curves was greater than the area under the main diagonal of a random classifier, both for calls that were ≥90% outside tandem repeats and for calls that were ≥90% inside tandem repeats, with the former class showing a consistently larger area. This held true in every SVLEN bin induced by thresholds 50, 100, 500, 2.5k, 5k and 10k bps. We trained a classifier over DP, AD, SQ, KS, GQ, GT, SVLEN and the caller support fields annotated by Truvari collapse. Specifically, we assumed that the distribution of SV annotations might vary from sample to sample, thus we trained one XGBoost classifier^16^ per sample, as follows. Let *B* be an inter-sample HPRC callset obtained by merging the dipcall VCFs of all HPRC samples using BCFtools merge, and let *A* be the merged VCF of one specific AoU sample. We labeled a subset of calls *A*’ ⊆ *A* as positive using Truvari bench against *B*, limited to dipcall’s confident regions in HG002 and with stringent parameters:

--sizemin 50 --sizemax 1000000 --sizefilt 50 --pctsize 0.9 --pctseq 0.9

and we kept the rest of *A* unlabeled (*A*’ is large in practice: 70% of *A* on GRCh38 and 65% on CHM13; this holds over several length bins). We evaluated the performance of this approach by training an XGBoost model on *A*’ and on its complement over chromosomes 6 to Y of every HPRC sample, and then running the model over the remaining chromosomes of that sample and comparing to the dipcall truth (here *B* does not contain the sample at hand). The model emits a raw score for every VCF record: thresholding such a score yielded smooth ROC curves whose area was greater than the curves traced by every single feature in isolation. Thus, we proceeded to train an XGBoost model over the entire *A*’ of every AoU sample, and to score every record in that sample. In addition to the raw score, the model outputs a version of the score rescaled to [0..1] using *A*’as a calibration set. This rescaled score is a proxy for TPR, i.e., the set of all calls with a rescaled score ≤*x* has TPR=*x* with respect to the calibration set. Since the precision of such a subset is typically inversely proportional to its TPR, we built a stringent (less sensitive, more precise) and a lenient (more sensitive, less precise) version of the merged VCF of each sample, by setting *x*=0.7 and *x*=0.9, respectively. We used the implementation in the GATK Variant Extract-Train-Score tool^17,18^, which has been previously deployed in AoU for filtering short variants with an isolation-forest model^19^.

##### Inter-sample merging

We integrated all the intra-sample-merged and filtered VCFs across the cohort using Truvari collapse with default parameters, except for --gt all --keep common which make the method not collapse calls from the same sample and choose a call with the largest minor allele count as a representative of each cluster. Regions with more than 10k total overlapping calls from all samples were skipped, to make Truvari complete in a practical amount of time: in the case of lenient filtering, this did not remove any call on GRCh38 and it removed 4.5% of distinct calls on CHM13. By virtue of our earlier normalization on the raw files, the inter-sample-merged VCFs contained no multiallelic record, and they preserved the information of which caller supported each call in each sample, and of which cluster representatives were chosen by Truvari at each step. Our final VCFs covered 1074 samples (1027 AoU samples and 47 HPRC samples downsampled to 8x) and contained approximately 600K records in the stringent GRCh38 callset and 1.2M records in the lenient callset (these numbers become 800K and 1.7M on CHM13). Of these records, 78% are such that ≥90% of their interval is covered by the tandem repeat track, both in the stringent and in the lenient callset (this number becomes 82% on CHM13). Subsetting was done by bedtools intersect -u -f 0.9 -a calls.vcf.gz -b tandem.bed.

After merging, we re-genotyped the inter-sample VCF using the reads of each sample. This allowed us to take full advantage of our large cohort, since an SV that is present in a sample S but that was not discovered in S might have been discovered in another sample, and it might have enough reads supporting it in S to be confidently assigned to S. Several calls in the inter-sample VCFs are in close proximity to other calls. For example, consider every insertion to be an interval of 20bp centered at the insertion point, and consider every other SV to be an interval that extends from the first to the last position affected by the variant, included. On GRCh38, 37% of all calls in the stringent callset and 49% of all calls in the lenient callset overlap with ten or more other calls. These numbers become 29% and 45% on CHM13. If we restrict the analysis to calls such that ≥90% of their interval is covered by the tandem repeat track, we get 43% and 59% for GRCh38 and 33% and 52% for CHM13. We again used the kanpig genotyper, since it was specifically designed to perform well on this type of input^15^. Indeed, rather than considering each call or breakpoint in isolation like several previous long-read SV genotypers^5,20–25^, kanpig builds variant graphs over windows of nearby or overlapping calls, and it chooses highest-scoring paths through multiple, sequentially-compatible calls. We used the following parameters, which were observed to perform best on large cohort-level callsets with many clustered calls^15^:

--sizemin 20 --sizemax 10000 --chunksize 500 --gpenalty 0.04 --hapsim 0.97

Based on the final genotypes, just approximately 250k records of the stringent GRCh38 callset and just 280k records of the lenient GRCh38 callset are present in the HPRC samples at 8x (these numbers become approximately 300k in both callsets on CHM13; counts were computed using bcftools view --samples-file hprc.txt --min-ac 1). We computed a lower bound on how many calls are new with respect to the HPRC assemblies, by comparing our stringent and lenient callsets to the BCFtools merge of all the dipcall VCFs using Truvari bench with default parameters (which require just ≥70% sequence and length similarity to assign a match) and allowing a record to participate in multiple matches (--pick multi). Based on this permissive definition of a match, approximately 28% of the stringent GRCh38 callset and 35% of the lenient GRCh38 callset are not found in the HPRC assemblies (these numbers become 41% and 48% on CHM13).

To evaluate the effect of filtering on our cohort-level files, we repeated the inter-sample merging process over just the 47 filtered HPRC VCFs, we considered only calls of length between 50 bp and 1Mbp, and we compared them to the BCFtools merge of all the corresponding dipcall VCFs, limited to calls of length ≥50 bp inside dipcall’s confident regions. Comparisons were made with Truvari bench, allowing a record to participate in multiple matches (--pick multi) and using --pctsize 0.9 to require ≥90% length similarity for a match; we turned off sequence similarity with --pctseq 0 to make Truvari run in a practical amount of time. In these comparisons the sample columns of the VCFs were discarded, since we only wanted to compare sets of distinct records.

On GRCh38, the Truvari collapse of our unfiltered VCFs had 54% precision, whereas the leniently-filtered callset had 80% precision and the stringent callset had 89% precision. Conversely, recall decreased from 95% in the unfiltered callset to 91% and 81% in the filtered callsets, respectively. On CHM13 precision increased from 40% to 63% and 78%, respectively, while recall decreased from 84% to 79% and 68%. For calls whose interval was ≥90% outside tandem repeats, we performed a more stringent comparison with --pctseq 0.9 added to Truvari bench to also require low edit distance to establish a match. On GRCh38 precision went from 35% in the Truvari collapse of our unfiltered VCFs, to 86% and 93%, respectively, while recall decreased from 94% to 92% and 89%, respectively. We observed similar numbers on CHM13, with precision increasing from 33% to 84% and 92% and recall decreasing from 92% to 90% and 87%.

#### Genotyping and imputation of srWGS case samples with LRS panel resources: motivation

A key question is whether smaller LRS panel callsets can be leveraged to improve sensitivity to panel variants in larger srWGS callsets. Imputation of LRS panel variants in the srWGS callsets is a possible approach. A clear precedent has been set by extant alignment-free, kmer-count-based genotyping methods such as PanGenie^26^, which has been used to impute variants from various panels in the high-coverage 1000 Genomes Project (30x 1kGP) short-read cohort of 3,202 samples^27^. For example, earlier efforts used a panel consisting of high-quality, haplotype-resolved assemblies constructed for 32 samples as part of the Human Genome Structural Variant Consortium Phase 2 (HGSVC2) project^7^; later work as part of the HPRC project expanded this panel to 44 samples^9^. Our initial goal is thus to build an analogous panel from our mid-pass LRS samples, which can then be used as a resource for genotyping and imputation methods.

Despite the successes of PanGenie in this arena, it does suffer from limitations due to the relatively high computational cost of its underlying model and implementation. Crucially, both PanGenie v1.0.0^26^ and PanGenie v2.1.1 (the most recent version available at the outset of this work) are limited to panels containing 127 samples or fewer, making our use case intractable. While even more recent versions of PanGenie (e.g., v4.1.1) now support larger panels, its high memory requirements still result in relatively high costs. Fortunately, other work^28,29^ suggests that a similar but more computationally efficient approach that combines the KAGE alignment-free, kmer-count-based genotyping method and the GLIMPSE phasing and imputation method^30^ could provide a path forward.

In this work, we extend and scale the KAGE+GLIMPSE method to enable genotyping and imputation against our panel of 1,074 AoU+HPRC samples, with a focus on increasing accuracy for SVs. Aside from scaling and engineering improvements at the workflow level, we also take advantage of the correlation of nearby variants to reduce the number of short variants required in the panel to maintain the desired level of SV accuracy.

Where possible, we leverage existing workflows and methods. We thus adopt two heuristics that have been used in previous work: 1) we assume variation in our panel can be preprocessed to yield a pangenome graph consisting of non-overlapping bubbles; bubbles may be multiallelic and individual “bubble alleles” may be composed of distinct, adjacent, and non-overlapping variant alleles^26^, and 2) we split multiallelics to biallelics and treat them naively during phasing and imputation. Although these heuristics may artificially constrain the variety of haplotypes that may be imputed against our panel and reduce performance, we empirically find that our approach yields satisfactory accuracy while maintaining a reasonable computational cost.

##### Creation of a LRS reference panel

To create a phased, haplotype-resolved reference panel with a focus on imputation of SVs, we further integrated the AoU+HPRC short and structural variant callsets, using the strict TPR=0.7 GRCh38 structural-variant callset to minimize any negative effects from false positives. We first performed read-based physical phasing of both callsets jointly using HiPhase^31^, followed by postprocessing steps to remove 1) singleton variants, 2) variants duplicated across both callsets, 3) short variants with AF < 0.5% and further than 25kb from any structural variant, and 4) variant calls originating from one or both callsets that collided at the sample level and yielded inconsistent haplotypes. After splitting multiallelic variants in the resulting integrated SNV/indel/SV callset, we then performed statistical phasing and imputation using SHAPEIT4^32^, heuristically treating split multiallelic variants as non-overlapping biallelic variants. We then used another round of postprocessing to again remove colliding sample-level variants introduced by this treatment. Finally, variant alleles in the resulting callset were converted to bubble representation. Further details of the methods for each of these stages are given below.

The resulting phased, imputed, and haplotype-resolved SNV/indel/SV reference panel of 1,074 AoU+HPRC samples covers the autosomal chromosomes and consists of 19,942,647 variant sites in bubble representation, which contain 30,918,204 variant bubble alleles; 189,382 of these bubbles are SV-length (with an absolute difference in REF and ALT length 50bp or greater) and 74,550 are further multiallelic. For comparison, the 44-sample (32-sample) HPRC (HGSVC2) panel contains 21,304,582 (15,684,910) bubble sites in the autosomes, of which 71,873 (74,650) are SV-length and 55,156 (29,988) are further multiallelic. After decomposing bubble alleles back to their original representations, the panel contains 27,143,444 distinct variant alleles, including 770,473 SVs (653,740 insertions and 117,698 deletions).

On the Terra cloud-computing platform, total cost and runtime of physical phasing (statistical phasing and postprocessing) were ~$200 (~$600) and ~3 hours (~10 hours), respectively. This relatively low computational burden allows for quick iteration of the panel and establishes an encouraging baseline for scaling to larger panels.

##### Genotyping, phasing, and imputation of srWGS case samples

Genotyping of srWGS case samples against our LRS reference panel can be performed using a modified version of KAGE, which uses counts of k-mers that are low frequency in the panel to genotype biallelic variants. We then use the resulting genotype likelihoods as input to GLIMPSE to perform phasing and imputation, again heuristically treating multiallelic sites as biallelic and performing a postprocessing step to remove colliding variants. This process thus generates phased haplotypes given srWGS case samples. Again, further details of the methods are given below.

To enable eQTL, GWAS/PheWAS, and additional quality-control analyses, we imputed against our panel 1) all 3,202 1kGP samples, and 2) 10,000 self-identified Black or African American AoU srWGS participants. Figure 17 shows unfiltered, per-sample counts of heterozygous variants, stratified by variant type and genetic ancestry group, for both the HPRC samples in our panel and the 1kGP samples. We can roughly see that the relative distribution of counts over all populations that is exhibited by the HPRC samples in our panel is recapitulated in the 1kGP cohort, despite the majority of the panel being composed of self-identified Black or African American participants. Figures 18 and 19 show panel-case allele-frequency correlations and plots of Hardy-Weinberg equilibrium for the 1kGP and AoU cohorts, respectively. Here, the effects of genetic ancestry matching become more apparent, as the latter cohort shows higher allele-frequency correlation and qualitatively appears to be closer to Hardy-Weinberg equilibrium. Nevertheless, the high allele-frequency Pearson correlation coefficients for both cohorts suggest that our panel yields good imputation results. Roughly comparing these coefficients to those found in Liao *et al.* 2023^9^ when imputing the 1kGP cohort against the 44-sample HPRC panel with PanGenie, we see that 1) our unfiltered 1kGP coefficients are comparable, and 2) our unfiltered AoU coefficients are comparable to the filtered 1kGP coefficients there.

On the Terra cloud-computing platform, total cost and runtime for the one-time building of per-chromosome KAGE kmer indices and count models from the phased panel VCF were ~$25 and ~5 hours, respectively. Subsequent costs for KAGE genotyping and GLIMPSE imputation of the 3,202 1kGP and 10,000 AoU srWGS samples against the reduced AoU+HPRC panel were typically ~24 cents and ~45 cents per sample, respectively; total runtime for the 1kGP cohort was roughly 1 day. These costs and runtimes include initial CRAM decompression per sample (which contributes a nonnegligible fraction of the cost of the genotyping step), as well as postprocessing and merging to a final cohort VCF.

Note that all quoted costs are sensitive to the time at which the workflows were run, since the rate of Google Cloud Platform VM preemption (and hence, the rate of task restarts) varies throughout the day and week. These costs could be further optimized by adjusting VM CPU/memory sizing, further reducing the number of short variants in the panel, adjusting the GLIMPSE sharding scheme or minibatch size (for example, one test GLIMPSE batch of 500 samples run with all samples in a single minibatch and sharded at ~5Mbp—rather than per-chromosome—achieved a cost of ~21 cents per sample; note also that in other studies, GLIMPSE or comparable tools are routinely run on minibatches of ~10,000 samples), eliminating KAGE genotyping by using previously derived short-variant genotypes and relying only on GLIMPSE imputation to further access SVs (see discussion in^29^), or updating from GLIMPSE to the more scalable GLIMPSE2^33^. We leave exploration of these optimizations to future work with larger panels and case cohorts.

#### Evaluations of the LRS reference panel and imputed srWGS cases

##### Leave-out evaluation of PanGenie and KAGE+GLIMPSE

As a part of preliminary work, in order to test the accuracy of our KAGE+GLIMPSE implementation and compare it against that of the PanGenie method, we performed leave-out evaluations. Such evaluations treat the panel callset as a baseline for calculating concordance metrics and are performed by: 1) creating a leave-out reference panel by removing a single or many samples from the panel callset, 2) using this reference panel in conjunction with matched short-read sequencing for the left-out sample to perform genotyping and imputation, and 3) calculating genotype concordance (at loci retained in the leave-out panel) against calls for the left-out sample(s) in the full panel. This design avoids leakage from the presence of the matched case samples in the full reference panel.

The unfiltered performance of both methods is shown in Figure 20, which summarizes: 1) in the top row, leave-one-out evaluations performed using a reference panel of 32 HGSVC2 haplotype-resolved assemblies and 10 matched 30x 1kGP srWGS cases, comparing against PanGenie v2.1.1 (which can only accommodate panels up to 127 samples), and 2) in the bottom row, a leave-many-out evaluation performed with our AoU+HPRC panel using the same 1kGP srWGS cases matching 40 of the HPRC samples in our panel, comparing against the more recent PanGenie v4.1.1 (released during the final stages of this work). Following previous work^26^, exact genotype concordance (macro-averaged across the three genotype classes) is taken as the accuracy metric (although note discussion below and in Grytten et al. 2023 on the shortcomings of this metric). Bubbles are split to biallelic and concordance is computed for each bubble allele (note that out of 2,212,411 SV-length bubble alleles in our AoU+HPRC panel, only 36,682 are entirely composed of non-SV original-representation alleles). Both sets of evaluations are performed only on chr1 for cost reasons.

In general, the performance of KAGE+GLIMPSE is comparable to or better than that of PanGenie over stratifications in allele frequency, variant length, and genomic context. Furthermore, the modular design of the KAGE+GLIMPSE workflows allows for more flexibility in sharding and runtime optimization. This suggests that KAGE+GLIMPSE is currently more suitable for our use case, as well as a strong candidate for scaling more favorably to future work.

##### Leave-out evaluation of imputation quality metrics

The second of the leave-out experiments above (i.e., a leave-many-out evaluation with the AoU+HPRC panel and 40 HPRC/1kGP cases) can also be used to compute additional metrics reflecting imputation quality, again assuming the reference-panel genotypes as a baseline and using exact genotype concordance. Figures 21 and 22 show dosage r^2^ (averaged over all 40 evaluation samples) and the non-reference concordance rate (per sample) as calculated by the GLIMPSE2 v2.0.0 concordance utility tool^33^ over chr6, stratified by variant type, allele frequency, and TR/homopolymer context. The effect of genotype-level filtering using various thresholds on the maximum genotype posterior observed at each genotype is also demonstrated. Accuracy varies over the stratifications as expected: it a) increases with minor allele frequency, b) is relatively higher for deletions and SNPs than for insertions, and c) improves outside of TR/homopolymer regions (notably, the impact of filtering is also lower here for deletions and SNPs, suggesting that the corresponding unfiltered genotypes may suffice for certain use cases). Although only a rough comparison is possible, accuracy appears to be similar to that for AFR samples exhibited by a LRS reference panel built from 888 diverse 1kGP samples^34^.

##### Evaluation against assembly-based truth

To more independently assess the accuracy of our LRS panel and the srWGS case samples imputed against it, we further performed evaluations using the same truth VCFs used for intrasample filtering described above (i.e., those generated by running dipcall on high-quality HPRC assemblies). Using this truth data in conjunction with Vcfdist v2.5.3^11^, an alignment-based benchmarking method for locally phased short and structural variants, we may evaluate the accuracy of VCFs generated by various stages of both our panel-creation and case pipelines. We can also compare the accuracy of other locally phased callsets that contain the HPRC truth samples—in particular, the Byrska-Bishop et al. panel constructed using srWGS from the 3,202 1kGP samples^27^. These comparisons are aided by the fact that Vcfdist is relatively robust to differences in variant representation, so that accuracy metrics can be roughly compared across callsets with different representations. Furthermore, in contrast to the exact genotype concordance metrics used above, metrics computed by vcfdist allow for slack in genotype matching, which may reflect a more practical view of accuracy for SVs, in particular. We ran vcfdist with the command-line arguments --realign-truth --realign-query, along with --largest-variant set to 5kb (1kb) outside (inside) of repeat/homopolymer regions for runtime/cost considerations.

The top row of Figure 23 (24) shows the resulting precision and recall over 40 HPRC truth samples in chr6 dipcall-confident regions outside (inside) of the GIAB v3.0 tandem repeat and homopolymer regions, stratified by variant type. Metrics are shown for 4 different callsets: 1) the LRS AoU+HPRC integrated panel after filtering, concatenation, and deduplication, 2) the final LRS AoU+HPRC panel after statistical phasing and imputation, collision removal, and bubble conversion, 3) srWGS 30x cases phased and imputed using a leave-out panel that drops all 40 HPRC truth samples, and 4) the srWGS Byrska-Bishop et al. 30x 1kGP panel after sequence-resolving deletions and inversions. We can draw various conclusions: 1) statistical phasing and imputation improves phased SV recall at the expense of some precision, 2) unfiltered accuracy in the srWGS case samples is comparable to that of the LRS panel itself, and 3) recall of both our LRS panel and srWGS cases is significantly higher than that of the resolved srWGS Byrska-Bishop et al. panel.

In 40 HPRC samples, the LRS panel achieved 94.0 ± 1.1% precision and 91.4 ± 1.3% recall for SVs with length <5kb in dipcall-confident, non-repeat/homopolymer regions on a single evaluation chromosome; the unfiltered matched srWGS cases achieved 95.5 ± 1.0% precision and 87.9 ± 2.0% recall when imputed against the leave-out LRS panel. In comparison, the Byrska-Bishop et al. reference panel constructed using srWGS-only methods from the 3,202 1kGP samples achieved 89.1 ± 3.0% precision and 37.0 ± 1.7% recall after resolving haplotype sequences for deletions and inversions where possible in that panel. Although overall accuracy is lower in the difficult TR/homopolymer regions that contain a majority of SVs, precision remains relatively high and roughly comparable to that for short variants in these regions. That we can achieve accuracy in srWGS that is comparable to that of the full LRS panel demonstrates the additional power that can be unlocked by using LRS resources in conjunction with readily available srWGS data.

The bottom rows of Figures 23 and 24 show the same, but now for a full AoU+HPRC panel for which no reduction has been performed (i.e., we retain all non-singleton short variants, not just those with AF > 0.005 within 25kb of an SV). Here, we see that recall of short variants in the LRS panel and srWGS cases recovers, but no significant change in SV accuracy is observed, validating our strategy of selectively filtering short variants to reduce cost while maintaining SV performance.

##### Evaluation of Mendelian consistency using 602 1kGP imputed trios

Finally, we also evaluate accuracy using Mendelian consistency on chr6 across 602 trios in the imputed 1kGP callset. The non-reference Mendelian error rate per trio is shown in Figure 25, stratified by variant type, allele frequency, and repeat/homopolymer context; the effects of filtering on by various genotype-posterior thresholds are again illustrated. Note that we return to requiring exact genotype concordance on bubble alleles split to biallelic, which is especially strict in the trio context; when filtering, we further require that all genotypes in all members of a given trio pass the filter to be included in evaluation. Nevertheless, we see that the trends in accuracy observed in the leave-out evaluations above are recapitulated here. Moreover, the effect of genetic ancestry matching is apparent in the lower error rates for AFR trios.

#### Creation of the LRS reference panel: methods

We provide additional implementation details for the procedure used to create the LRS reference panel described above. The primary inputs to this procedure are 1) the short-variant callset and the integrated, filtered SV callset; the resulting output is a phased, imputed, and haplotype-resolved SNV/indel/SV reference panel. We detail each of the stages and the evaluation of this panel-creation procedure below.

##### Physical phasing with HiPhase

The short-variant and SV VCFs are first preprocessed to unphase all phased genotypes, as is recommended for use with HiPhase. HiPhase v1.4.5 with recommended parameters (including --global-realignment-cputime 300) is then run for each sample, providing the corresponding LRS BAM as input. This results in two separate VCFs for each sample, respectively containing short-variants and SVs from the corresponding input VCFs that are now physically phased. Each of the two sets of single-sample VCFs is then merged into a cohort VCF using bcftools.

##### Filtering, concatentation, and deduplication

To maintain SV accuracy while reducing computational requirements, we strategically filter the physically phased short-variant cohort VCF. Using a combination of bcftools and GATK, we filter out short variants that are not within 25kb of a variant in the SV VCF. We also retain only short variants with AF > 0.005. Finally, we also filter out singleton SVs. The filtered short-variant and SV VCFs are then split to biallelic and concatenated with bcftools, with deduplication of exact-match alleles enabled in such a way that calls in the SV VCF are preferentially preserved over calls in the short-variant VCF. This yields a single cohort VCF that contains physically phased short variants and SVs.

##### Removing colliding variants

Imputation relies on building a pangenome where clusters of nearby variants on the same haplotype are transformed into disjoint parallel paths (one path per distinct haplotype). We say that two phased variants *collide* if they overlap on the same haplotype: in this case, reconstructing the haplotype unambiguously is impossible. HiPhase may assign colliding calls to the same haplotype if it considers them to be similar representations of the same variant. Later, SHAPEIT4 may also impute events that might overlap with those that are already present in the physically-phased haplotypes.

We remove a set of calls of smallest total weight so that the resulting haplotypes have no collision, as follows. Let a window be a maximal set of records that overlap on the reference (without taking their samples or phase into account). For each window, sample and haplotype, we compute a maximum-weight independent set of the overlap graph where every call in that haplotype is a node, and where there is an edge iff two calls overlap, using a linear-time algorithm^35^. For simplicity, we assign equal weight to every call (whether it is a SNP, indel or SV); we leave to future work experimentation with weights that are proportional to some measure of size or confidence, as well as exploration of an alternative cleaning strategy where the unit that we are allowed to remove per sample is an entire VCF record, rather than the occurrence of that variant on a specific haplotype. This would require computing a maximum-weight independent set on a graph where there is a node for every call in the sample (phased or unphased), and where there is an edge between two nodes if their calls overlap on any haplotype; in this case, the linear-time algorithm used^35^ would no longer apply.

We apply this collision-removal method on the cohort VCF produced by the previous step, yielding a physically phased, collisionless cohort VCF.

##### Statistical phasing with SHAPEIT4

We then perform statistical phasing over the autosomes using SHAPEIT v4.2.2; although SHAPEIT4 has been superseded by SHAPEIT5, we use this older version as it can take into account physical-phasing information inferred by HiPhase, which is conveyed via phase-set tags. Recall that all variants have been split to biallelic at this stage; we thus will naively treat multiallelics as biallelics, as SHAPEIT4 cannot rigorously model multiallelic sites.

We first use the GLIMPSE v1.1.1 chunk utility tool to identify genomic chunks for phasing and imputation with SHAPEIT4, selecting --window-size 5000000 --buffer-size 500000 so that the resulting chunks can be run with SHAPEIT4 in VMs with 32CPU and 128GB memory, using recommended parameters and genetic maps. After ligating chunks with the SHAPEIT 5.1.1 ligate utility tool, we perform another round of collision removal, yielding a fully phased, collisionless cohort VCF.

##### Converting to bubble representation

The final step in our panel-creation procedure is the conversion of variants to bubble representation, which involves combining adjacent variants in haplotypes until the pangenome graph consists solely of bubbles. This is accomplished using a preprocessing pipeline (<https://bitbucket.org/jana_ebler/vcf-merging/src/master/pangenome-graph-from-callset/>) recommended for use with PanGenie v3.1.0. This pipeline also removes sites with a missing fraction greater than 20% as an initial step. The final result of our panel-creation procedure is then a fully phased, imputed, and haplotype-resolved SNV/indel/SV reference-panel VCF in bubble representation over the autosomes.

#### Genotyping, phasing, and imputation of srWGS cases: methods

Once a pangenome panel in bubble representation has been created, it can be used as a resource to perform genotyping, phasing, and imputation in srWGS case samples. We now describe in more detail the corresponding methods.

##### Genotyping with KAGE

As discussed above, we selected the extant KAGE alignment-free, kmer-count-based genotyping method due to its ability to scale to the required panel size and its favorable accuracy and runtime requirements. We further developed the method by: 1) adding code functionality to enable sharding by genomic region and improve scalability, 2) refactoring method implementations and modifying choices of data structures to decrease memory and disk requirements, 3) substantially ablating unused code, and 4) developing optimized WDL pipelines for running KAGE and automated evaluations in the Terra cloud computing environment^36,37^. In particular, we forked our codebase from commit 9bcbcd0 of <https://github.com/kage-genotyper/kage>, which follows version v0.1.1 but predates version v2.0.0. Although the latter emphasizes support for SVs^29^, sufficient functionality for our purposes already exists in the forked commit. However, it is possible that version 2.0.0 may feature additional improvements (e.g., support for multiallelic bubbles) that complement those introduced in our codebase, which are thus a clear target for future work.

KAGE is an alignment-free genotyping method based on counts of low-frequency kmers, which are identified from both the pangenome panel and the reference and used to construct a kmer index over all biallelic bubbles. Reads from a case sample can then be kmerized to generate kmer counts against the index; these counts are then used to generate genotype likelihoods based on a binomial count model. In this work, we generate 31bp kmer indices on a per-chromosome basis to improve scalability, subsequently using only case reads from a given chromosome in conjunction with the corresponding index to generate kmer counts. Note that this last choice deviates slightly from the alignment-free viewpoint of the method, but we take advantage of the fact that srWGS alignments are already computed for AoU samples.

In our case pipeline, kmer counts and genotype likelihoods are generated on a per-case-and-chromosome basis, yielding single-sample, single-chromosome, unphased, genotyped VCFs over all biallelic bubbles in the pangenome panel given aligned reads from an input case BAM.

##### Phasing and imputation with GLIMPSE

The KAGE genotyping likelihoods are then used as input to the GLIMPSE phasing and imputation method. Multiallelic bubbles are also imputed at this stage, again by splitting and heuristically treating them as biallelic. On a per-chromosome basis, we run the GLIMPSE v1.1.1 phase tool on minibatches of 20-30 case samples, using the same genetic maps used with SHAPEIT4 during phasing of the panel. For each sample, we then run the GLIMPSE sample tool on each per-chromosome phased VCF to sample haplotypes for these case samples, subsequently concatenating the resulting per-chromosome VCFs using bcftools. The resulting VCFs from all case batches are then merged using Ivcfmerge^38^ hierarchically to create a single VCF for the entire case cohort. Finally, collision removal is performed, yielding a phased, imputed, and haplotype-resolved cohort VCF in bubble representation over the autosomes.

Although no additional filtering is applied in the released callsets, several site-level and genotype-level features available from various stages of the pipeline can be used for filtering, including the original XGBoost scores for the integrated SV callset, genotype-level posteriors from GLIMPSE, and population-level metrics from the imputed 1kGP and AoU callsets.

#### Variant annotation and comparison to external datasets

Variants were matched to external long-read callsets from HGSVC [cite] and HPRC [cite] using truvari bench with parameters

--pctseq 0.7 --pctsize 0.7 -r 500 --pctovl 0 --pick multi

Calls from HPRC were generated with dipcall on the provided assemblies [cite]. Variants were additionally matched to SV calls generated on the same samples from the short-read data using truvari bench as follows. Short-read DEL and INV calls were matched with parameters

--pctseq 0.5 --pctsize 0.5 --pctovl 0.1 --pick multi -r 10000 --chunksize 10000

(with -r and --chunksize both increasing to 20000 for short-read variants over 5kb, which are primarily depth-based with lower accuracy breakpoints), using the reference to fill in sequences for short-read calls. Short-read DUPs were matched with parameters

--pctseq 0.5 --pctsize 0.5 --pick multi --dup-to-ins -r 10000 --chunksize 10000

(with -r and --chunksize both increasing to 20000 for short-read variants over 5kb), with an additional custom filter to only retain short-read variants whose start sites were within +/- SVLEN of the start site of the matched long-read variant. This breakpoint flexibility for DUPs was intended to account for the typically lower breakpoint resolution of DUP calls in the short-read callset, while retaining specificity with the --pctseq 0.5 parameter. Short-read INS calls, which are not sequence-resolved, were matched with parameters

--pctseq 0 --pctsize 0.1 -r 100 --pctovl 0 --pick multi --dup-to-ins

Finally, for all matched short-read-to-long-read comparisons, variants were required to be called in at least one overlapping sample. Variants were further annotated by genomic context if their position was overlapped by at least 90% with GIAB-defined regions ‘GRCh38_segdups_gt10kb’, ‘GRCh38_AllTandemRepeatsandHomopolymers_slop5’, and ‘GRCh38_allOtherDifficultregions’.

#### Detection and annotation of mobile element insertions

Mobile element insertions (MEIs) were detected by running PALMER^39^ for *Alu*, LINE-1, and SVA MEI types on the GRCh38-aligned BAMs for each sample. Each MEI 2.call was required to have support from at least one fully-spanning read. Calls for each MEI type were converted to VCF format with a custom script, and then merged across the cohort using truvari collapse with parameters "-k common --refdist 50 --pctseq 0 --pctsize 0". Separately, the sequences of each insertion in the integrated SV callset were annotated with RepeatMasker. Insertions in the integrated SV callset were annotated with MEI calls if they met the following criteria: 1) the Repeatmasker annotation of the insertion contained a matching MEI class (*Alu*, LINE-1, or SVA) to a PALMER MEI call within 50 bp; 2) for *Alu* and LINE-1 elements, the insertion from the SV callset and associated MEI call from PALMER had lengths that were within 80% of one another; this requirement was waived for SVAs, whose PALMER length estimates were unreliable; 3) the insertion from the SV callset and the associated MEI call from PALMER were detected in at least one shared sample. The integrated SV callset included multiple entries for insertions at the same site with substantially different lengths, which in some cases resulted in multiple entries for the same MEI type at the same site. Therefore, for the purpose of MEI analyses, the MEI-annotated insertions in the SV callset were split by MEI type and additionally merged using truvari collapse with parameters

-k common --refdist 50 --pctseq 0 --pctsize 0

For allelic heterogeneity analysis of SVAs, intrasample-merged and filtered SV calls for each individual were annotated with MEI calls as above. MEI-annotated insertions were combined across the cohort using ‘bcftools merge,’ which preserves all sequence variability, and further grouped by position using ‘bedtools cluster -d 1.’

#### *CYP2D6* variation

##### Assembly sequence extraction

Hifiasm haplotype-resolved assemblies from the 1,027 samples were obtained from the All of Us v7 data release. The assembly for each haplotype was aligned to T2T-CHM13v2.0 using minimap2 (v2.17) with the parameters

-x asm20 -m 10000 -z 10000,50 -r 50000 --end-bonus=100 -O 5,56 -E 4,1 -B 5 --secondary=no -a -t 4 --eqx -Y

Sequences were extracted corresponding to the full T2T-CHM13v2.0 *CYP2D6-7* region (chr22:42605990-42623972) with a maximum of 20 kbp of flanking sequence extending 3’ of *CYP2D6* and 5’ of *CYP2D7*, or to the end of the haplotig. Haplotig to reference coordinates were mapped with pysam (v0.22.1) get_aligned_pairs() and extracting haplotig sequences for the coordinates with SAMtools (v1.9) faidx. Haplotypes lacking a haplotig covering the full locus or with multiple haplotigs covering the full locus were filtered out. Single haplotigs with multiple alignments over the locus were retained.

##### CNV/ SV k-mer plotting

Haplotig sequences corresponding to *CYP2D6* and *CYP2D7* were divided into 100bp k-mers and aligned to *CYP2D6*, *CYP2D7*, and *CYP2D7*-spacer T2T-CHM13v2.0 reference sequences using minimap2 (v2.24) with the parameters

-a -Y --rmq=yes -k19 -w19 -g10k -A2 -B4 -O6,26 -E2,1 -s200

Based on the T2T reference, each k-mer was assigned as a best match to *CYP2D6*, *CYP2D7*, *CYP2D7*-specific spacer sequence, both *CYP2D6* and *CYP2D7*, or none of these, based on MAPQ score. The resulting alignments were plotted and visualized (see Code Availability) in order to classify copy-number changes and hybrid alleles. This pipeline was tested on 47 high-quality phased HPRC assemblies, for which the copy number of *CYP2D6* and *CYP2D7* showed 100% concordance to reported HPRC haplotypes, disregarding hybrid alleles and benchmarking samples^9^.

##### Star allele and phenotype assignment

GRCh38 reference sequences per star allele were downloaded from PharmVar and aligned to each haplotype sequence using minimap2 (v2.24) with the parameters

-a -Y --rmq=yes -k19 -w19 -g10k -A2 -B4 -O6,26 -E2,1 -s200 -N 2000 --MD --eqx --sam-hit-only

A best-matching star allele *S* was determined for each non-CNV haplotype by maximizing the alignment score, as calculated from the number of matching bases (*n*), mismatching bases (*m*), insertions (*i*), and deletions (*d*) in the CIGAR string as *n - m - i - d*, where the haplotig contains all variants defining *S*, and the number of variants corresponding to *S* present in the haplotig is equal to or exceeds that of any other suballele. Diplotypes and phenotypes were derived according to CPIC guidelines for samples for which both haplotypes were fully resolved and had a canonical *CYP2D6*-*CYP2D7* structure.

##### Star allele validation

Short read haplotyping from CRAM files for the 1,027 samples was completed using validated haplotype callers Aldy v4.4^40^ with command

aldy genotype --param min_avg_coverage=0 -p wgs -g CYP2D6 -o {out} {input_file}

Cyrius v1.1.1^41^ with

cyrius -m {manifest} -p cyrius_{sample_id} -g 38 -o {out}, PyPGx v0.20.0^42^ with pypgx create-input-vcf --assembly GRCh38 --genes CYP2D6 variants.vcf.gz Homo_sapiens_assembly38.fasta {bam_file}

pypgx prepare-depth-of-coverage --assembly GRCh38 --genes CYP2D6 depth.zip bams.txt

pypgx compute-control-statistics --assembly GRCh38 EGFR control.zip bams.txt

pypgx run-ngs-pipeline --variants variants.vcf.gz --depth-of-coverage depth.zip --control-statistics control.zip --assembly GRCh38 CYP2D6 results

and StellarPGX v1.2.7^43^ with

nextflow run main.nf -profile standard --build hg38 --gene cyp2d6

A consensus call was calculated as a composite value of the haplotype call from all callers based on majority agreement between callers (minimum 2). An independent short read analysis in which PyPGx (v0.25.0) was run equivalently except using control gene VDR produced the same PyPGx calls.

Our long read assembly-based star allele calling pipeline was validated by comparing results for the 47 Year 1 HPRC assemblies to the short read consensus calls for the 40 1KGP high-coverage samples in the same set. The short read calls matched our calls for 95% of unambiguously assigned non-SV haplotypes, and in cases of disagreement our call was supported by at least one caller. Short and long read calls were generated for AoU samples using the same methods.

##### Missense variant analysis

Coding sequence and protein prediction was performed by lifting over T2T-CHM13v2.0 gene structure coordinates downloaded from NCBI with rustybam (v0.1.24), and running Augustus (v3.5.0) separately on each haplotype sequence with options --species=human --codingseq=on, passing the lifted over gene structure as the --hintsfile. Missense variants were identified by aligning the predicted haplotype coding sequences to the GRCh38 coding sequence by running minimap2 with the above parameters, inferring amino acid sequences, mapping haplotype to reference coordinates with pysam (v0.22.1) get_aligned_pairs(), and extracting missense variants as single bases that differ from the aligned reference base at the same position in the codon, resulting in a different amino acid. Amino acid changes beyond the star allele were defined as changes not listed in the Variant Impact field of the PharmVar VCF for the assigned suballele.

### SUPPLEMENTARY NOTES

#### Supplementary Note 1: Noncanonical and rare *FMR1* and *HTT* interruptions

Examining noncanonical and rare repeat interruptions, we first noted a rare *FMR1* CGG repeat with five AGG interruptions (Fig. 3b). Such alleles are infrequently observed but thought to remain intergenerationally stable even at near-premutation lengths^44^. We catalogued 255 non-canonical *FMR1* haplotypes and 158 non-canonical *HTT* haplotypes, compared to 1,498 and 1,786 canonical haplotypes, respectively. Most of these are likely attributable to homopolymer-related sequencing errors, which is supported by the fact that of 17 *FMR1* and 10 *HTT* noncanonical haplotypes in the 50 samples with matched ONT data, none were confirmed by LongTR^45^ run on the ONT reads. However, we did confirm one previously reported^46^ CTG interruption in a 43-repeat *FMR1* allele using long-read evidence.

### SUPPLEMENTARY FIGURES

#### Supplementary Fig. 1: Overview of the AoU long-read variant discovery, reference panel construction, and imputation pipelines.

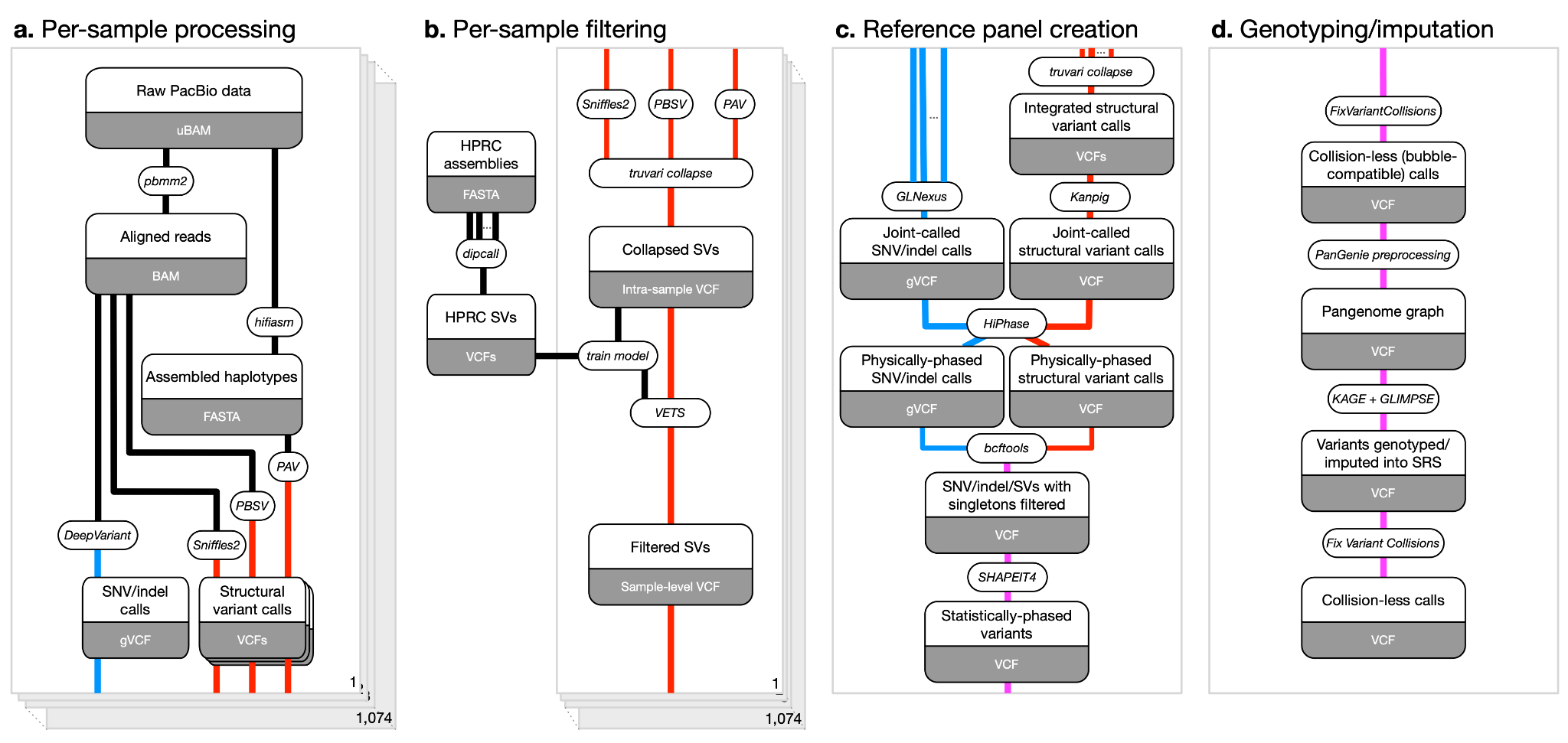

**a,** Per-sample processing of raw PacBio HiFi data. Reads were aligned with pbmm2, assembled with hifiasm, and variants were called using DeepVariant (for SNVs and indels), and three structural variant (SV) callers: Sniffles2, PBSV, and PAV. **b,** Per-sample SV filtering. Calls were first merged within each sample using truvari collapse, then scored using a per-sample XGBoost model trained on HPRC dipcall labels. Calls were filtered using score thresholds calibrated to target true positive rates (TPR), producing both lenient (TPR=0.9) and stringent (TPR=0.7) sample-level SV callsets. **c,** Reference panel creation. Sample-level SNV/indel and SV callsets were merged across samples using GLnexus and Truvari, respectively, and re-genotyped in LRS participants with Kanpig. Joint calls were phased using HiPhase, followed by concatenation, singleton filtering, and statistical phasing with SHAPEIT4 to generate a haplotype-resolved reference panel. **d,** Genotyping and imputation. The phased reference panel was transformed into a pangenome graph using PanGenie preprocessing, with removal of overlapping alleles (Fix Variant Collisions). Variants were imputed into short-read samples using KAGE and GLIMPSE. A final overlap removal step was applied to the imputed calls to resolve collisions in short-read haplotypes.

#### Supplementary Fig. 2: Median Mendelian discordance rates of SVs for three trios on reference GRCh38

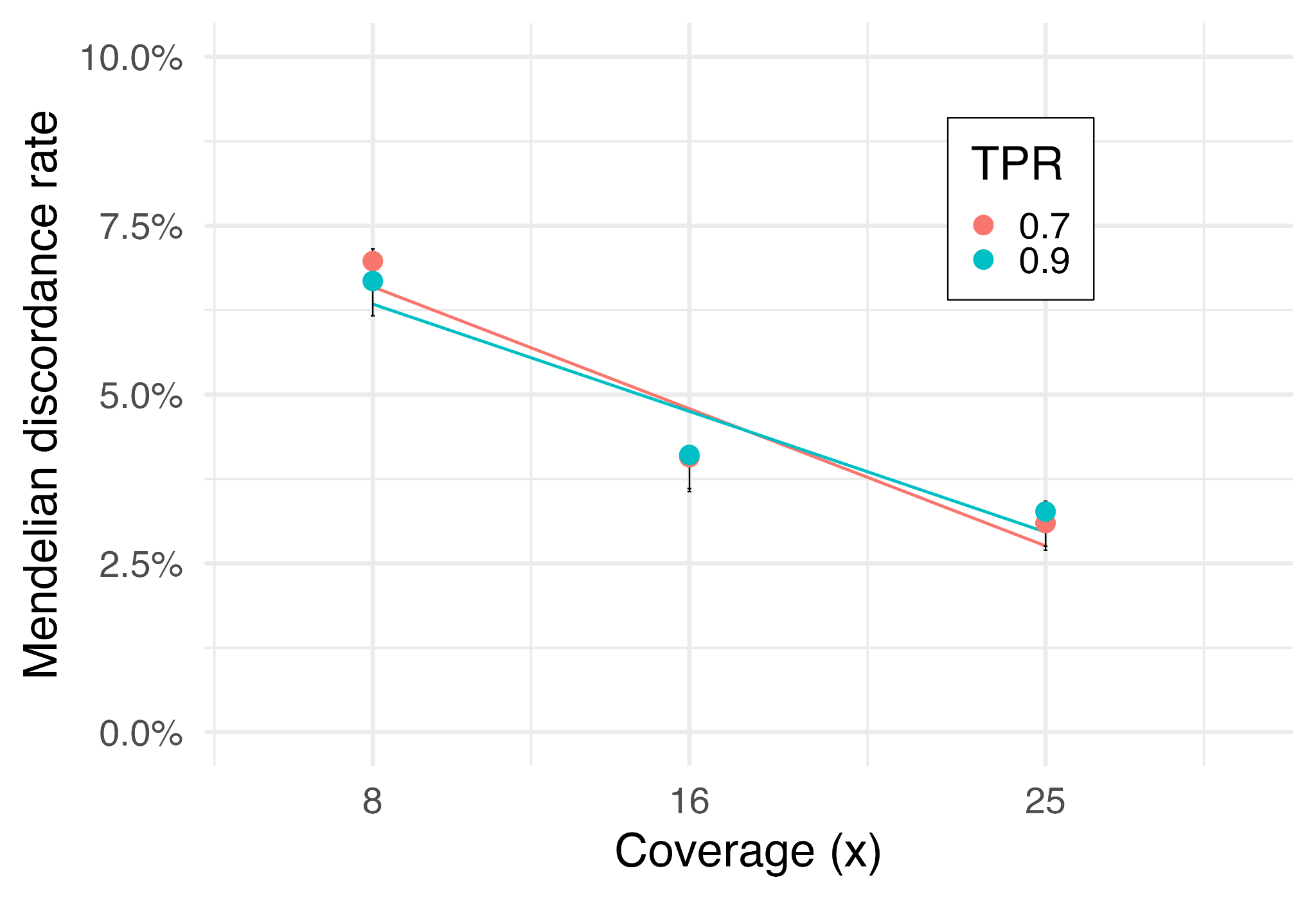

Median Mendelian discordance rates of SVs for three trios on reference GRCh38, stratified by coverage level and SV filtering stringency. Error bars represent 25th and 75th percentiles, and lines represent a linear model of median discordance rate against coverage.

#### Supplementary Fig. 3: Median Mendelian discordance rates of SVs for three trios on reference T2T-CHM13

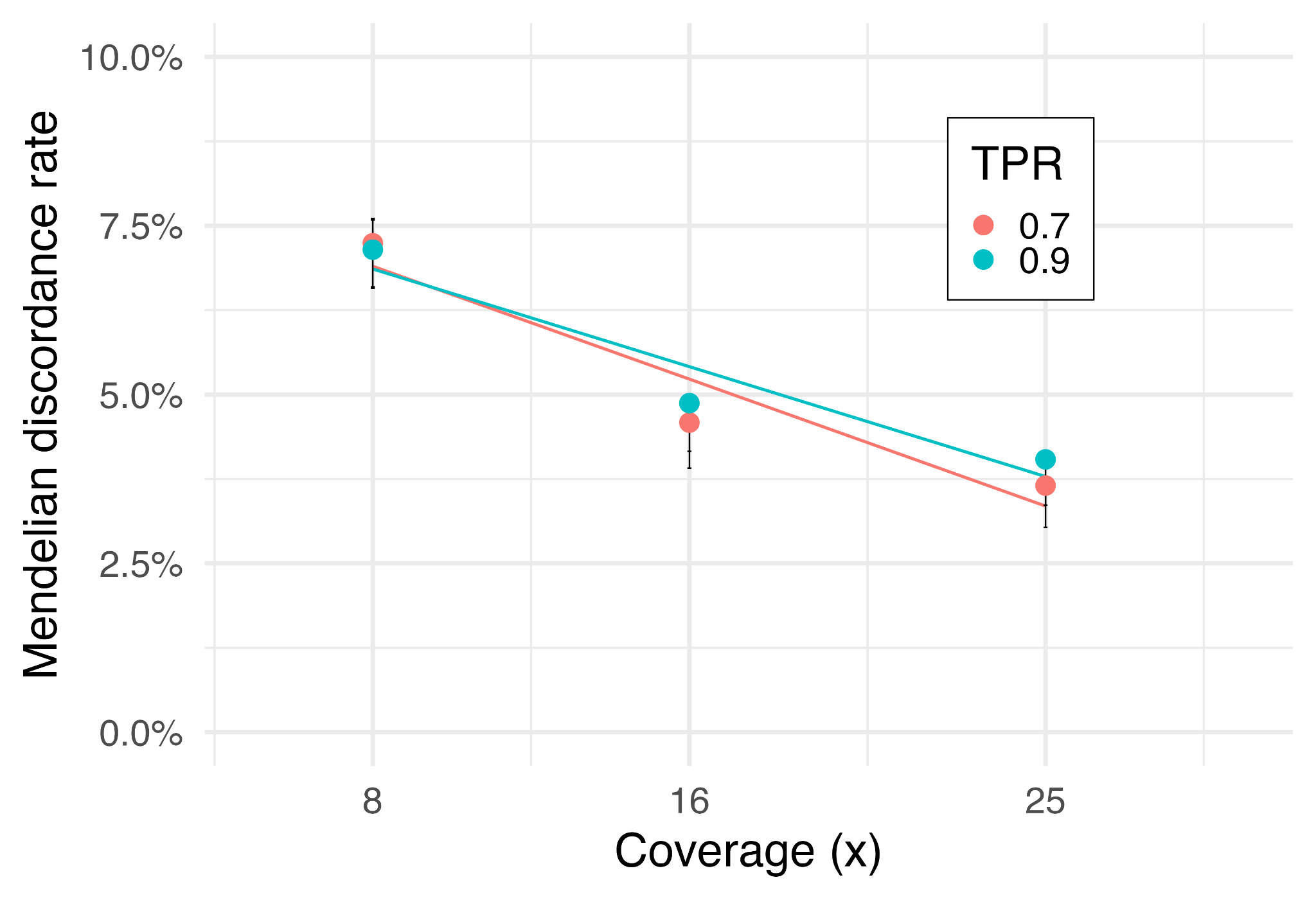

Median Mendelian discordance rates of SVs for three trios on reference T2T-CHM13, stratified by coverage level and SV filtering stringency. Error bars represent 25th and 75th percentiles, and lines represent a linear model of median discordance rate against coverage.

#### Supplementary Fig. 4: Median Mendelian discordance rates of SVs for three trios, stratified by child coverage level, SV filtering stringency, and genome context.

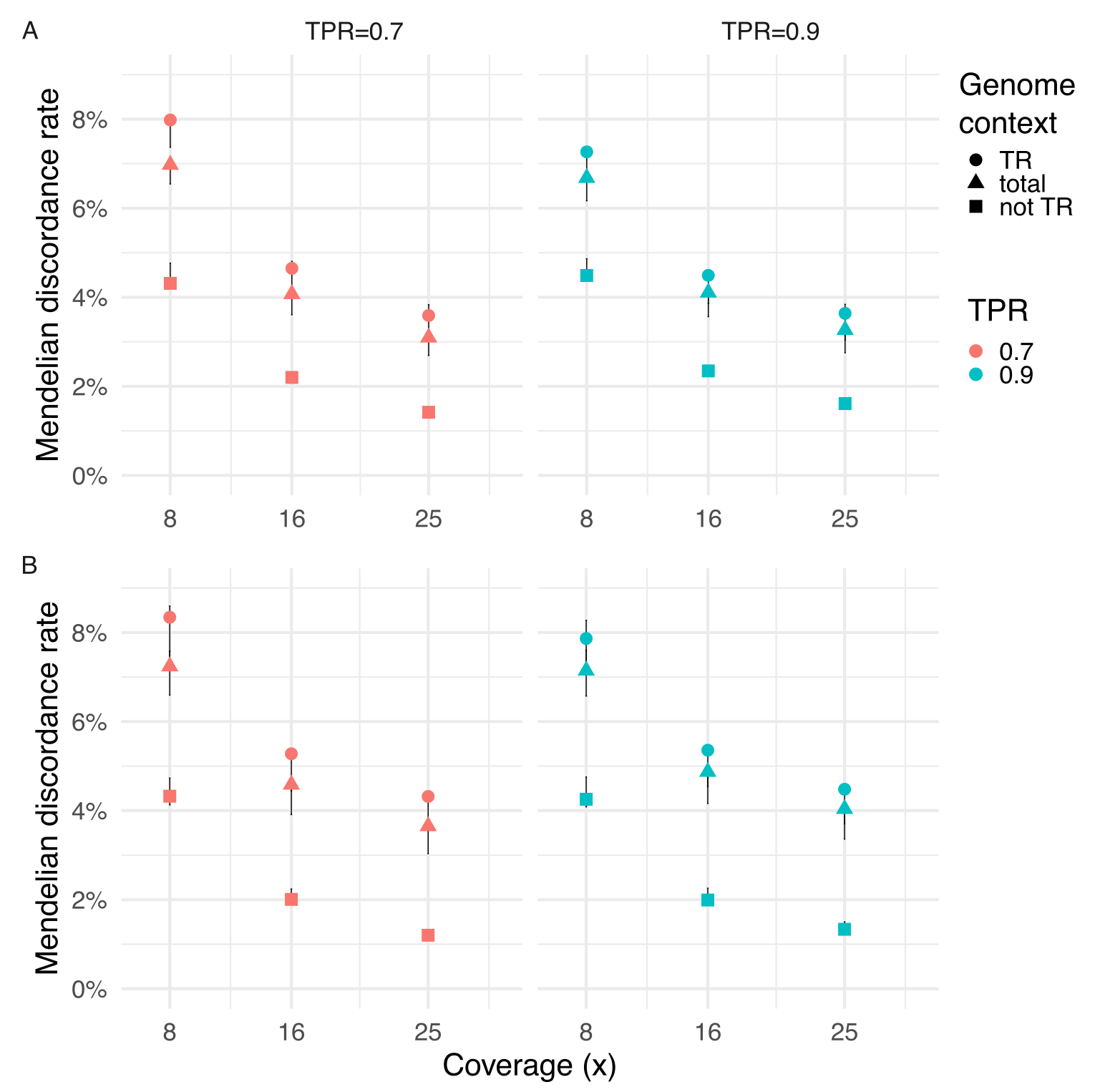

Median Mendelian discordance rates of SVs for three trios, stratified by coverage level, SV filtering stringency, and genome context. Error bars represent 25th and 75th percentiles. TR, at least 90% of the variant falls within an annotated tandem repeat (circle); not TR, all other variants not meeting the TR criteria (square); total, all variants (triangle). **a,** Analysis performed on GRCh38 reference. **b,** Analysis performed on T2T-CHM13 reference.

#### Supplementary Fig. 5: Mobile element insertions and allelic heterogeneity in SVAs in the lenient SV callset.

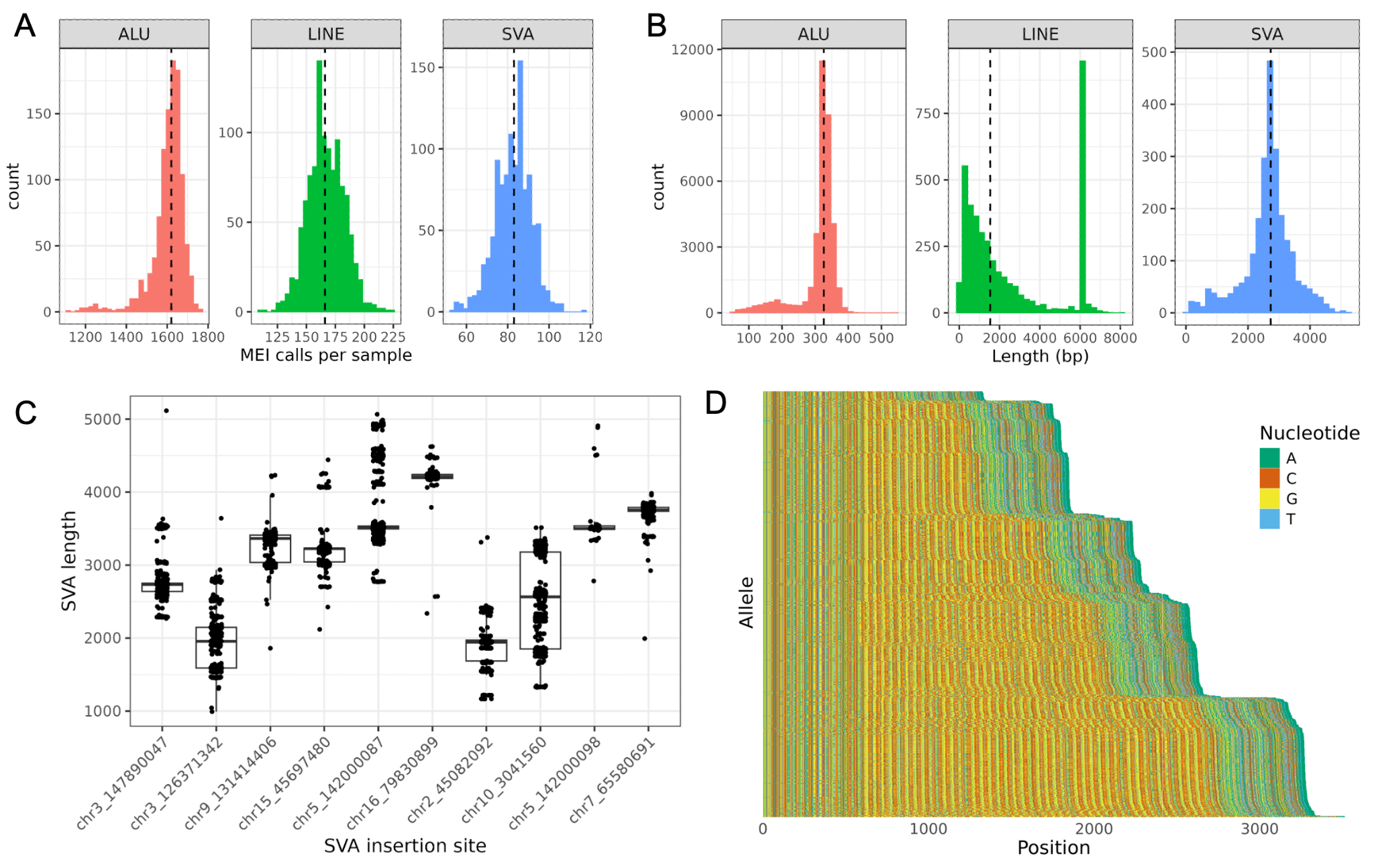

**a,** Number of MEIs per sample across the three detected classes (*Alu*, LINE-1, and SVA). **b,** Length distribution across the three MEI classes. **c,** Length distribution of SVAs at the top ten most length-variable sites. **d,** Example nucleotide composition of the SVA insertion at chr10:3,041,560.

#### Supplementary Fig. 6: Population-level analysis of genotyped structural variant callset

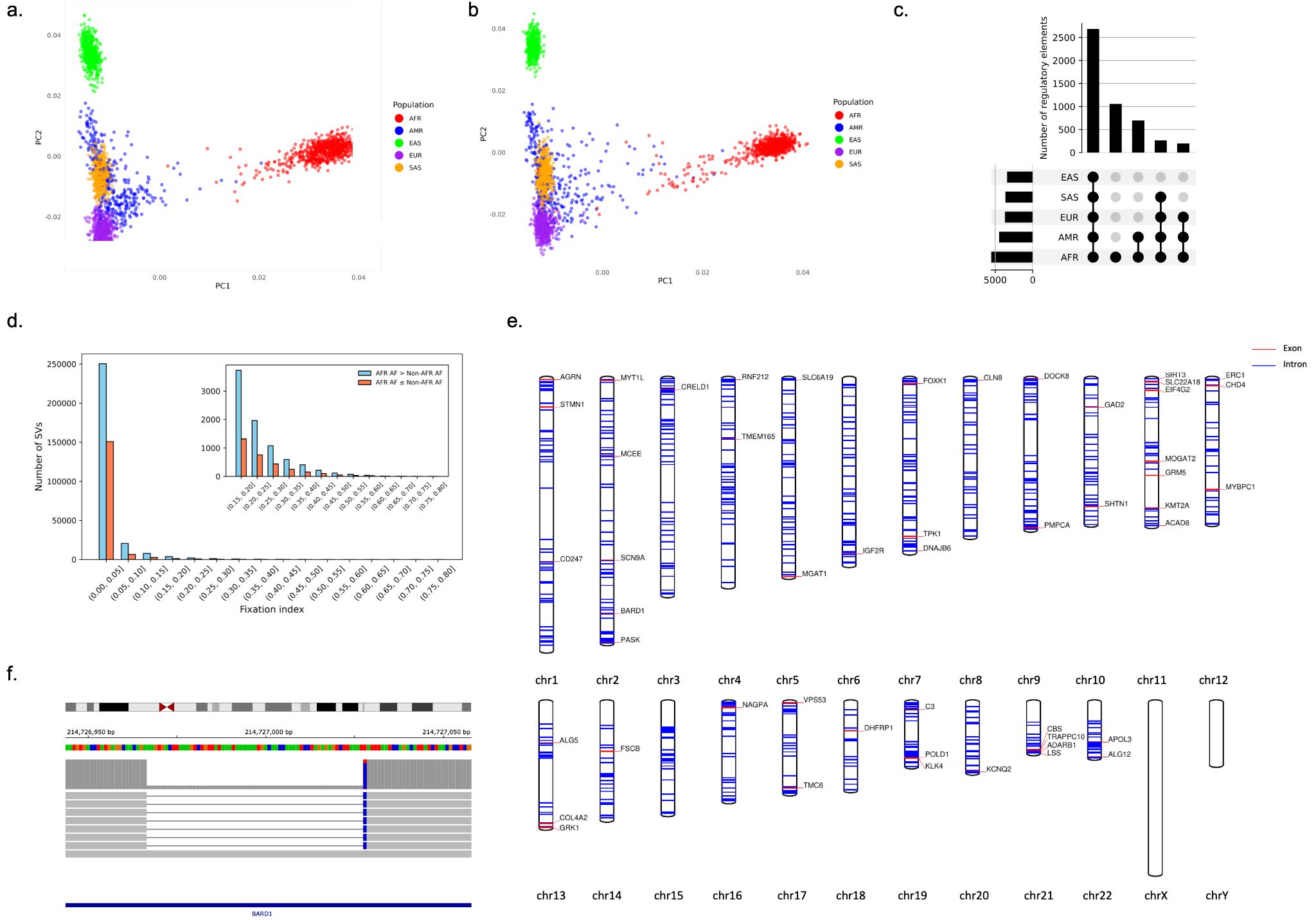

**a,** Principal Component Analysis (PCA) of SVs imputed in 2,504 unrelated samples from the 1000 Genomes Project. **b,** PCA of SNPs from the same 2,504 samples in the 1000 Genomes Project. **c**, UpSet plots show the number of regulatory elements intersected by an SV across the five continental groups in 1KGP. **d**, SV distribution as a funciton of F_st_. Sky blue represents SVs with higher allele frequencies (AFs) in individuals of African genetic ancestry (AFR) than in individuals of non-African genetic ancestry (Non-AFR), and coral represents SVs with higher AFs in non-African ancestry groups. **e**, Genomic locations of SVs intersecting high-priority genes associated with inherited diseases ^47^. Red lines represent SVs overlapping gene exons and blue lines indicate overlaps with gene introns. Gene names are labeled for exon intersections. **f,** Integrative Genomics Viewer (IGV) screenshot of a 61bp bp insertion in the exon of the *BARD1* gene.

#### Supplementary Fig. 7: Repeat expansion at ATXN3

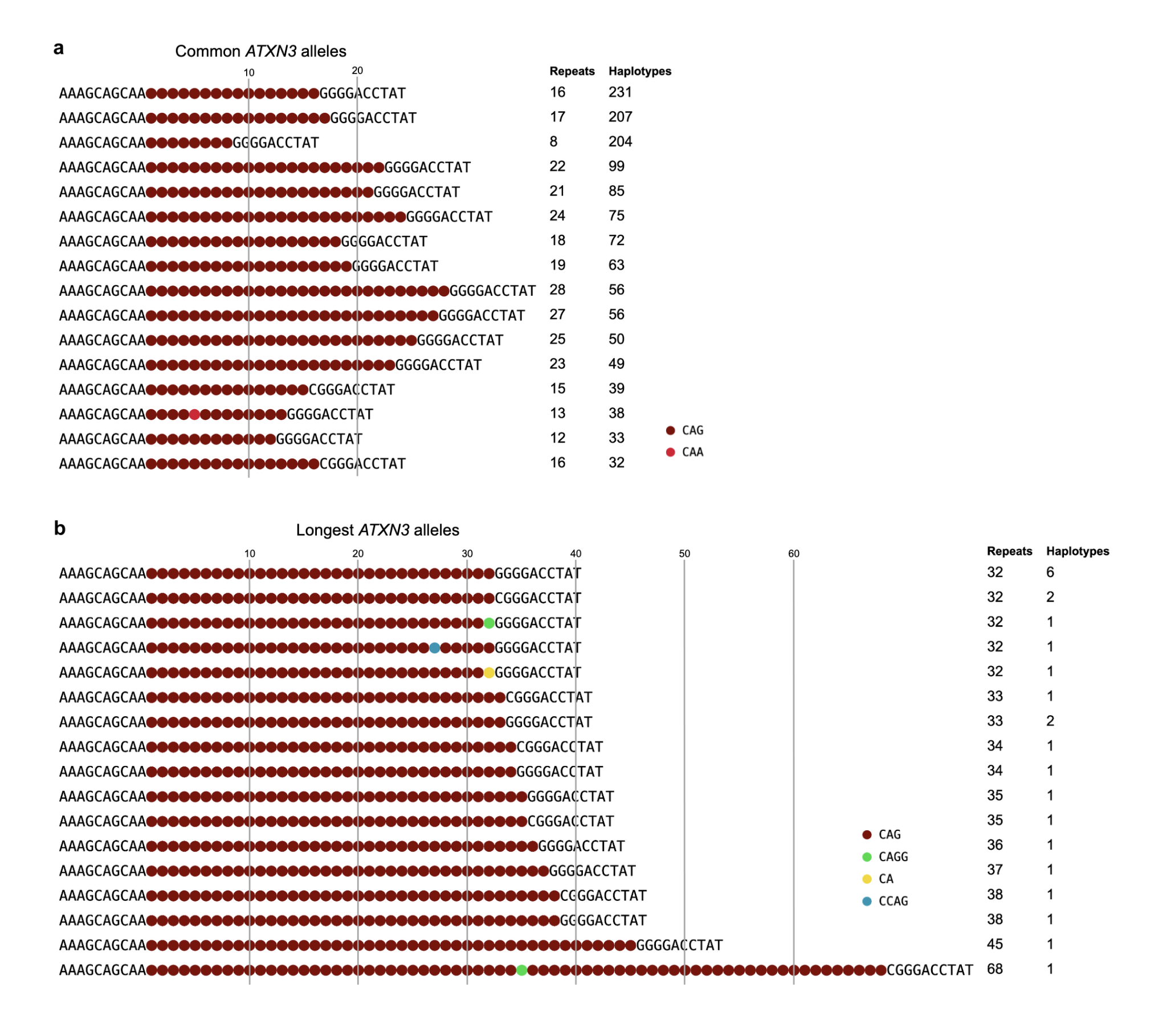

**a,** Most common and **b,** longest *ATXN3* repeat alleles. Sequences were extracted from 2,019 long read haplotypes with TRGT and plotted according to repeat and interruption patterns. The high-penetrance pathogenic expansion range is >55 CAG repeats.

#### Supplementary Fig. 8: Functional annotation and gene associations of AoU SVs

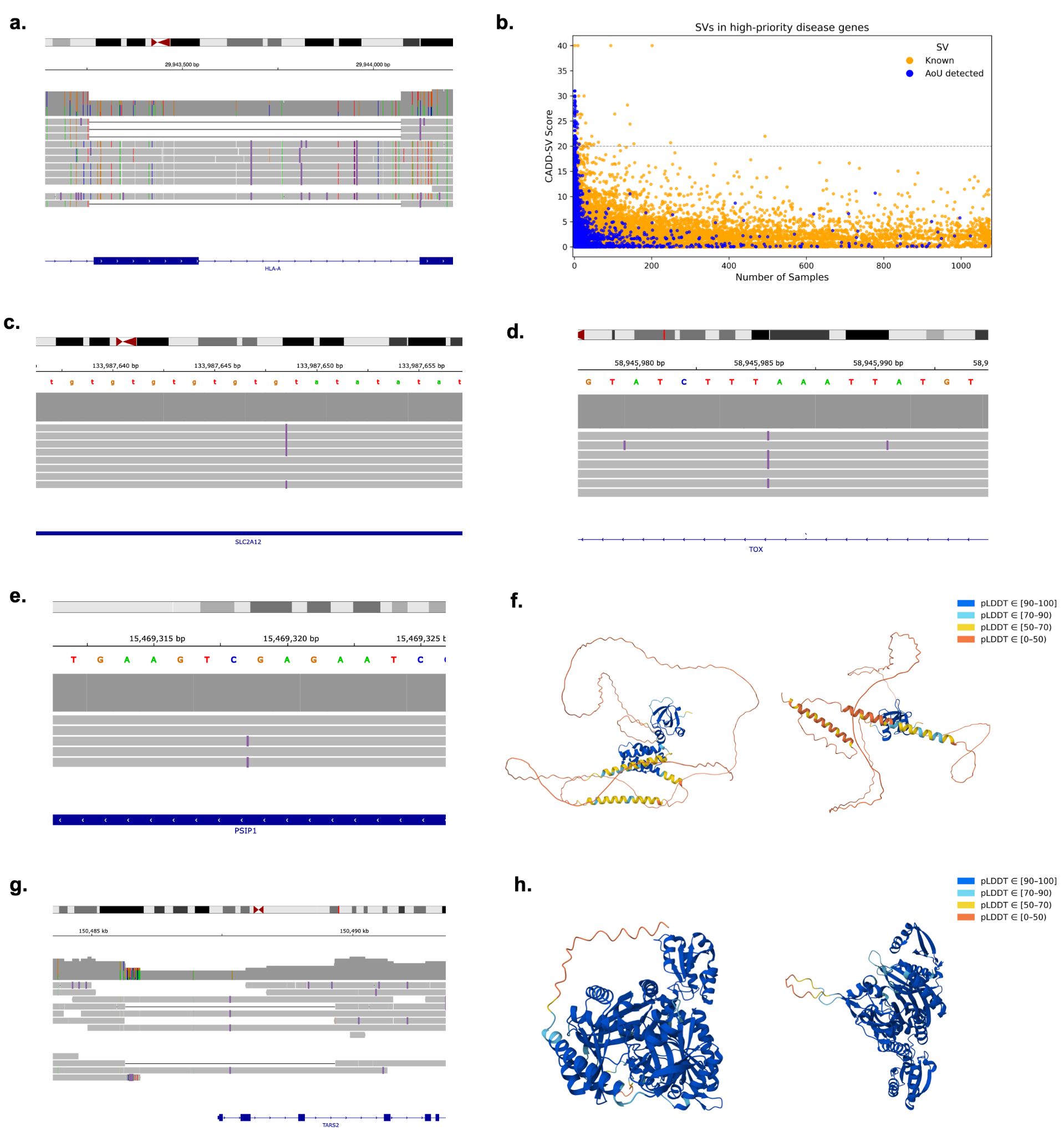

**a**, Integrative Genomics Viewer (IGV) screenshot of a 816 bp deletion intersecting *HLA-A* gene. **b,** CADD-SV score distribution of SVs overlapping genes associated with inherited diseases ^47^ in the AoU strict cohort. The x-axis indicates the number of samples containing the SV, and the y-axis shows the PHRED-scaled CADD score. “Known” SVs are those identified in at least one of the 1000G-ONT, HGSVC, or HPRC datasets ^7,9,12^, while "AoU detected" SVs are those absent in the dataset. The dashed gray horizontal line denotes the score threshold above which SVs are considered likely pathogenic. **c–e,** IGV screenshots of representative high-scoring SVs, including a 52 bp insertion intersecting *SLC2A12* (**c**), a 536 bp insertion within *TOX* (**d**), and a 226 bp deletion in the coding region of *PSIP1* (**e**). **f**, Predicted structure of PSIP1 protein from AlphaFold, showing the reference (left) and the truncated structure resulting from the SV (right). **g**, IGV screenshot of a 4,025 bp deletion in the exon of *TARS2* gene. **h**, Predicted structure of TARS2 protein from AlphaFold: reference (left) and truncated structure (right).

#### Supplementary Fig. 9: Population patterns and functional examples of SV-eQTLs

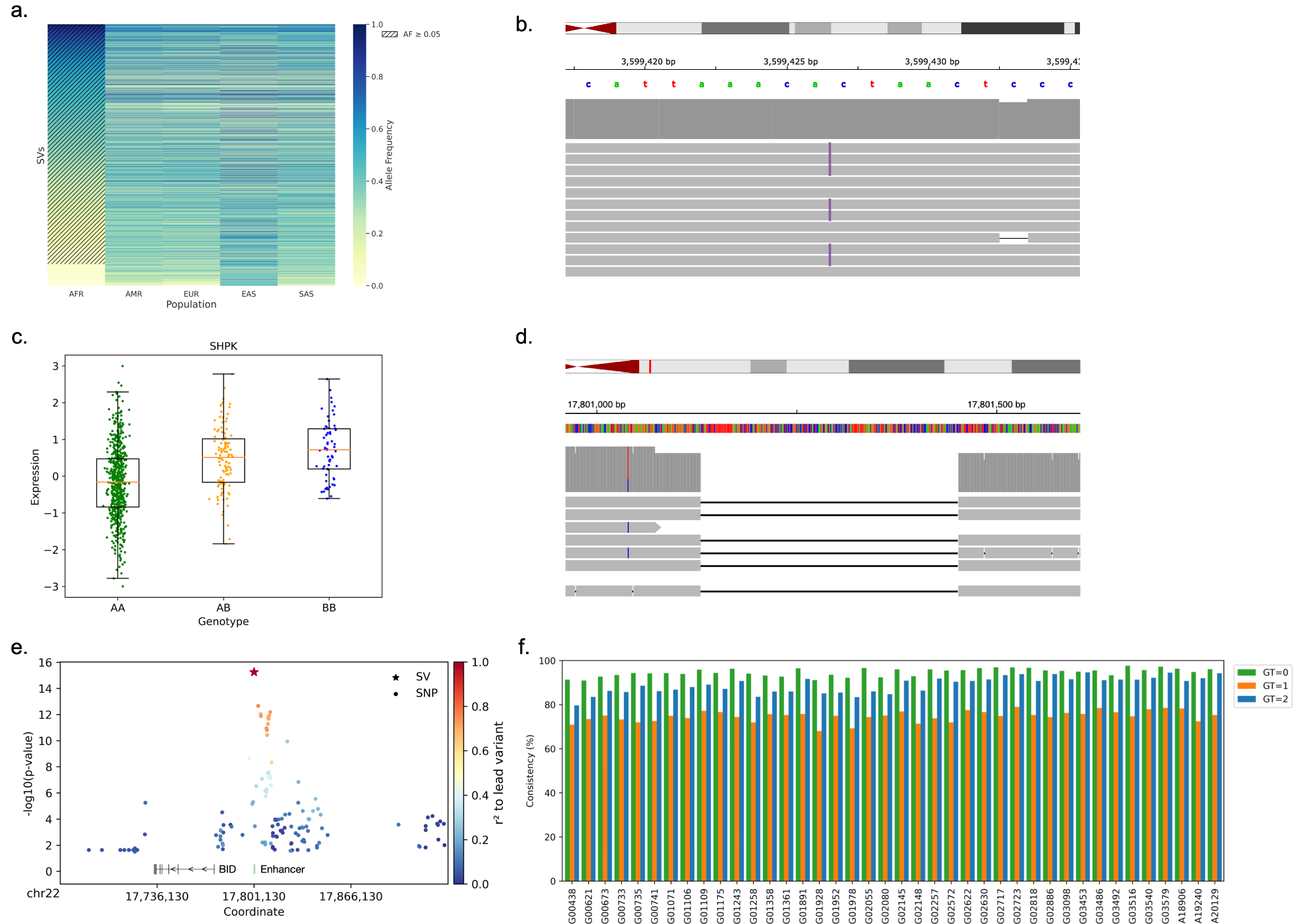

**a**, Heatmap of allele frequencies for SVs in significant SV-eQTLs, with each row representing the allele frequency of a SV across five superpopulations. Rows are sorted by descending frequency in African populations. Common SVs in Africans (AF ≥ 0.05) are marked with hashes ("///"). **b,** Integrative Genomics Viewer (IGV) screenshot of a 512 bp insertion located downstream of the 3′ transcription terminal site (TTS) of the *SHPK* gene. **c**, Distribution of genotypes and gene expression levels in 731 samples for the *SHPK*-associated deletion, with q-value of 1.31 $\times$ 10^-14^. **d**, IGV screenshot of a 322 bp deletion upstream of *BID* gene. **e**, Manhattan plot of the 322 bp deletion and nearby SNVs negatively associated with *BID* expression, with log_10_ P values. The deletion is the top variant, and points are colored by their linkage disequilibrium (r^2^) with this SV. The enhancer overlapping the SV is shown in green. **f**, Proportion of genotypes matching between short- and long-read SV calls for SV-eQTLs in the 40 common 1KG samples. Consistency is separated by the short-read genotype: 0 (green), 1 (orange), and 2 (blue). Mean genotype concordance: GT=0, 94.7%; GT=1, 74.8%; GT=2, 89.0%.

#### Supplementary Fig. 10: SVs in linkage disequilibrium with GWAS variants associated with diseases and traits

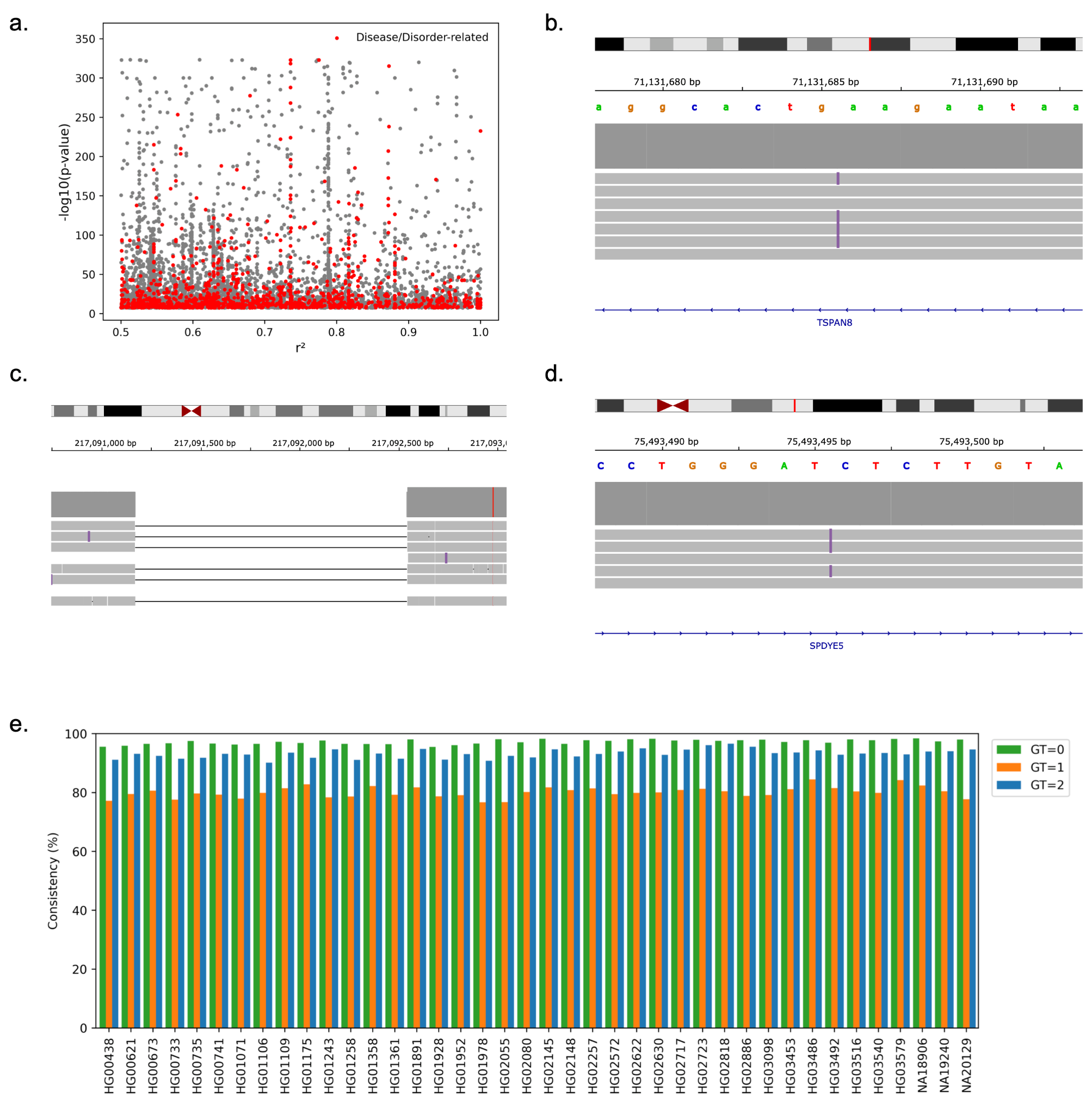

**a**, Association between GWAS trait significance and LD values between GWAS variants and our SVs. The x-axis represents the computed LD (r^2^) value for each GWAS variant-SV pair. The y-axis shows the corresponding GWAS trait p-value for the SNP, scaled as -log10. Red dots indicate traits classified as disease- or disorder-related. Only variant pairs with LD (r^2^ ≥ 0.5) are included. **b**, Integrative Genomics Viewer (IGV) screenshot of a 321 bp insertion in *TSPAN8* in LD with rs11178649, associated with hypothyroidism. **c**, IGV screenshot of a 1375 bp deletion in high LD with rs16856925, associated with breast cancer. **d**, IGV screenshot of a 63 bp insertion in high LD with six GWAS variants. **e**, Proportion of genotypes matching between short- and long-read SV calls for SVs in LD with GWAS variants in the 40 common 1KG samples. Consistency is separated by the short-read genotype: 0 (green), 1 (orange), and 2 (blue). Mean genotype concordance: GT=0, 97.2%; GT=1, 80.1%; GT=2, 93.2%.

#### Supplementary Fig. 11: Population structure and evaluation of SV genotypes

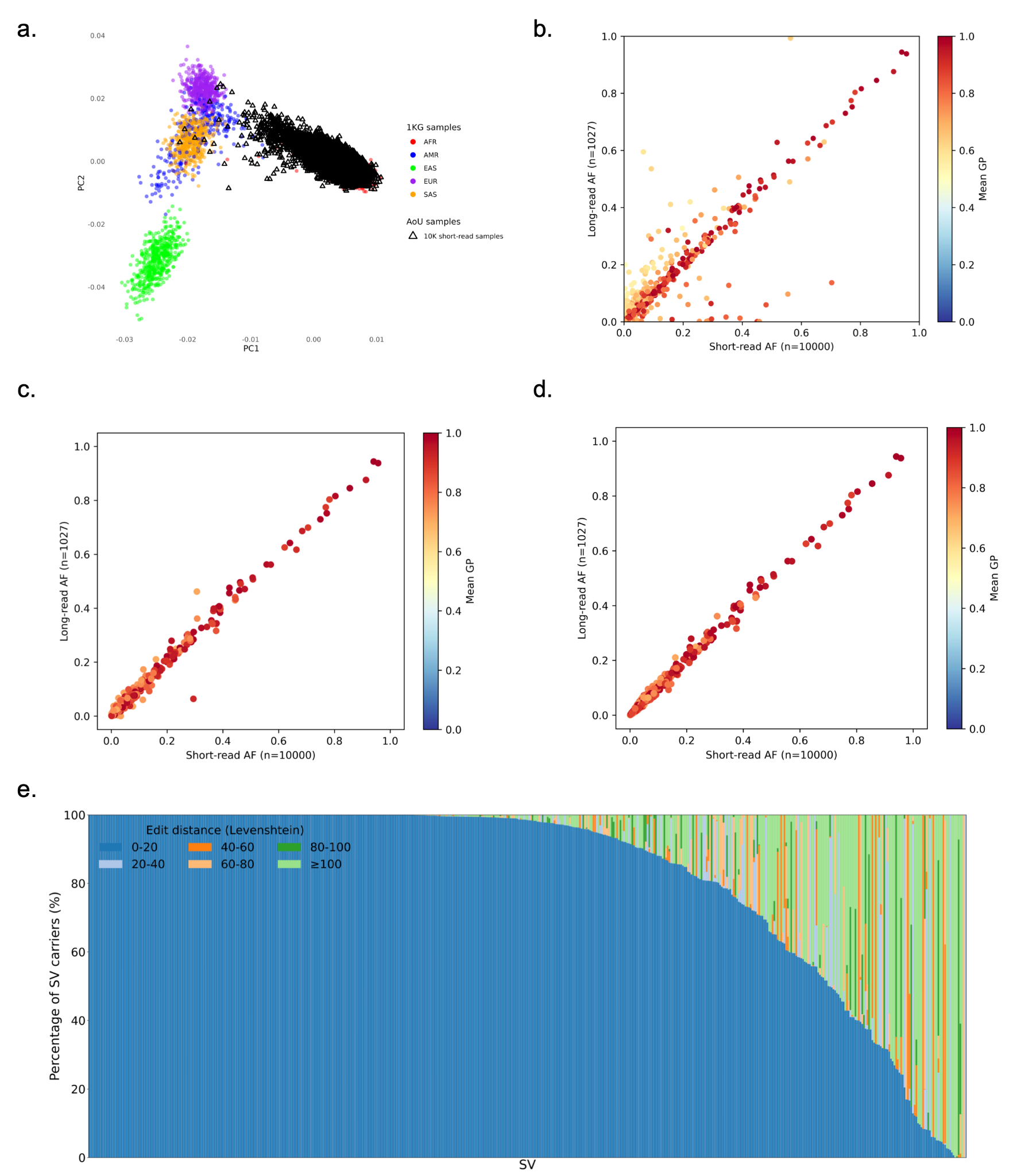

**a**, Principal component analysis (PCA) of SV genotypes from 2,504 unrelated samples in the 1000 Genomes Project and 10,000 short-read samples from the AoU cohort. **b–d**, Comparison of allele frequencies (AFs) between 1,027 AoU long-read samples and 10,000 AoU short-read samples for (b) all disease-associated SVs (n = 1,460), (c) SVs with mean genotype posterior (GP) ≥ 0.7 and Hardy–Weinberg equilibrium (HWE) ≥ 1×10^-5^ (n = 704), and (d) SVs further restricted to those with AF ratios within 1.5-fold between short- and long-read datasets (n = 546). Color indicates the mean GP per variant. **e**, Proportion of SV carriers by edit distance (Levenshtein) between imputed alleles (KAGE + GLIMPSE) and alleles generated by Locityper, shown per variant. 399 SVs have ≥70% of carriers with allele differences <20 between imputed and Locityper calls.

#### Supplementary Fig. 12: Evaluation of SV–disease associations

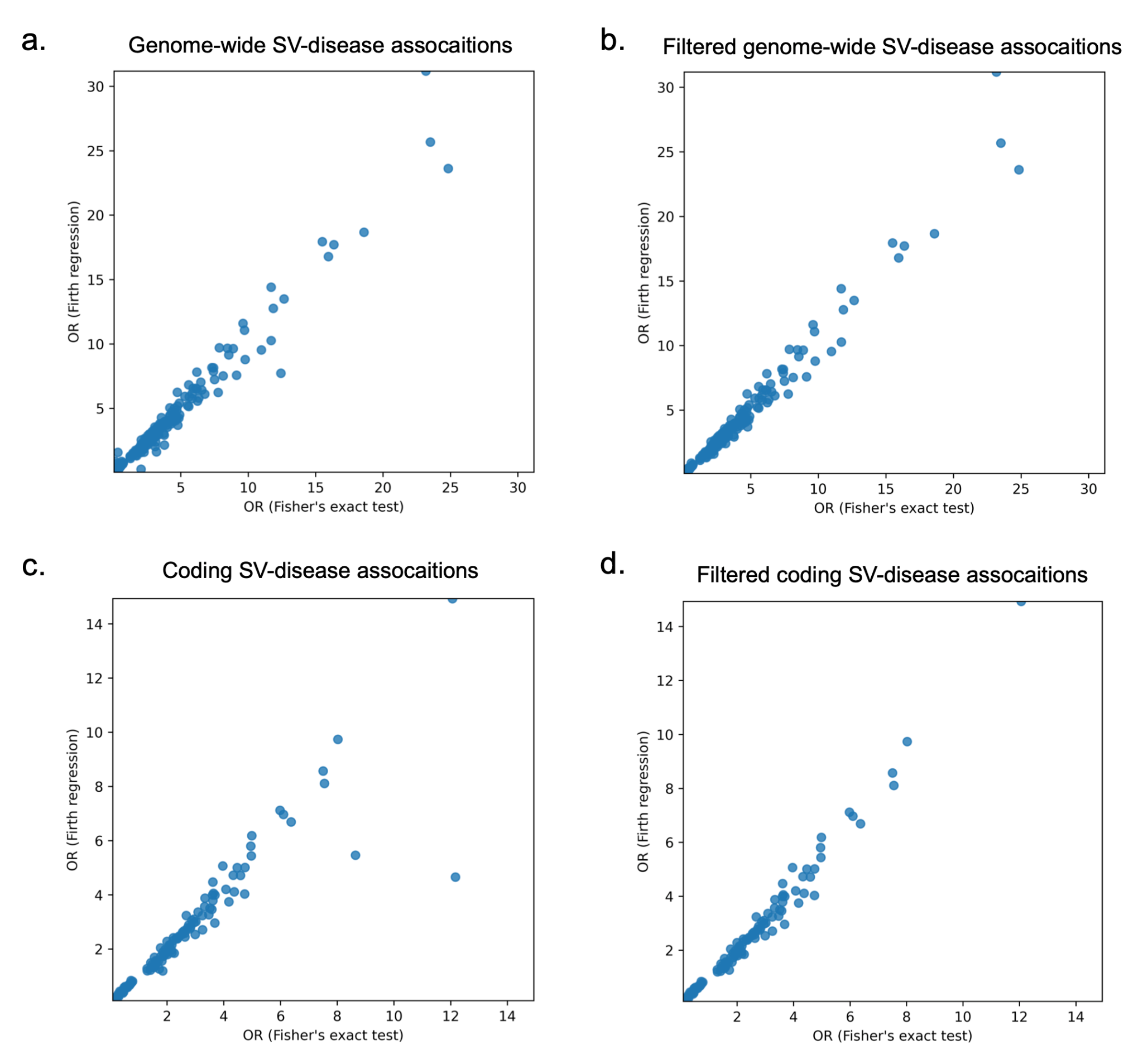

Comparison of odds ratios (ORs) derived from Fisher’s exact test and Firth regression for **a,** genome-wide SV–disease associations (n = 304), **b,** genome-wide associations filtered to those with OR differences ≤ 1.5-fold between methods (n = 291), **c,** coding SV–disease associations (n = 119), and **d,** coding associations filtered to those with OR differences ≤ 1.5-fold between methods (n = 114).

#### Supplementary Fig. 13: Variant evidence and population structure of disease-associated SVs

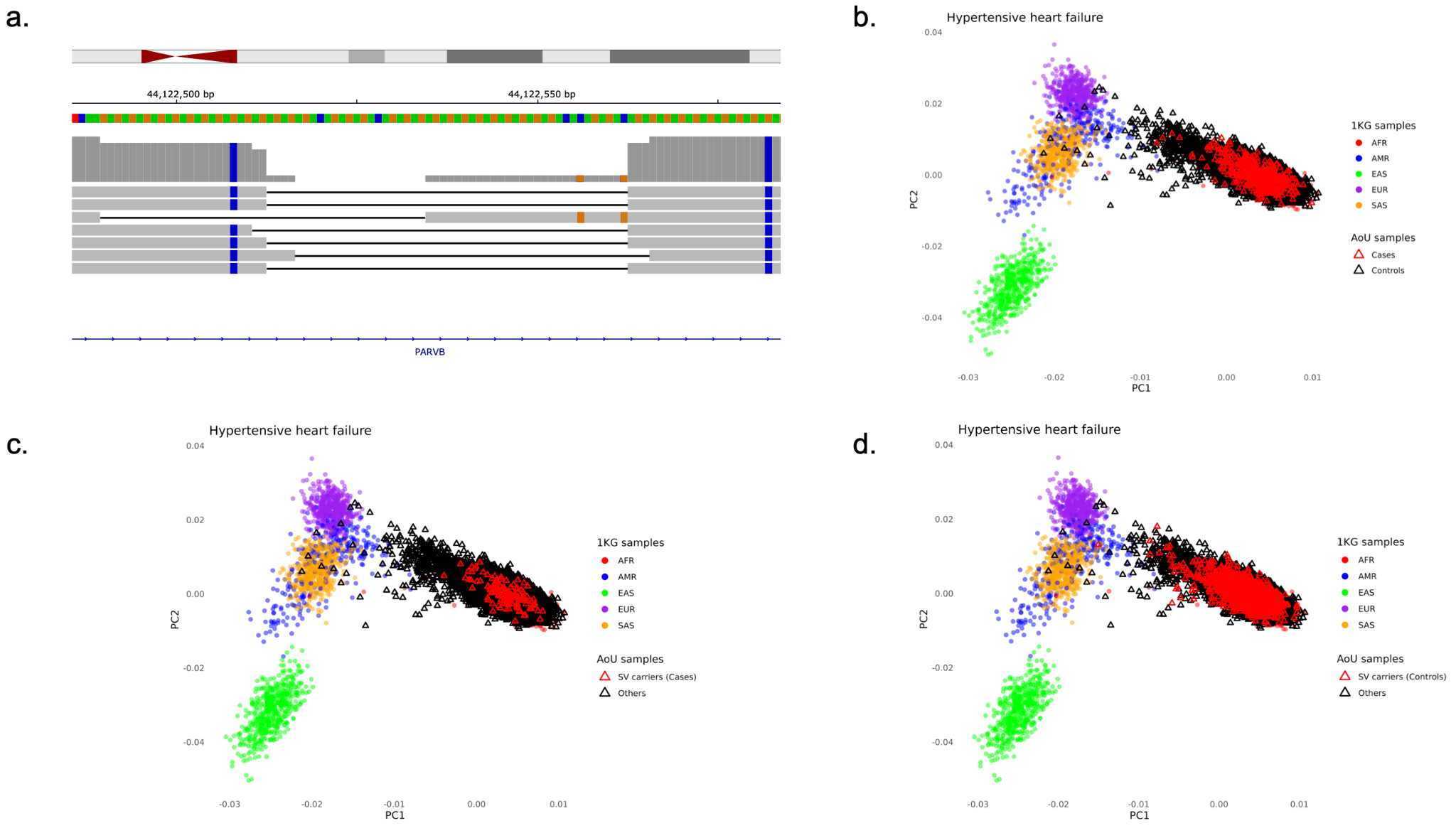

**a**, Integrative Genomics Viewer (IGV) screenshot of a 50 bp deletion associated with hypertensive heart failure. **b**, Principal component analysis (PCA) of SV genotypes from 2,504 samples in the 1000 Genomes Project and 10,000 short-read samples from the AoU cohort. Red triangles indicate AoU cases with hypertensive heart failure. **c**, Same PCA plot, with red triangles indicating AoU cases carrying the 50 bp deletion. **d**, Same PCA plot, with red triangles indicating AoU controls carrying the 50 bp deletion.

#### Supplementary Fig. 14: Clinical associations of structural variants in coding regions

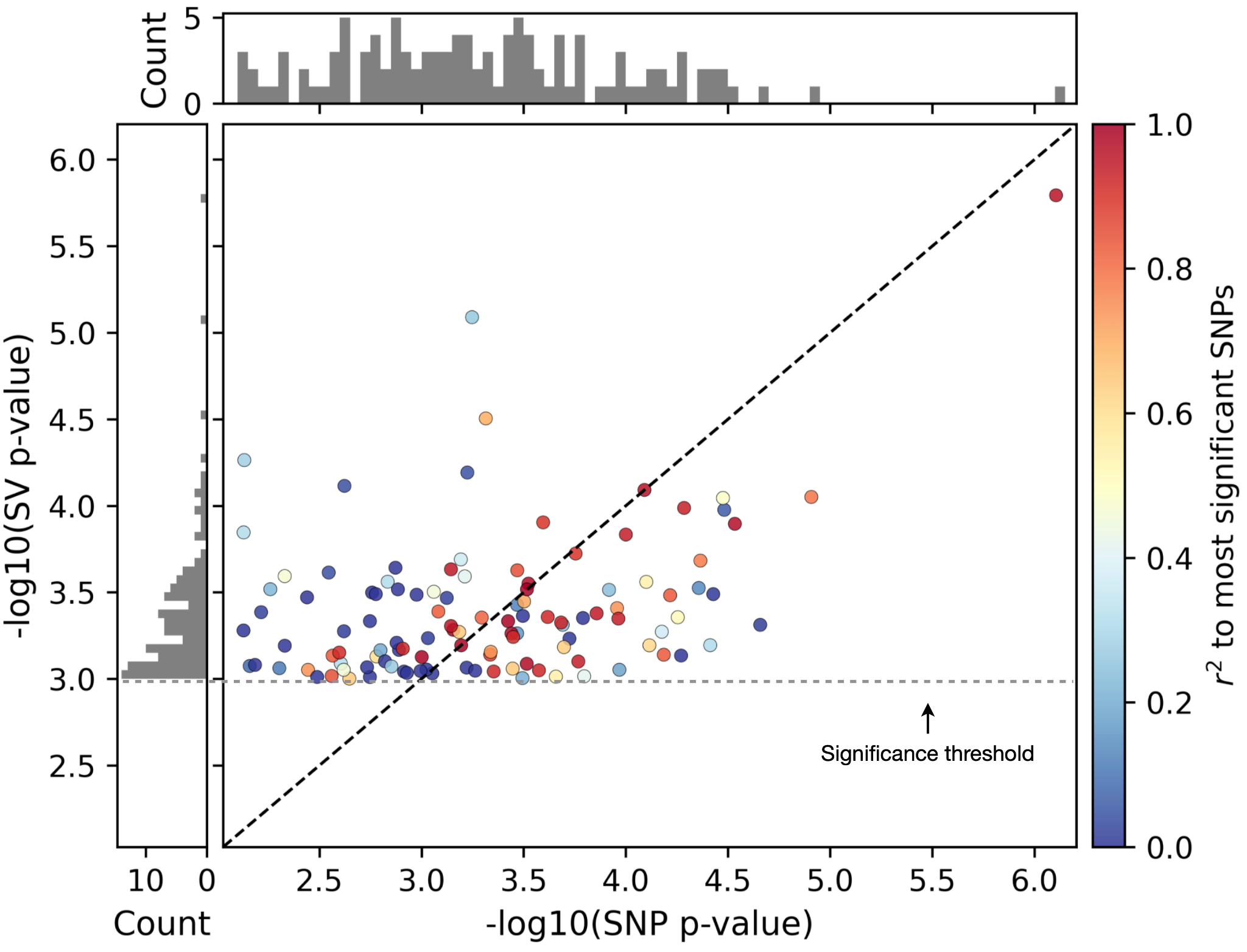

Comparison of significance levels between each coding-region SV and the most significantly associated nearby SNP (within $\pm$100 kb) for the same phenotype. Color indicates linkage disequilibrium (r^2^) with the top associated SNP; histograms show the distributions of SV and SNP p-values.

#### Supplementary Fig. 15: Characterization of an atelectasis-associated SV

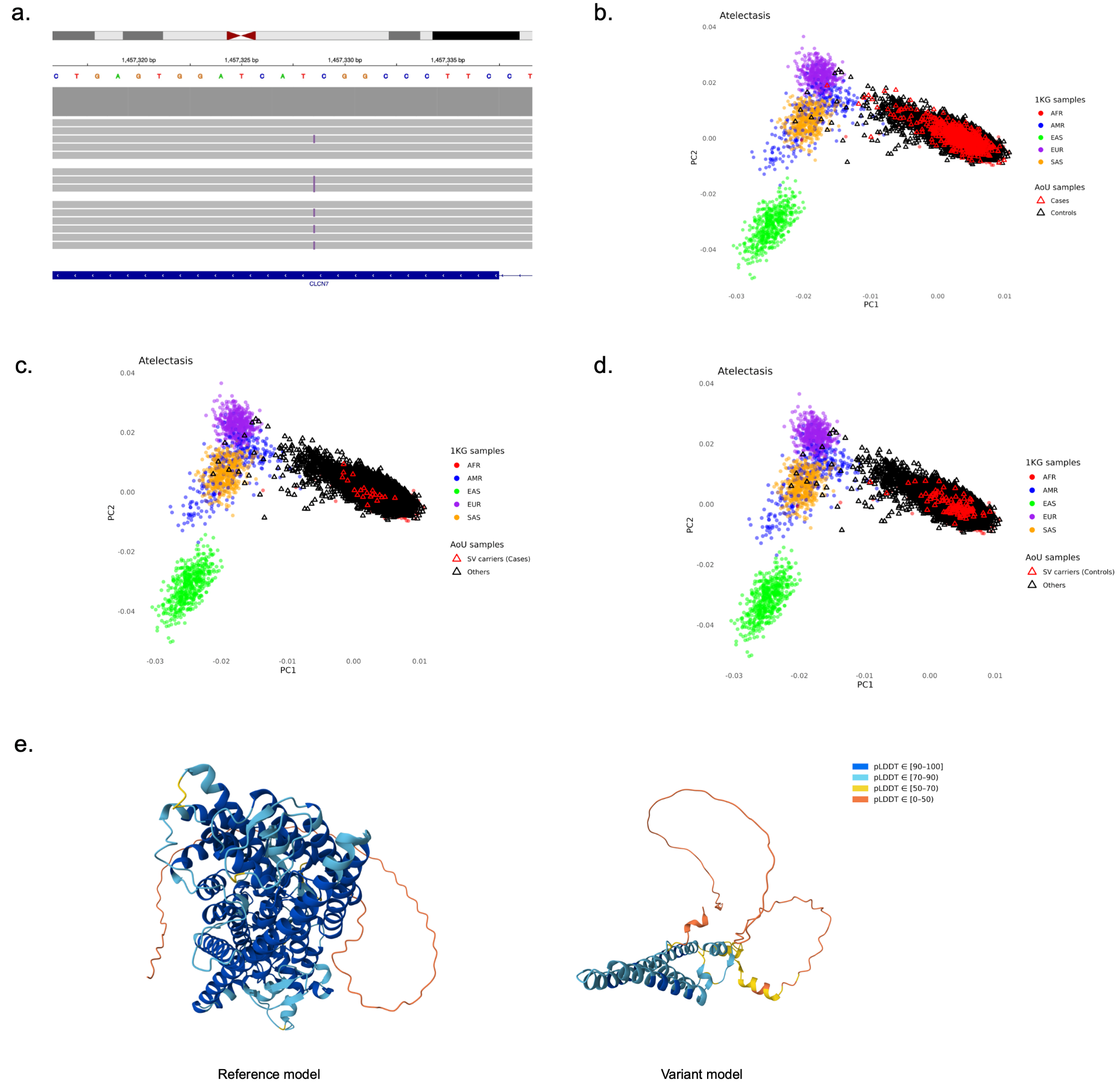

**a**, Integrative Genomics Viewer (IGV) screenshot of the 200 bp insertion in *CLCN7* associated with atelectasis. **b**, Principal component analysis (PCA) of SV genotypes from 2,504 samples in the 1000 Genomes Project and 10,000 short-read samples from the AoU cohort. Red triangles indicate AoU cases with atelectasis. **c**, Same PCA plot, with red triangles indicating AoU cases carrying the 200 bp insertion. **d**, Same PCA plot, with red triangles indicating AoU controls carrying the 200 bp insertion. **e**, Predicted structure of CLCN7 protein from AlphaFold: reference (left) and truncated form resulting from the SV (right).

#### Supplementary Fig. 16: Characterization of an SV associated with periodontal disease

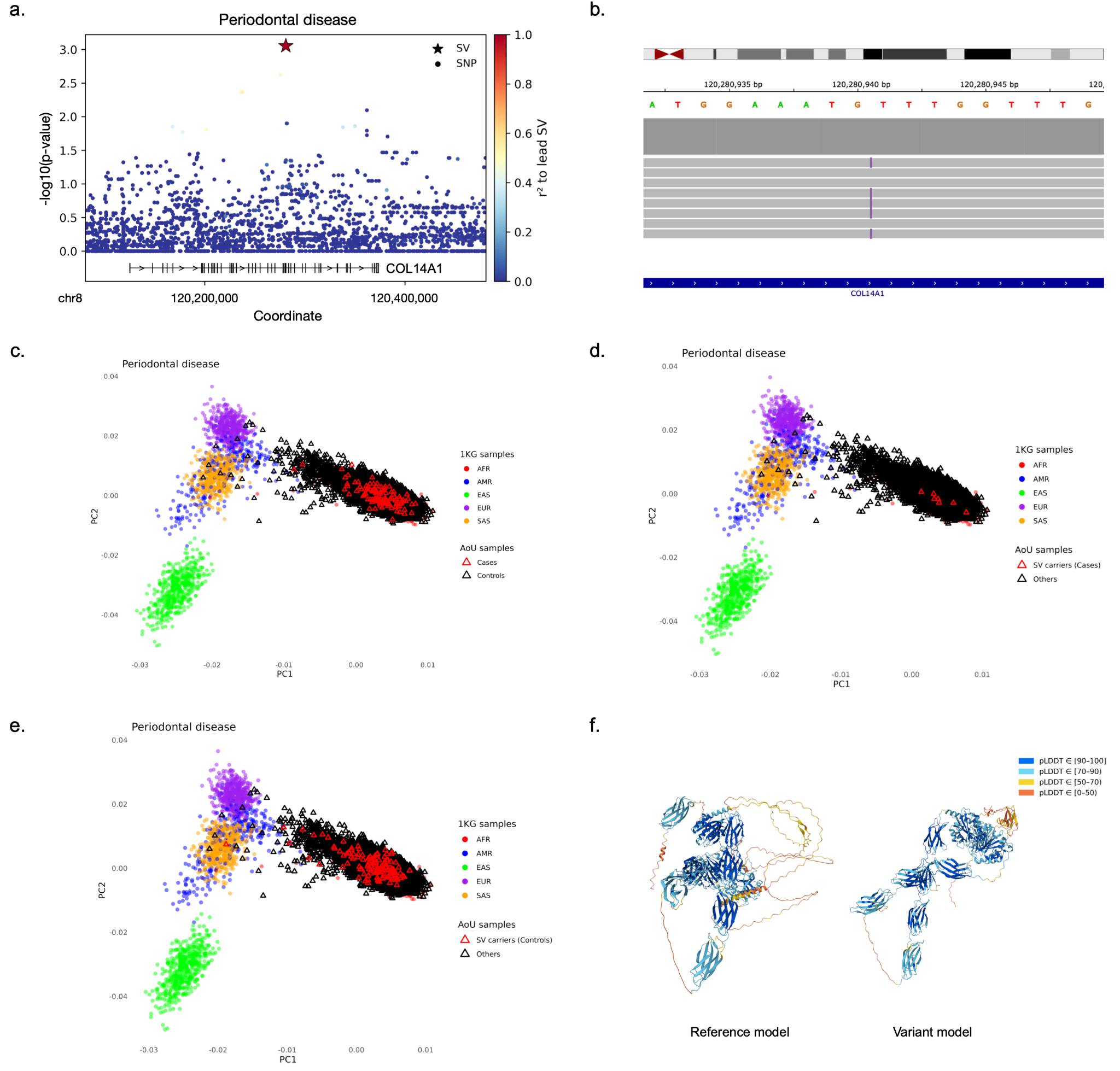

**a**, Manhattan plots showing variants associated with periodontal disease. The insertion is the lead variant, and surrounding points are colored based on the LD (r^2^) nearby SNPs and the SV. **b**, Integrative Genomics Viewer (IGV) screenshot of a 190 bp insertion in *COL14A1* linked to periodontal disease. **c**, Principal component analysis (PCA) of SV genotypes from 2,504 samples in the 1000 Genomes Project and 10,000 short-read samples from the AoU cohort. Red triangles indicate AoU cases with periodontal disease. **d**, Same PCA plot, with red triangles indicating AoU cases carrying the 190 bp insertion. **e**, Same PCA plot, with red triangles indicating AoU controls carrying the 190 bp insertion. **f**, Predicted structure of protein from AlphaFold: reference (left) and truncated form resulting from the SV (right).

| Supplementary Fig. 17: Heterozygous-variant counts for AoU+HPRC panel and 3,202 1kGP imputed samples | | |
| --- | --- | --- |
| 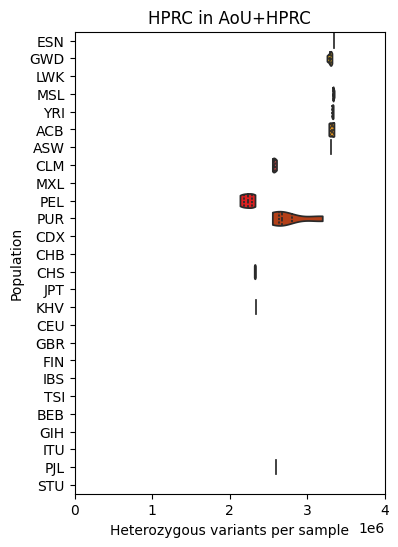 | 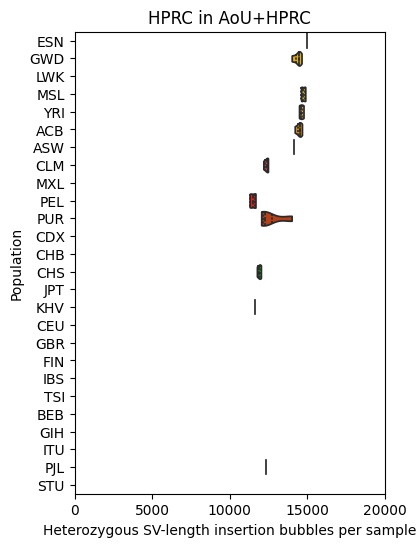 | 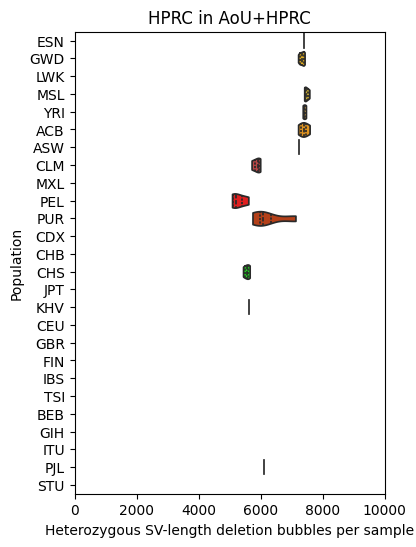 |
| 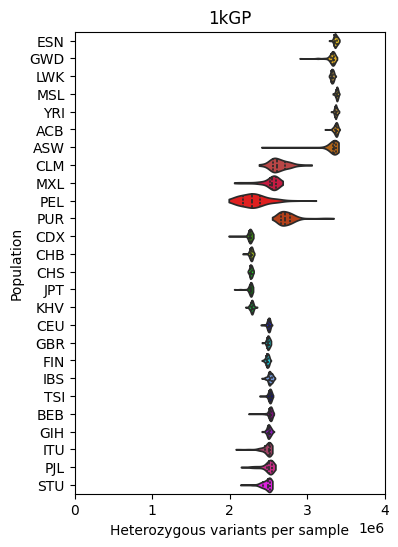 | 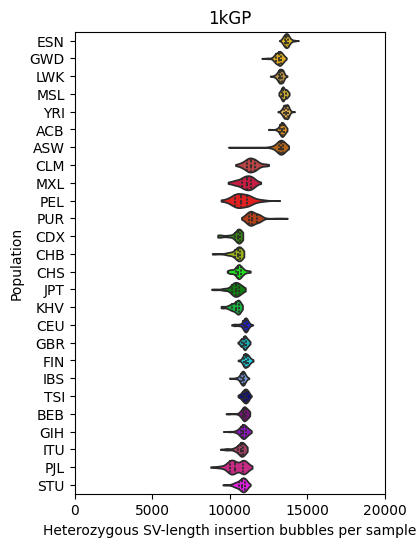 | 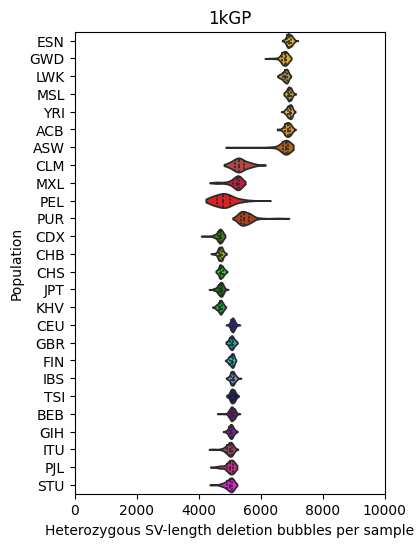 |
| Top row: Unfiltered counts of heterozygous variants per sample (left, all variants; center/right, SV-length insertion/deletion bubbles), stratified by population over the 47 HPRC samples in the AoU+HPRC panel.  Bottom row: The same for 3,202 1kGP samples imputed against the AoU+HPRC panel. | | |

| Supplementary Fig. 18: Allele-frequency correlation and Hardy-Weinberg equilibrium for AoU+HPRC panel and 3,202 1kGP imputed samples | | |
| --- | --- | --- |
| 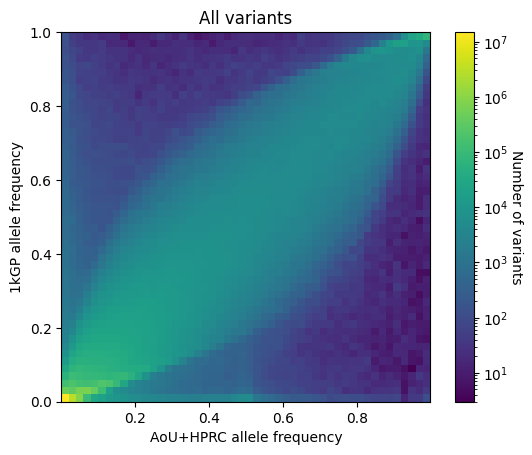 | 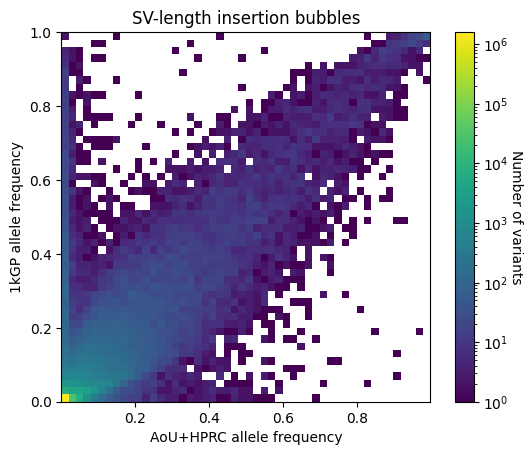 | 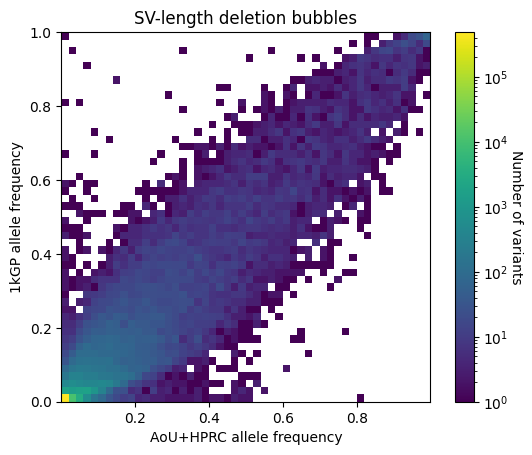 |
| 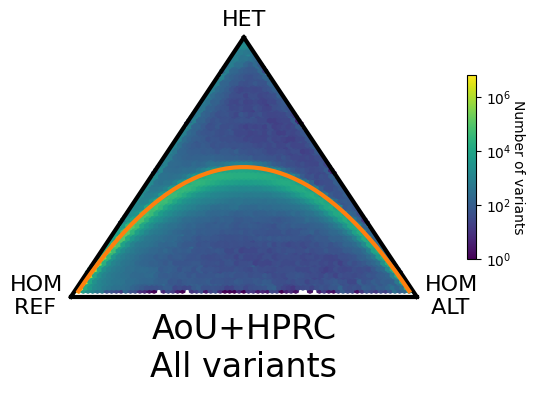 | 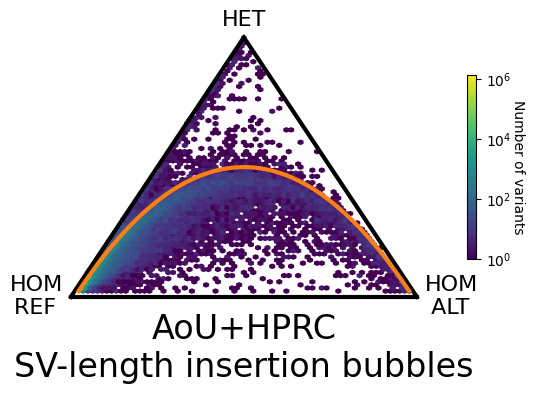 | 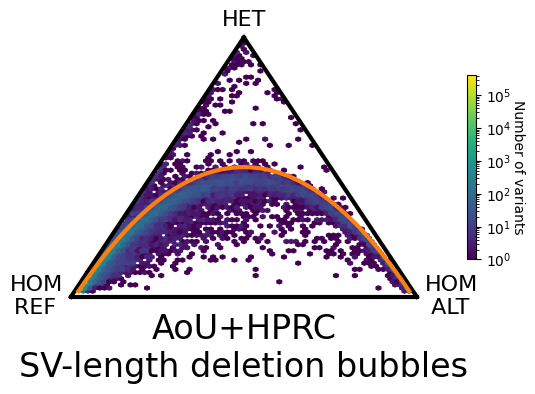 |
| 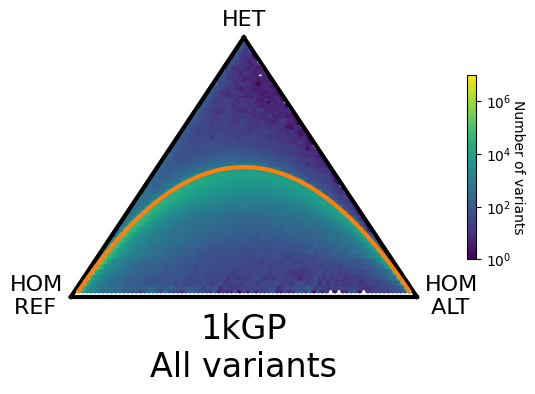 | 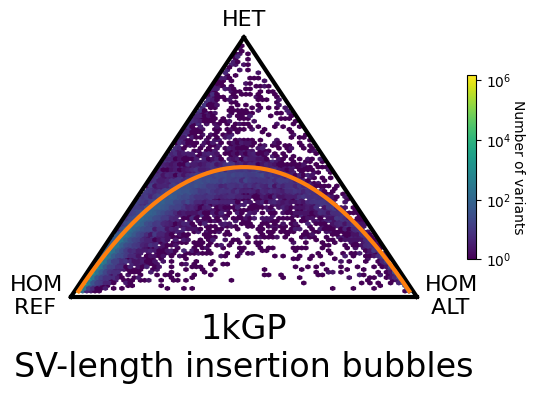 | 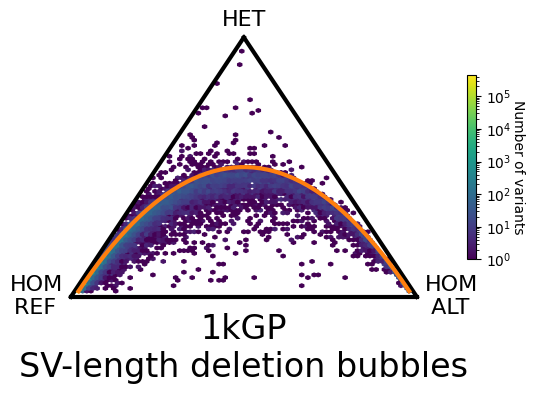 |
| Top row: Heatmaps of allele-frequency correlations from the AoU+HPRC panel (x-axis) and 3,202 unfiltered 1kGP samples imputed against it (y-axis), for all variants (left) and SV-length insertion/deletion (center/right) bubbles; Pearson correlation coefficient = 0.94, 0.84, and 0.94, respectively.  Middle and bottom rows: Di Finetti heatmaps of unfiltered genotype frequencies with Hardy-Weinberg equilibrium shown in orange, again stratified from left-to-right by variant type, for both the AoU+HPRC panel (middle row) and the 3,202 1kGP imputed samples (bottom row). Bubbles have been split to biallelic variants in all plots. | | |

| Supplementary Fig. 19: Allele-frequency correlation and Hardy-Weinberg equilibrium for AoU+HPRC panel and 10,000 self-identified Black or African American AoU imputed samples | | |
| --- | --- | --- |
| 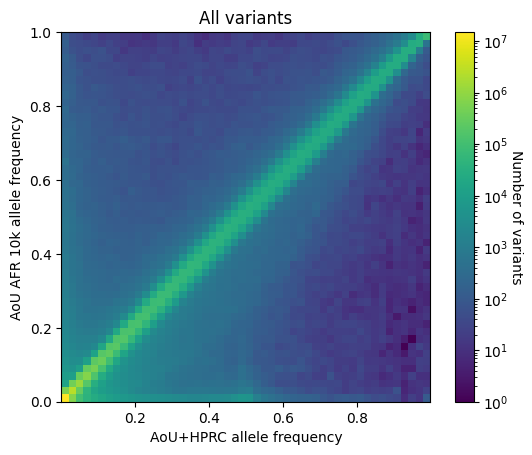 | 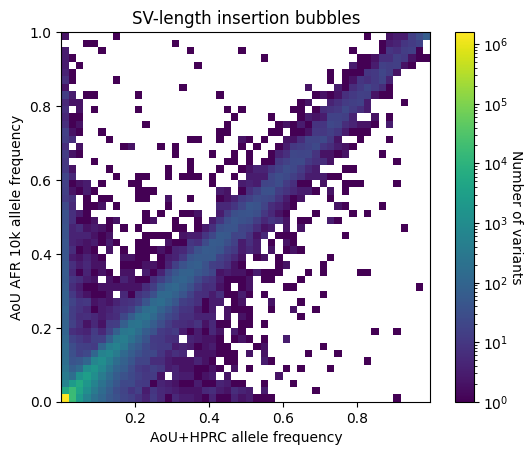 | 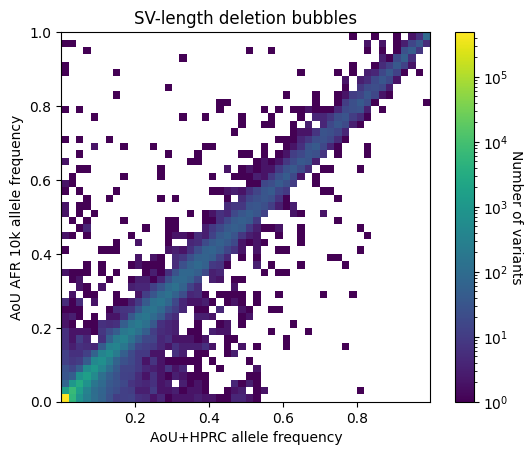 |
| 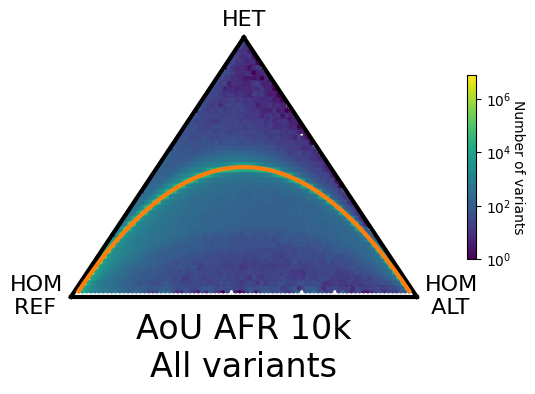 | 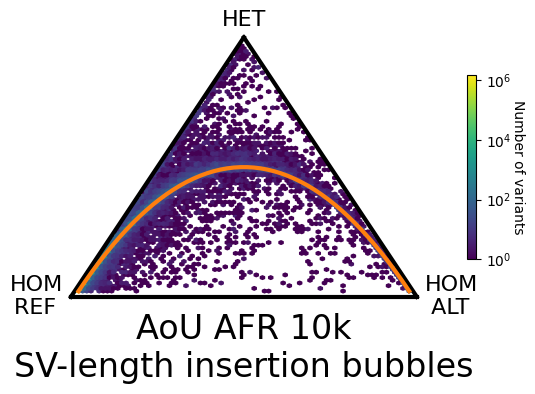 | 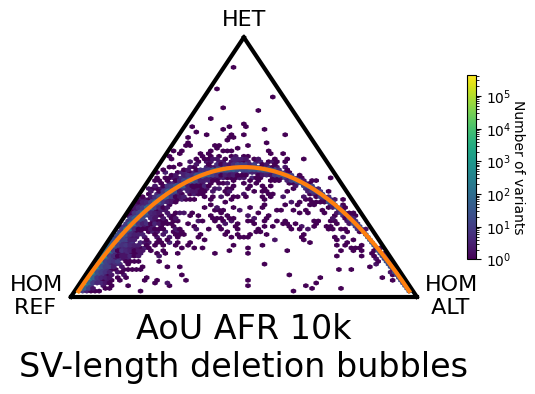 |
| The same as Fig. 18, but for 10,000 unfiltered self-identified Black or African American AoU imputed participants. Allele-frequency Pearson correlation coefficient = 0.98, 0.91, and 0.98, respectively, with higher values than the corresponding for 1kGP reflecting the effect of genetic ancestry matching. | | |

#### Supplementary Fig. 20: Comparison of KAGE+GLIMPSE and PanGenie accuracy in leave-out evaluations with HGSVC2 (assembly-based) and AoU+HPRC (8x LRS) panels

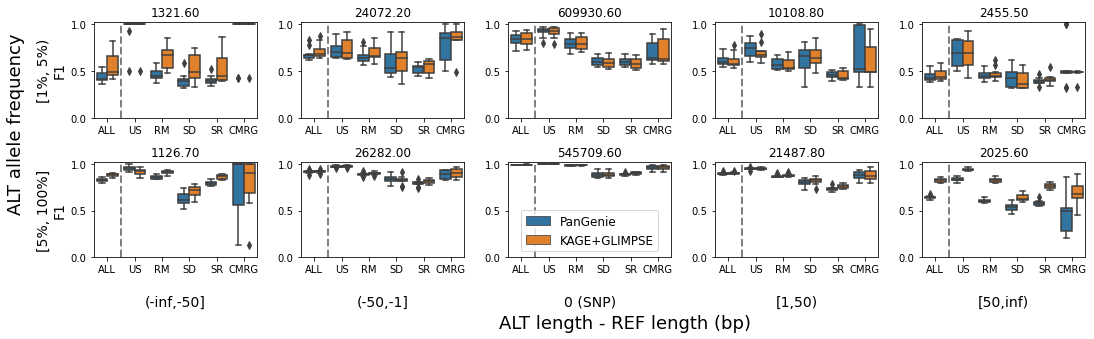

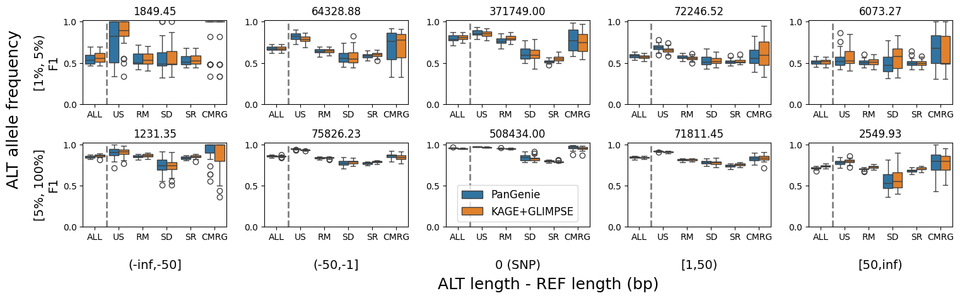

Unfiltered KAGE+GLIMPSE and PanGenie F1-scores, calculated from macro-averaged, exact-match genotype concordance and stratified by variant type, allele frequency, and genomic context (US = unique sequence, RM = RepeatMasker track, SD = segmental-duplications track, SR = simple-repeats track^48–51^. Boxplots indicate the score distribution across leave-out samples in chr1. The average number of biallelic variant bubble alleles evaluated per sample in each stratification is given at the top of each plot.

Top row: Results from using 10 samples in the HGSVC2 assembly-based panel in 10 leave-one-out experiments. PanGenie v2.1.1, which only supports panels containing up to 127 samples, is used.

Bottom row: Results from using 40 HPRC samples in our AoU+HPRC LRS panel in a single leave-many-out experiment. PanGenie v4.1.1, which now supports panels containing more than 127 samples, is used.

#### Supplementary Fig. 21: Imputation dosage r^2^ in a leave-many-out evaluation over 40 HPRC samples

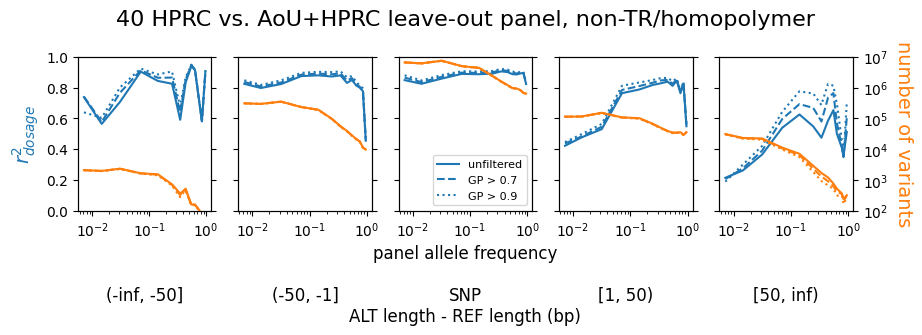

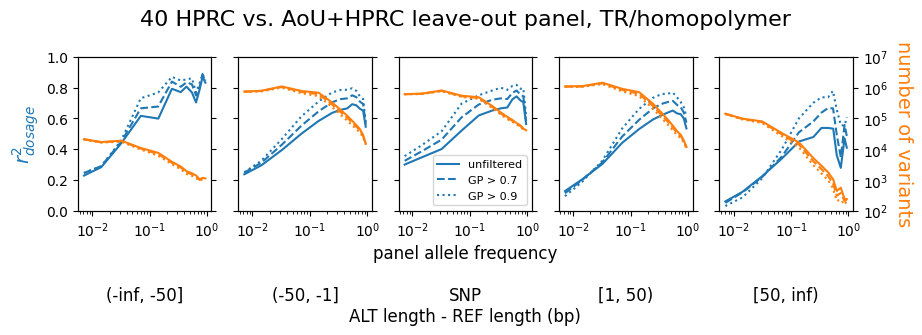

Imputation dosage r^2^ as a function of panel allele frequency, averaged over 40 HPRC samples in our AoU+HPRC LRS panel used in a single leave-many-out experiment over chr6 and stratified by variant type and repeat/homopolymer context. The number of biallelic variant bubble alleles as a function of panel allele frequency in each stratification is also shown, as are the effects of genotype-level filtering by maximum genotype posterior at various thresholds. Filtering has a larger effect for SV insertions (which may reflect the strictness of exact genotype match across all samples in each trio) and in repeat/homopolymer regions.

| Supplementary Fig. 22: Non-reference concordance rate in a leave-many-out evaluation over 40 HPRC samples | |
| --- | --- |
| 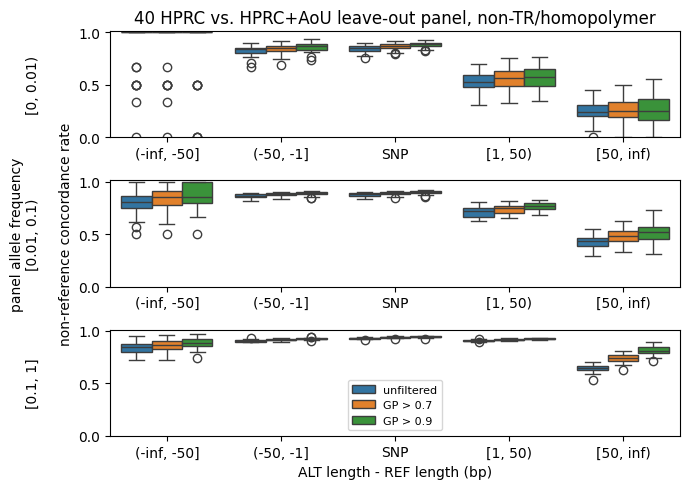 | 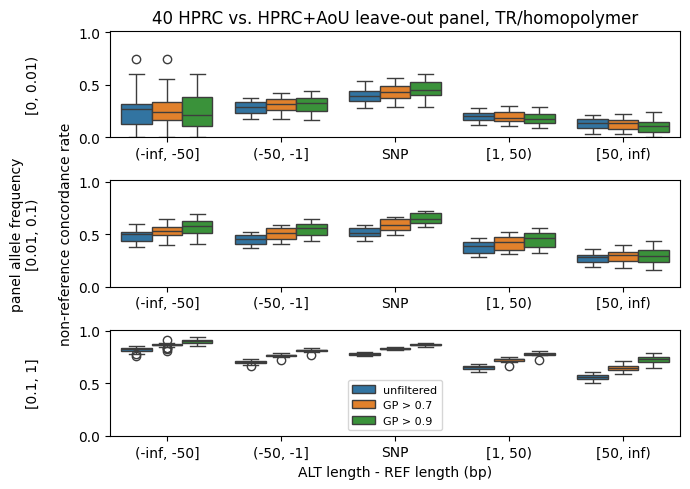 |
| Non-reference concordance rate over 40 HPRC samples in our AoU+HPRC LRS panel used in a single leave-many-out experiment over chr6, stratified by variant type, panel allele frequency, and GIAB tandem-repeat/homopolymer context. As in Fig. 21, biallelic bubble alleles are evaluated using exact genotype matching and similar trends in filtering by maximum genotype posterior are also observed. | |

| Supplementary Fig. 23: Accuracy of imputation panel and cases against assembly-based truth, outside of TR/homopolymer regions | |
| --- | --- |
| 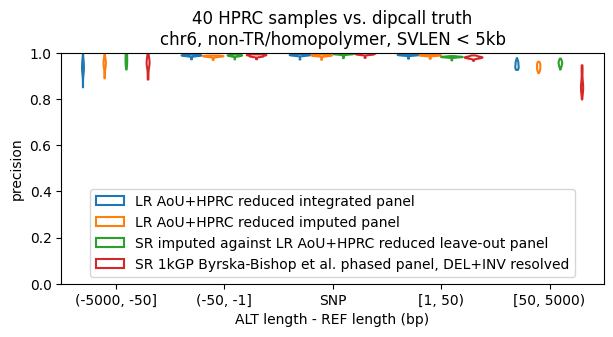 | 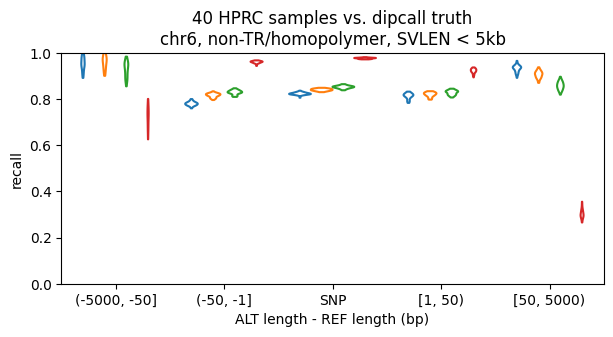 |
| 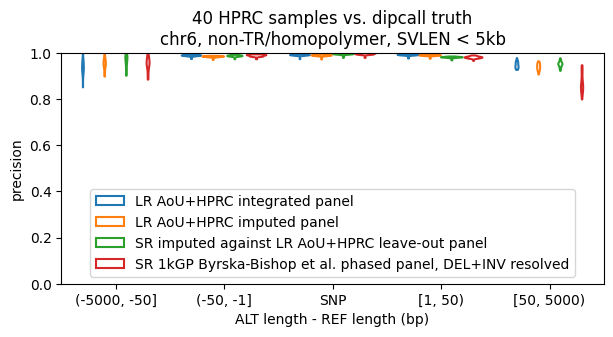 | 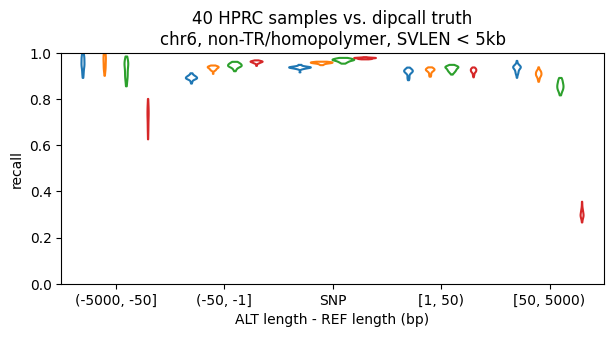 |
| Vcfdist precision and recall in chr6 dipcall-confident regions outside of GIAB tandem-repeat/homopolymer regions over 40 ground-truth HPRC samples for reduced (top row) and full (bottom row) AoU+HPRC LRS integrated and imputed panels, unfiltered HPRC srWGS cases imputed against the corresponding leave-many-out panel, and the Byrska-Bishop et al. 1kGP srWGS panel. Unlike the methods for computing metrics based on exact genotype or Mendelian concordance in Figs. 20-22 and 25, Vcfdist assigns partial credit and allows alignment-based comparison between the dipcall (truth) and bubble (query) variant representations. | |

| Supplementary Fig. 24: Accuracy of imputation panel and cases against assembly-based truth, inside of TR/homopolymer regions | |
| --- | --- |
| 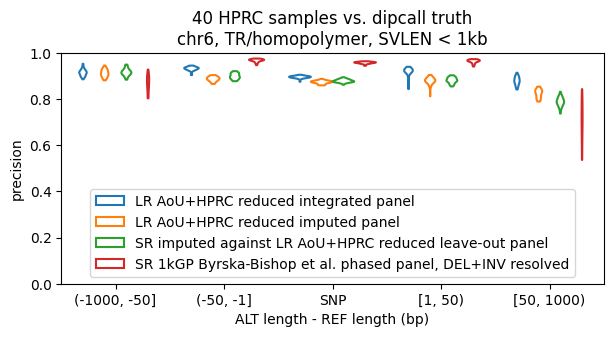 | 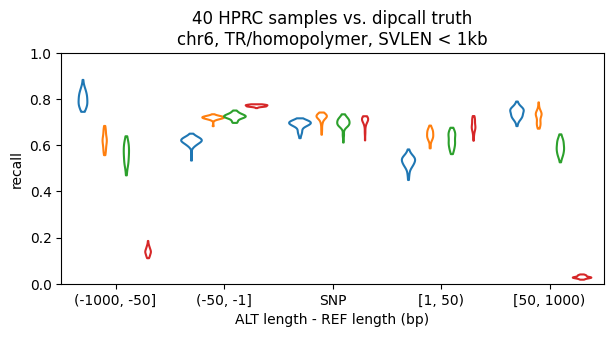 |
| 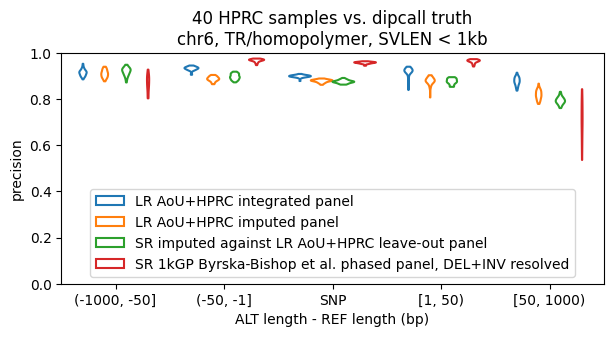 | 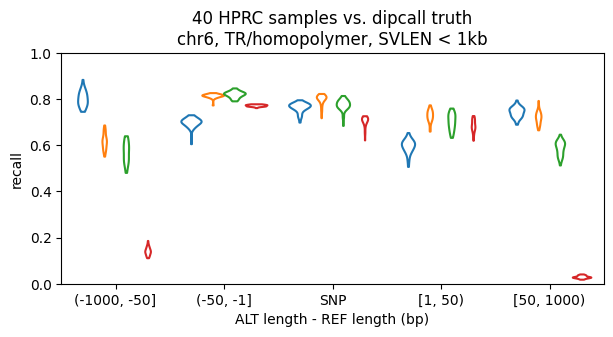 |
| The same as Fig. 23, but inside of GIAB tandem-repeat/homopolymer regions. | |

| Supplementary Fig. 25: Mendelian consistency for 602 1kGP imputed trios | |
| --- | --- |
| 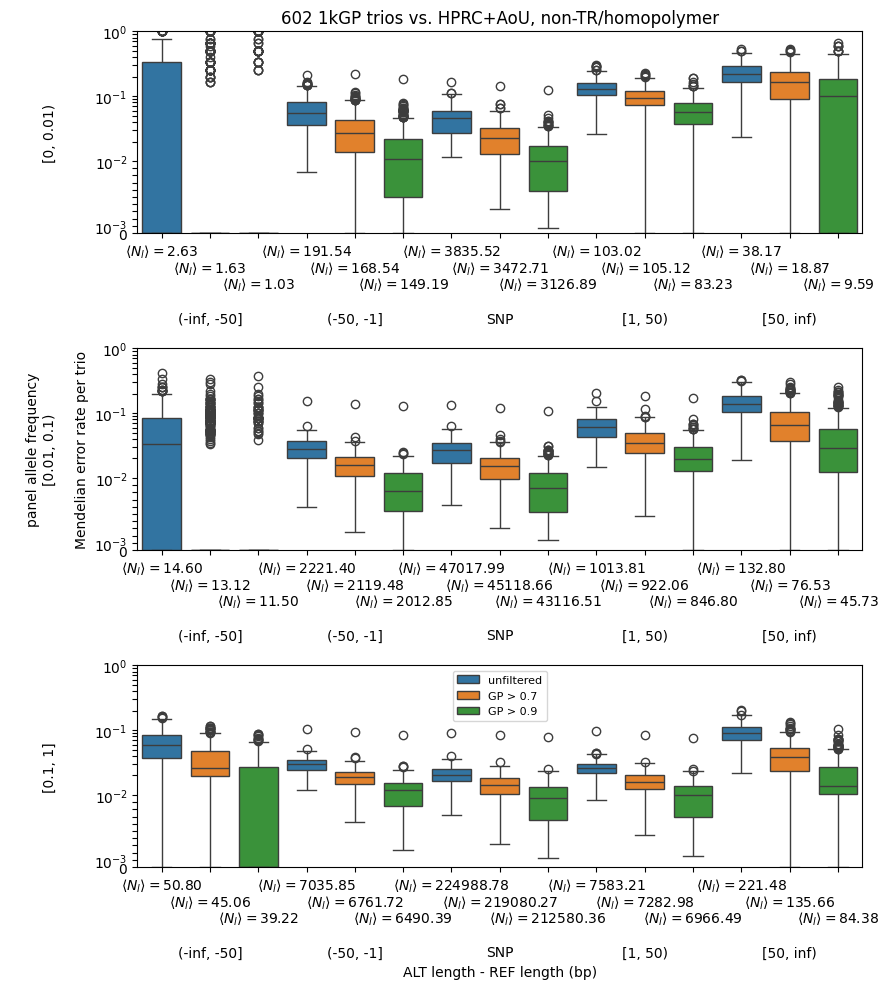 | 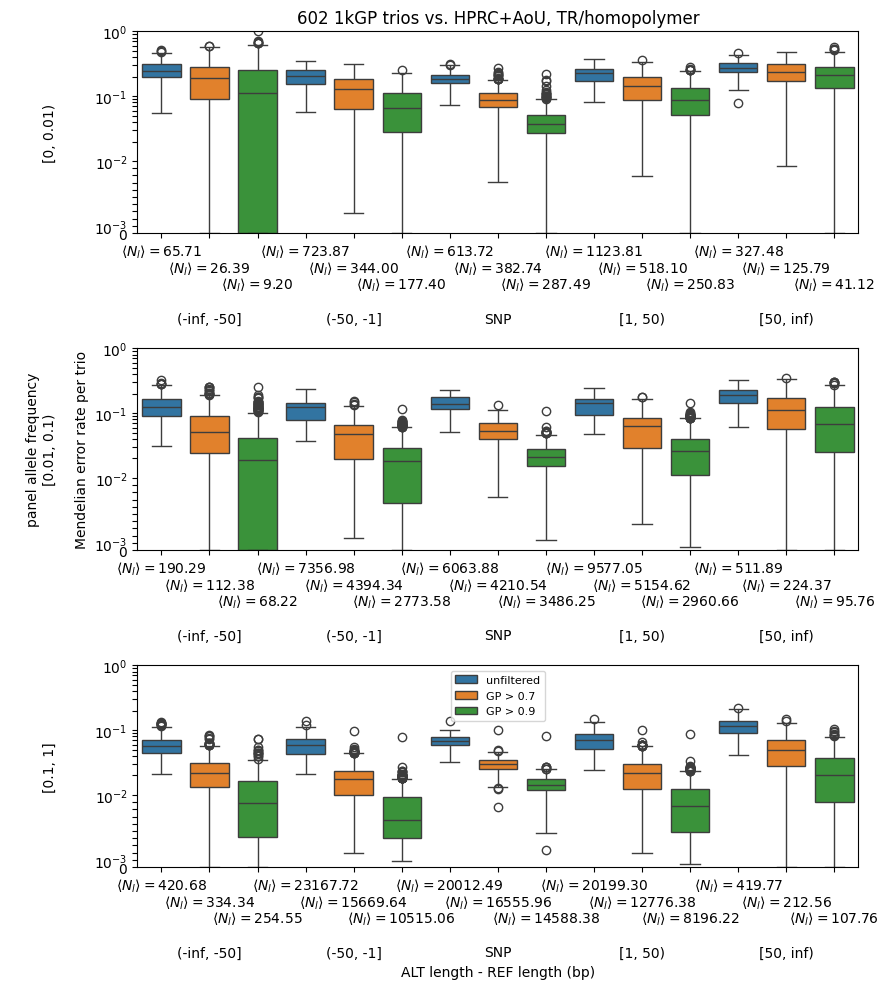 |
| 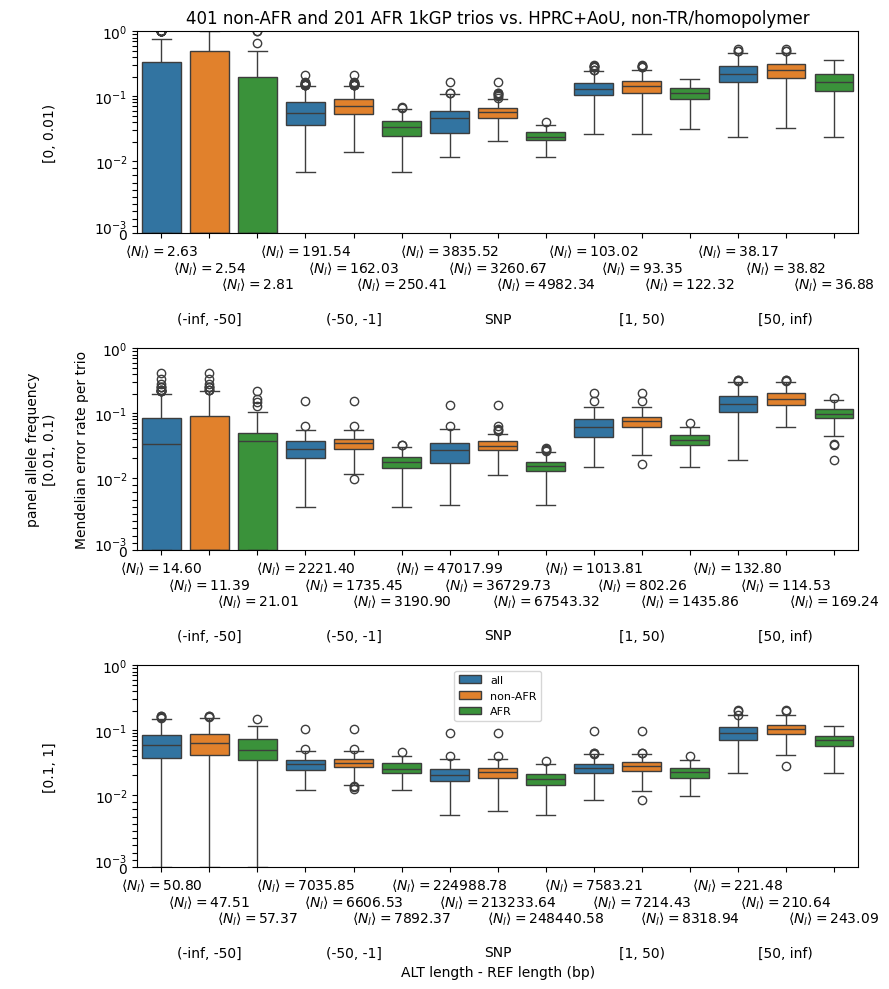 | 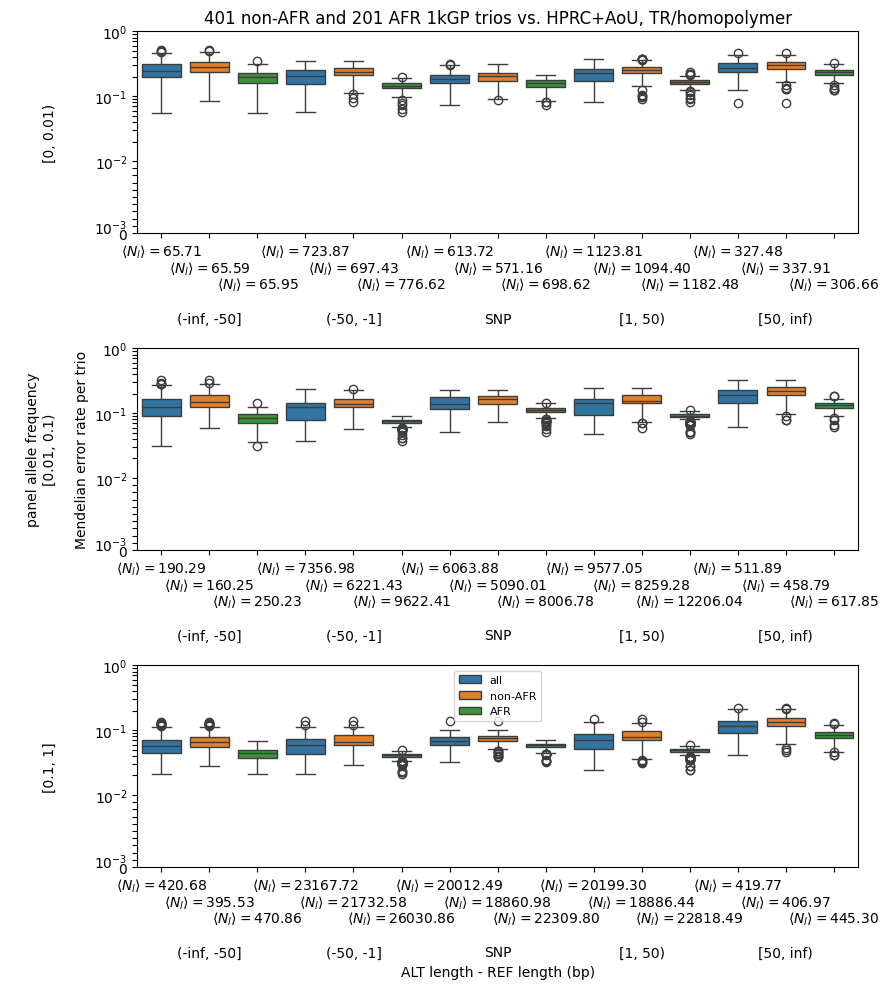 |
| Mendelian error rate in chr6 regions inside (left) and outside (right) of GIAB tandem-repeat/homopolymer regions over 602 1kGP trios, stratified by variant type and panel allele frequency. Filtering by maximum genotype posterior (top row) or stratifying trios by African genetic ancestry (AFR) / non-African genetic ancestry (Non-AFR) (bottom row) yields expected trends in accuracy. Error rates are defined using exact genotype matching over biallelic bubble alleles over loci where all members of a trio are non-reference and unfiltered; the mean number of such loci per trio, *<N_l_>*, is given for each stratification. | |

### REFERENCES

1.	*HG002: A Complete Diploid Human Genome*. (https://github.com/marbl/HG002).

2.	GIAB HG002 GRCh38 Assembly-Based Small and Structural Variants Draft Benchmark Sets. <https://ftp-trace.ncbi.nlm.nih.gov/ReferenceSamples/giab/data/AshkenazimTrio/analysis/NIST_HG002_DraftBenchmark_defrabbV0.011-20230725/README.md>.

38.	*Ivcfmerge: A Utility to Merge a Large Number of VCF Files Incrementally*. (Github).
